# Deep learning predicts onset acceleration of 38 age-associated diseases from blood and body composition biomarkers in the UK Biobank

**DOI:** 10.1101/2025.03.16.25323714

**Authors:** Mica Xu Ji, Marjola Thanaj, Léna Nehale-Ezzine, Brandon Whitcher, E. Louise Thomas, Jimmy D. Bell

## Abstract

A major challenge in multimorbid aging is understanding how diseases co-occur and identifying high-risk groups for accelerated disease development, but to date associations in the relative onset acceleration of disease diagnoses have not been used to characterise disease patterns. This study presents the development and evaluation of a neural network Cox model for predicting onset acceleration risk for age-associated conditions, using demographic, anthropomorphic, imaging and blood biomarker traits from 60,396 individuals and 218,530 outcome events from the UK Biobank. Risk prediction was evaluated with Harrell’s concordance index (C-index). The model performed well on internal (C-index **0.6830 *±* 0.0902**, ***n* = 8, 931**) and external (C-index **0.6461 *±* 0.1264**, ***n* = 855**) test sets, attaining C-index ***≥* 0.6** on 38 out of 47 (**80.9%**) conditions. Inclusion of body composition and blood biomarker input traits were independently important for predictive performance. Kaplan-Meier curves for predicted risk quartiles (log-rank ***p ≤* 1.16*E −* 16**) indicated robust stratification of individuals into high and low risk groups. Analysis of risk quartiles revealed cardiometabolic, vascular-neuropsychiatric, and digestive-neuropsychiatric disease clusters with strong statistically significant inter-correlated onset acceleration (***r ≥* 0.6**, ***p ≤* 3.46*E −* 5**), while 13 and 19 conditions were strongly associated with onset acceleration of all cause mortality and all cause morbidity respectively. In prognostic survival analysis, the proportional hazards assumption was met (Schoenfeld residual ***p >* 0.05**) in 435 out of 435 or 100% (1238 out of 1334 or 92.8%) of cases across outcomes, ***aHR* = 6.11 *±* 9.00** (***aHR* = 3.67 *±* 5.78**) with (without) Bonferroni correction. The neural architecture of OnsetNet was interpreted with saliency analysis and several significant body composition and blood biomarkers were identified. The results demonstrate that neural network survival models are able to estimate prognostically informative onset acceleration risk, which could be used to improve understanding of synchronicity in the onset of age-associated diseases and reprioritize patients based on disease-specific risk.

## 1 Introduction

As we age, the probability of developing more than one condition greatly increases [1]. Multimorbidity, which can be defined as co-occurrence of more than one condition in the same individual, is thus an important problem in aging societies [2, 3], being associated with lower quality of life [4], higher mortality risk [5], and greater usage of healthcare resources than healthy people or those with just one chronic condition [6, 7]. There is a high and increasing prevalence of multimorbidity in the general population, with elderly and socioeconomically deprived groups particularly at risk [1, 8–10]. Chowdhury et al. [11] determined that the global prevalence of multimorbidity was 37.2%, rising to 51.0% in people over 60 years old, while Nunes et al. [12] found that mortality risk of individuals with multimorbidity was 1.73 times greater than those without multimorbidity. Despite these concerns, healthcare delivery systems and quality evaluation remain focused on the treatment of single diseases [13, 14]. An important part of generating an evidence base for improvements in clinical practice is ascertaining patterns of disease and inter-disease associations beyond chance [15].

While epidemiological studies of multimorbidity vary widely in sample size, age settings and statistical methodology, there is a growing consensus that age-associated chronic diseases do not co-occur randomly but in associated fashion, including conditions across the physical and mental health divide [1, 16]. A 2014 meta-review of 14 studies [15] identified cardiometabolic, neuropsychiatric-gastrointestinal, and musculoskeletal-vascular-neuropsychiatric-gastrointestinal co-occurrence clusters, while more recently [17] identified cardiometabolic, neuropsychiatric, and allergic disease co-occurrence clusters as being commonly reported among 41 studies. The majority of reviewed studies used agglomeration methods such as agglomerative hierarchical clustering and explanatory factor analysis to link diseases on the basis of co-occurrence in individuals, and were not restricted to age-associated conditions. While co-occurrence is important for characterising disease associations, another significant factor is age of disease onset [1]. For example, whether an individual develops cardiovascular disease or hearing loss may be less informative for their health status than age of onset, given that prevalence is estimated at 79-86% and 80.6% in those older than 80 and 85 respectively in the United States [18, 19]. The timing of disease onset has been shown to be informative for risk of secondary diagnoses; early onset cancer is associated with increased risk of cardiovascular disease and secondary cancers compared to later onset cancer [20, 21]. Accelerated biological age estimated using DNA methylation or phenotypic aging clocks has been associated with increased risk of mortality and adverse health outcomes compared to less accelerated biological age [22, 23]. A recent study on chronic diseases [24] found that certain lifestyle factors affected only age of onset and not odds of diagnosis for 10 cardiometabolic, autoimmune and respiratory conditions. Taken together, this suggests that the temporal dimension of multimorbidity may be important for characterising disease patterns.

The present study aims to determine (1) the extent to which onset acceleration risk of age-associated diseases can be accurately predicted from individual traits, and (2) how strongly early onset risk of one disease is associated with early onset risk of another disease beyond chance. To this end, Cox models were trained and evaluated for prediction of individual risk for early onset of diverse age-associated diseases. Using data from 60, 396 individuals in the UK Biobank imaging cohort [25], we trained a single multimodal neural network, OnsetNet, to predict time-to-event log-risk of the first diagnosis of 47 conditions with birth as temporal baseline, including two composite conditions of all-cause mortality and all-cause morbidity.

First, we evaluated performance of neural network models across disease conditions using C-index measured on internal center and external center held-out test sets, where model hyperparameters were selected by fivefold cross-validation and internal test and C-index ≥ 0.6 was considered acceptable prediction [26]. We compared performance of neural network models on eight different combinations of five input trait groups (demographic, anthropomorphic, summary body composition, spatial body composition, and blood biomarkers) totalling 43 input variables. We assessed performance differences between neural network and linear architectures to evaluate the benefit of using a neural network, which is state-of-the-art and may be a more appropriate choice where it is too simplistic to assume linearity of the log-risk function [27]. OnsetNet score quartiles were computed to illustrate within-disease heterogeneity across onset acceleration risk quartiles. Between-disease risk relationships were analysed by computing all-to-all correlations in OnsetNet risk quartiles between diseases, and clusters containing disease conditions with high inter-correlated onset acceleration risk were identified with agglomerative hierarchical clustering. We trained standard prognostic linear adjusted Cox models (baseline as measurement time, pre-existing diagnoses excluded) with OnsetNet score as an input trait, yielding adjusted hazard ratios across disease pairs. The proportional hazards assumption was tested by examination of Schoenfeld residuals. Finally, the model was analysed with gradient-based saliency to infer relative importance of individual biomarkers and to assess for biological validity of learned associations between biomarkers and disease risk.

OnsetNet’s output is interpretable as a metric of accelerated biological aging, given that accelerated biological age can be characterized as event acceleration risk of age-associated disease from birth [28]. While the present study is purely associative, OnsetNet’s ability to stratify individuals into high and low risk groups may be useful for clinical settings, where important problems include early identification of high-risk individuals in order to effectively reduce disease burden on the general population [29] and characterising within-disease heterogeneity due to diversity in pathology [30, 31].

## 2 Methods

### 2.1 Study participants

This study was a secondary analysis of longitudinal data from the UK Biobank resource [32] under application number 44584. Data was obtained from the UK Biobank imaging cohort [33] for 60,396 individuals who were initially assessed between April 2007 and October 2010 and had outcomes recorded up to November 30*^th^* 2022 (follow-up 13.9 ± 0.819 years, min 12.2 years, max 15.6 years). Eligible individuals had full records of all covariates (29,873 male, 30,523 female, age at imaging 66.2 ± 8.01 years). Where multiple measurements were made, the latest measurement for each individual was taken. Data was split into a training and validation dataset (train-val) containing individuals assessed at Cheadle, Reading, Newcastle and unspecified centers (24,984 male, 25,626 female, age at imaging 66.2 ± 8.00 years), an internal test set containing individuals assessed at train-val centers (4,455 male, 4,476 female, age at imaging 66.0 ± 8.10 years), and an external test set contained individuals assessed at the Bristol center (434 male, 421 female, age at imaging 67.9 ± 7.54 years).

### 2.2 Covariates

Input variables were selected based on risk factors identified in literature review and were separated into 5 groups. Demographic (“demog”) contained sex, race, education level, and Townsend deprivation index. Basic body (“bb”) contained weight, height, body mass index (BMI), waist circumference, grip strength, systolic blood pressure, alcohol consumption, and smoking history. Basic body composition computed from neck-to-knee magnetic resonance imaging scans (“bbc”) contained total mass (MASS), total subcutaneous adipose tissue volume (SAT), total muscle volume (MUSC), total visceral adipose tissue volume (VAT), and total thigh intramuscular and intermuscular adipose tissue mass (TMAT). Multidimensional spatial body composition (“sbc”) contained MASS, SAT, MUSC, VAT, and TMAT measured along 370 slices across the vertical height dimension. Blood-based biomarkers (“blood”) contained low-density lipoprotein cholesterol (LDL-C), triglycerides, apolipoprotein A (ApoA), apolipoprotein B (ApoB), cholesterol, high-density lipoprotein cholesterol (HDL-C), glucose, glycated haemoglobin (HbA1c), insulin-like growth factor 1 (IGF-1), urate, urea, total protein, alanine aminotransferase (ALT), albumin (ALB), alkaline phosphatase (ALP), aspartate aminotransferase (AST), c-reactive protein (CRP), calcium (Ca), creatinine, cystatin C, gamma glutamyltransferase (GGT), phosphate, sex hormone binding globulin (SHBG), total bilirubin, and vitamin D.

For non-demographic variables, age at trait measurement was provided as an additional input variable with the measurement (Figure 1). Continuous input variables were scaled to zero mean and unit standard deviation using statistics from the train-val dataset, and discrete input variables were represented with one-hot encodings. Body composition variables were computed by automated segmentation of MRI instances by a previously published deep learning system that was used to produce publicly available UK Biobank body composition variables [34–37]. In brief, UNet [38] based deep learning models were trained to segment body composition tissues from UKBiobank neck-to-knee Dixon acquisitions using manual annotations. The UKBiobank protocol utilized the Siemens Aera 1.5T scanner (Syngo MR D13) and is detailed in [33].

**Fig. 1:**
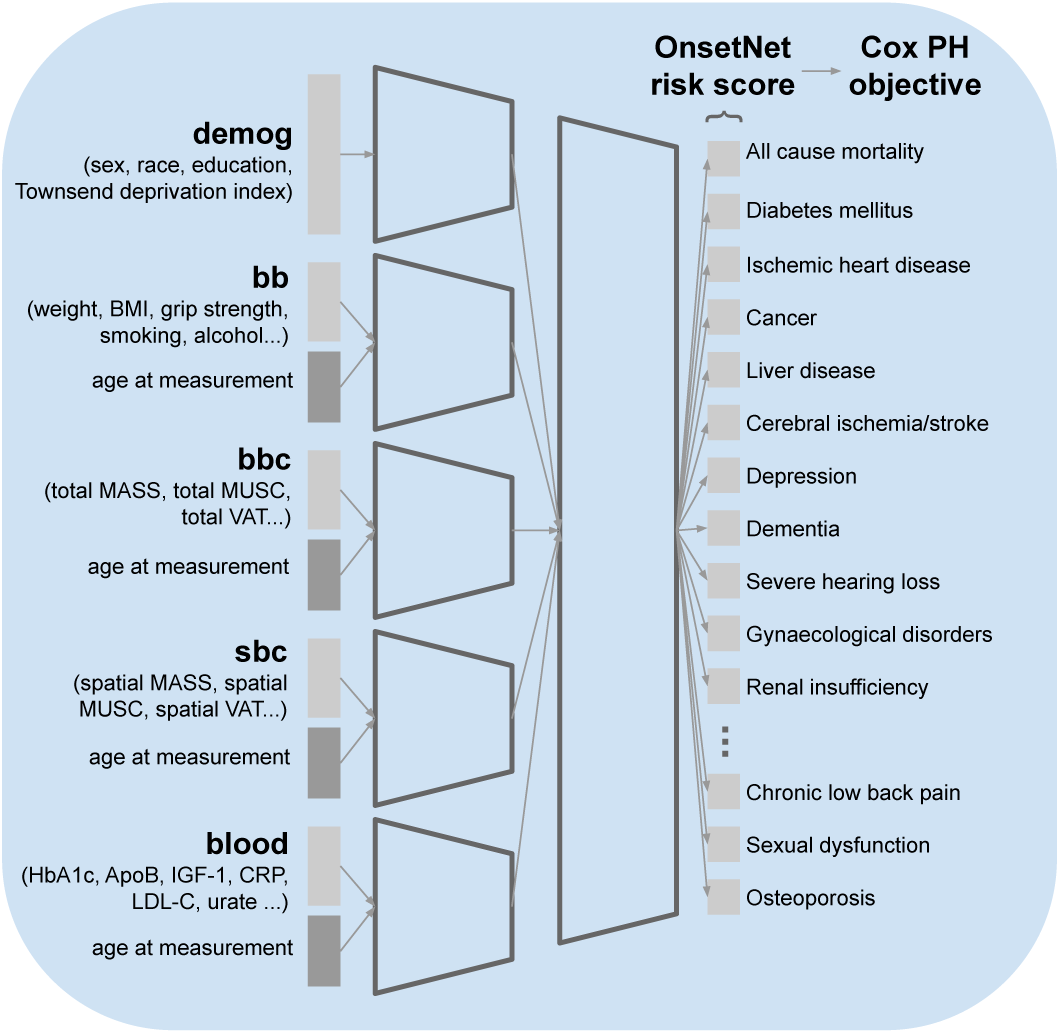
Architecture of OnsetNet, a neural network Cox model trained to predict onset acceleration as log-risk of first incidence of 47 disease conditions from individual traits, using birth as temporal baseline. Abbreviations: body mass index (BMI), body mass (MASS), muscle (MUSC), visceral adipose tissue (VAT), glycated haemoglobin (HbA1c), apolipoprotein B (ApoB), insulin-like growth factor 1 (IGF-1), c-reactive protein (CRP), low-density lipoprotein cholesterol (LDL-C).

Outcomes for 47 age-related conditions were chosen using literature on aging and multimorbidity [39, 40]. We obtained date of first occurrence for 45 non-composite disease categories defined by ICD10 codes (Appendix A.4) from the UK Biobank first occurrence register, which incorporates primary care, hospital inpatient, death register and self-reported records. Date of death was taken from the death register and date for all-cause morbidity was taken as first occurrence date for any of the 45 non-composite disease categories.

### 2.3 Deep learning estimation of onset acceleration risk

Neural network models trained to predict onset acceleration risk consisted of separate encoders per trait group each producing a feature vector of fixed length *k* ∈ {128, 256, 512}, followed by feature concatenation and a shared predictor head with one scalar output for each disease condition (Figure 1). Neural networks had an average of 2.31*E*6 ± 1.00*E*6 parameters (min 8.54E5, max 4.92E6, Appendix B).

Fivefold cross-validation was used to train models on the train-val dataset by optimizing objective *l*(*θ*), the Cox proportional hazard (Cox PH) partial log-likelihood objective function with weight decay regularization [41] over 47 disease conditions *d* ∈ D:

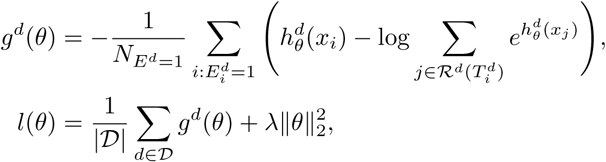

where *θ* are model parameters, *λ* is the weight decay coefficient, *x_i_* are input traits for individual *i*, log-risk 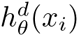 is the scalar output of the model for disease condition *d* and individual *i*, *E^d^* is the censoring vector for *d* where 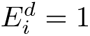 (uncensored) if individual with index *i* received a diagnosed for *d* and 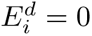 (censored) otherwise. 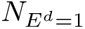 is the number of uncensored individuals for *d*. Birth was used as temporal baseline and time to event was measured in years (Figure 2). 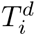 is age at first diagnosis for individual *i* with a diagnosis of *d* and otherwise age at censor date (November 30*^th^* 2022), risk set 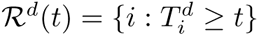 is the set of individuals undiagnosed with *d* at age *t*.

**Fig. 2:**
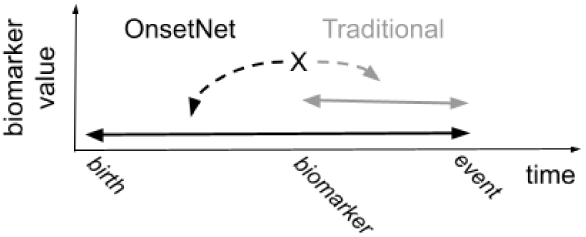
Cox models commonly predict risk of event earliness from measurement baseline while OnsetNet models event earliness from birth.

To evaluate performance, test sets were unseen during model selection of neural network Cox models and linear Cox comparison baselines. Hyperparameter values (*λ*, *k*, learning rate, dropout rate) for each input trait group combination were chosen by average validation fold C-index. Computation of C-index was batched with batch size of 2048 for tractability [27]. Neural networks were implemented in Python with GPU-accelerated PyTorch 2.4.1 and linear models in R 4.1.2. The code will be made available upon publication.

### 2.4 Statistical analysis

To investigate onset acceleration risk correlation between conditions, Pearson’s *r* was computed between OnsetNet score quartiles *q* ∈ [1, 4] for all pairs of conditions with internal test C-index performance ≥ 0.6. Correlation of *r* ≥ 0.6 was considered strong and *p* ≤ 3.46*E* − 5 was considered significant. Using the all-pairs correlation matrix, a clustering dendrogram was computed using unweighted pair group method with arithmetic mean (UPGMA) hierarchical agglomerative clustering for the linkage function (SciPy *cluster.hierarchy.linkage*) with cosine distance between correlation vectors as the clustering metric. Statin users (12,157 out of 60,396 in the study cohort reporting statin medication usage in verbal interview) were removed, and sensitivity analyses were conducted for removal of Bonferroni correction and inclusion of statin users.

To evaluate the prognostic ability of onset acceleration scores, a prognostic Cox model was trained for each pair of disease conditions (A, B), with OnsetNet risk quartile for condition A as input and individual log-risk of time-to-event for first occurrence of condition B as output. Baseline time was taken as imaging assessment time, corresponding to the latest date across input trait measurements, outcome events were within 10 years of baseline, individuals with pre-existing diagnoses were excluded, and time-to-event was measured in months. Demographic traits were included as input covariates for primary adjustment of hazard ratios, and secondary adjustment for both demographic and basic body traits was also performed to test for sensitivity. 6 outcome conditions with greater than 75% allocation to either sex (female: gynecological problems, thyroid diseases, osteoporosis; male: prostatic hyperplasia, hyperuricemia/gout, sexual dysfunction) were trained for the dominant sex only. Outcome events for 3 conditions with fewer than 50 uncensored individuals after removal of pre-imaging diagnoses were removed (hemorrhoids, Parkinson’s disease, and somatoform disorders).

Two-tailed tests were used to compute *p*-values and Bonferroni corrections were made at *p* = 0.05 level. The study adhered to the EQUATOR TRIPOD+ AI [42] guidelines for transparent reporting of an AI multivariable prediction model and STROBE [43] guidelines for reports of observational studies. Completed checklists are given in Appendix D.

## 3 Results

### 3.1 Predictive performance

A summary of participant characteristics for training and test datasets is given in Table 1. Eight different combinations of trait group inputs were evaluated for the neural network OnsetNet architecture and compared to a linear architecture (Table 2). High-dimensional spatial body composition was utilized for neural network models but not linear models due to correlation in body composition along the height dimension. The OnsetNet full traits model (demog+bb+bbc+sbc+blood) attained the highest internal test C-index (0.6830 ± 9.02*E* − 2), highest combined internal and external test C-index (0.6647 ± 1.11*E* − 1), and highest validation C-index (0.6856 ± 9.08*E* − 1) on average over conditions and cross-validation data folds. On the external test set, OnsetNet demog+bb+bbc+blood (0.6472 ± 1.23*E* − 1) performed best on C-index, followed by OnsetNet full traits (0.6461 ± 1.26*E* − 1). While performance differences between top OnsetNet models and the linear model were not statistically significant by estimated *p*-value, larger differences between models were statistically significant (Table 2, Table C10), and the linear model was consistently outperformed by neural networks on all metrics. Within specific conditions, diabetes mellitus non-T1 (0.940 ± 3.06*E* − 3 for OnsetNet full traits) and severe vision reduction (0.550 ± 2.54*E* − 3 for OnsetNet full traits) performed best and worst on internal test respectively (Figure 3).

**Fig. 3:**
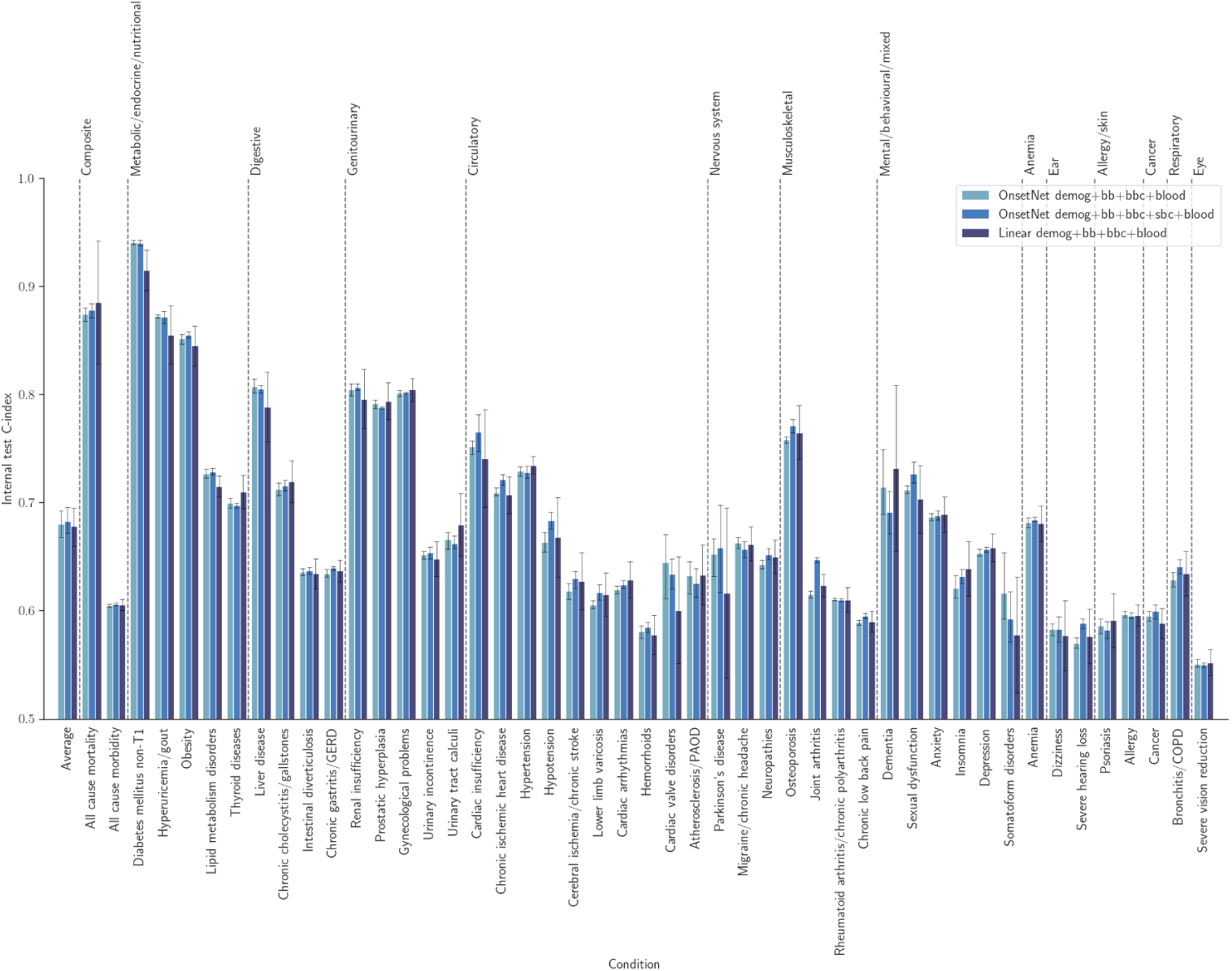
Internal test set C-index across disease conditions for linear and neural network architectures. Grey error bars illustrate 95% confidence intervals.

**Table 1:** Characteristics of UK Biobank participants used for the present study. *p*-values were computed with two-tailed t-test for continuous variables and *χ*^2^-test for categorical variables.

| Trait | Train-val<br>Value (ref.) | Internal test |  | External test |  |
| --- | --- | --- | --- | --- | --- |
| | | Value | $p$-value | Value | $p$-value |
| n | 50,610 | 8,931 | – | 855 | – |
| Age at baseline (yrs) | 54.89<br>$\pm 7.57$ | 54.74<br>$\pm 7.68$ | 9.80E-02 | 54.29<br>$\pm 7.53$ | 2.13E-02 |
| Sex |  |  | 9.18E-01 |  | 7.80E-01 |
| Female | 50.63 | 50.12 |  | 49.24 |  |
| Race |  |  | 9.99E-01 |  | 9.64E-01 |
| White | 96.66 | 96.93 |  | 97.31 |  |
| Education |  |  | 1.00E+00 |  | 9.99E-01 |
| College or University degree | 45.66 | 45.45 |  | 46.20 |  |
| Smoking |  |  | 9.99E-01 |  | 4.71E-01 |
| Never | 61.57 | 60.83 |  | 58.83 |  |
| Alcohol |  |  | 1.00E+00 |  | 9.67E-01 |
| Three or four times a week | 27.84 | 27.16 |  | 25.50 |  |
| BMI ($Kg/m^2$) | 26.55<br>$\pm 4.35$ | 26.61<br>$\pm 4.42$ | 2.20E-01 | 26.09<br>$\pm 4.38$ | 2.51E-03 |
| Grip strength ($Kg$) | 30.43<br>$\pm 10.61$ | 30.68<br>$\pm 10.65$ | 4.64E-02 | 30.23<br>$\pm 10.28$ | 5.76E-01 |
| HbA1c ($mmol/mol$) | 35.04<br>$\pm 5.09$ | 35.07<br>$\pm 5.16$ | 7.22E-01 | 34.62<br>$\pm 4.70$ | 1.55E-02 |
| HDL-C ($mmol/L$) | 1.47<br>$\pm 0.37$ | 1.47<br>$\pm 0.37$ | 6.20E-01 | 1.46<br>$\pm 0.36$ | 3.37E-01 |
| ApoB ($g/L$) | 1.01<br>$\pm 0.21$ | 1.01<br>$\pm 0.21$ | 7.74E-01 | 1.01<br>$\pm 0.22$ | 9.84E-01 |
| Hypertension (%) | 31.03 | 31.51 | 9.18E-01 | 32.98 | 6.74E-01 |
| Diabetes mellitus non-T1 (%) | 5.56 | 5.78 | 9.26E-01 | 4.80 | 7.37E-01 |
| Depression (%) | 11.03 | 11.29 | 9.34E-01 | 11.93 | 7.73E-01 |

**Table 2:** Test set performance across models measured by C-index. C-index values denote average over 47 conditions and 5 cross-valiation folds. *p*-values measure significance of difference with linear demog+bb+bbc+blood and were computed with two-tailed t-test. Bold font denotes top two methods for each metric.

| Method | Internal test<br>C-index | External test<br>C-index | All test<br>C-index |
| --- | --- | --- | --- |
| Linear |  |  |  |
| demog+bb+bbc+blood | $6.782E - 01$<br>$\pm 9.11E - 02$ | $6.435E - 01$<br>$\pm 1.50E - 01$ | $6.611E - 01$<br>$\pm 1.25E - 01$ |
| OnsetNet |  |  |  |
| demog | $5.863E - 01$<br>$\pm 6.96E - 02$<br>($p=2.73E-51$) | $5.683E - 01$<br>$\pm 1.06E - 01$<br>($p=8.05E-14$) | $5.774E - 01$<br>$\pm 9.01E - 02$<br>($p=1.84E-45$) |
| demog+bb | $6.404E - 01$<br>$\pm 7.66E - 02$<br>($p=4.55E-10$) | $6.187E - 01$<br>$\pm 1.35E - 01$<br>($p=1.41E-02$) | $6.297E - 01$<br>$\pm 1.10E - 01$<br>($p=1.16E-07$) |
| demog+bb+bbc | $6.649E - 01$<br>$\pm 7.81E - 02$<br>($p=2.74E-02$) | $6.304E - 01$<br>$\pm 1.21E - 01$<br>($p=1.93E-01$) | $6.478E - 01$<br>$\pm 1.03E - 01$<br>($p=2.50E-02$) |
| demog+bb+sbc | $6.699E - 01$<br>$\pm 7.88E - 02$<br>($p=1.69E-01$) | $6.309E - 01$<br>$\pm 1.23E - 01$<br>($p=2.10E-01$) | $6.506E - 01$<br>$\pm 1.05E - 01$<br>($p=7.68E-02$) |
| demog+bb+blood | $6.677E - 01$<br>$\pm 8.95E - 02$<br>($p=8.25E-02$) | $6.431E - 01$<br>$\pm 1.34E - 01$<br>($p=9.65E-01$) | $6.555E - 01$<br>$\pm 1.14E - 01$<br>($p=3.48E-01$) |
| demog+bb+bbc+blood | $6.799E - 01$<br>$\pm 9.06E - 02$<br>($p=7.84E-01$) | **$6.472E - 01$**<br>**$\pm 1.23E - 01$**<br>**($p=7.16E-01$)** | **$6.637E - 01$**<br>**$\pm 1.09E - 01$**<br>**($p=6.54E-01$)** |
| demog+bb+sbc+blood | **$6.820E - 01$**<br>**$\pm 9.02E - 02$**<br>**($p=5.41E-01$)** | $6.446E - 01$<br>$\pm 1.27E - 01$<br>($p=9.17E-01$) | $6.635E - 01$<br>$\pm 1.11E - 01$<br>($p=6.85E-01$) |
| demog+bb+bbc+sbc+blood | **$6.830E - 01$**<br>**$\pm 9.02E - 02$**<br>**($p=4.37E-01$)** | **$6.461E - 01$**<br>**$\pm 1.26E - 01$**<br>**($p=7.99E-01$)** | **$6.647E - 01$**<br>**$\pm 1.11E - 01$**<br>**($p=5.36E-01$)** |

We analysed the importance of adding individual trait groups to model inputs (Table C10). The largest performance gains on internal test C-index from adding traits to model inputs were adding bb to demog (Δ0.0542, *p* = 9.68*E* − 15), sbc to demog+bb (Δ0.0295, *p* = 4.81*E* − 5), and blood to demog+bb (Δ0.0272, *p* = 4.48*E* − 4). On internal and external test sets, utilizing body composition traits increased performance and was approximately twice as important in the absence of blood biomarkers. On internal test set C-index, adding bbc to demog+bb increased performance by Δ0.0244 (*p* = 6.85*E* − 4), sbc to demog+bb by Δ0.0295 (*p* = 4.81*E* − 5), bbc to demog+bb+blood by Δ0.0122 (*p* = 1.42*E* − 1), and sbc to demog+bb+blood by Δ0.0143 (*p* = 8.59*E* − 2). For models with blood traits included, small gains were observed on internal test C-index from utilizing increasingly complex body composition inputs (demog+bb+bbc+blood: 0.6799 ± 9.08*E* − 2, demog+bb+sbc+blood: 0.6820 ± 9.04*E* − 2, demog+bb+bbc+sbc+blood: 0.6830 ± 9.04*E* − 2), but performance on the external test set was highest for basic body composition (demog+bb+bbc+blood: 0.6472 ± 1.233*E* − 1, demog+bb+sbc+blood: 0.6446 ± 1.269*E* − 1, demog+bb+bbc+sbc+blood: 0.6461 ± 1.264*E* − 1). C-index was lower (Δ − 0.0306 ± 0.0080) with higher variance on the external test set compared to the internal test set for all input trait combinations.

Following model selection and performance evaluation, the full dataset was utilized for post-hoc analysis. The best performing neural network by both average validation C-index and test C-index was demog+bb+bbc+sbc+blood which provided OnsetNet scores for analysis. Onset acceleration score quartiles were computed by discretizing OnsetNet scores using sex and condition specific quartile bins computed on the train-val dataset. Kaplan-Meier estimates for onset acceleration score quartile illustrated within-condition heterogeneity (Figure 4).

**Fig. 4:**
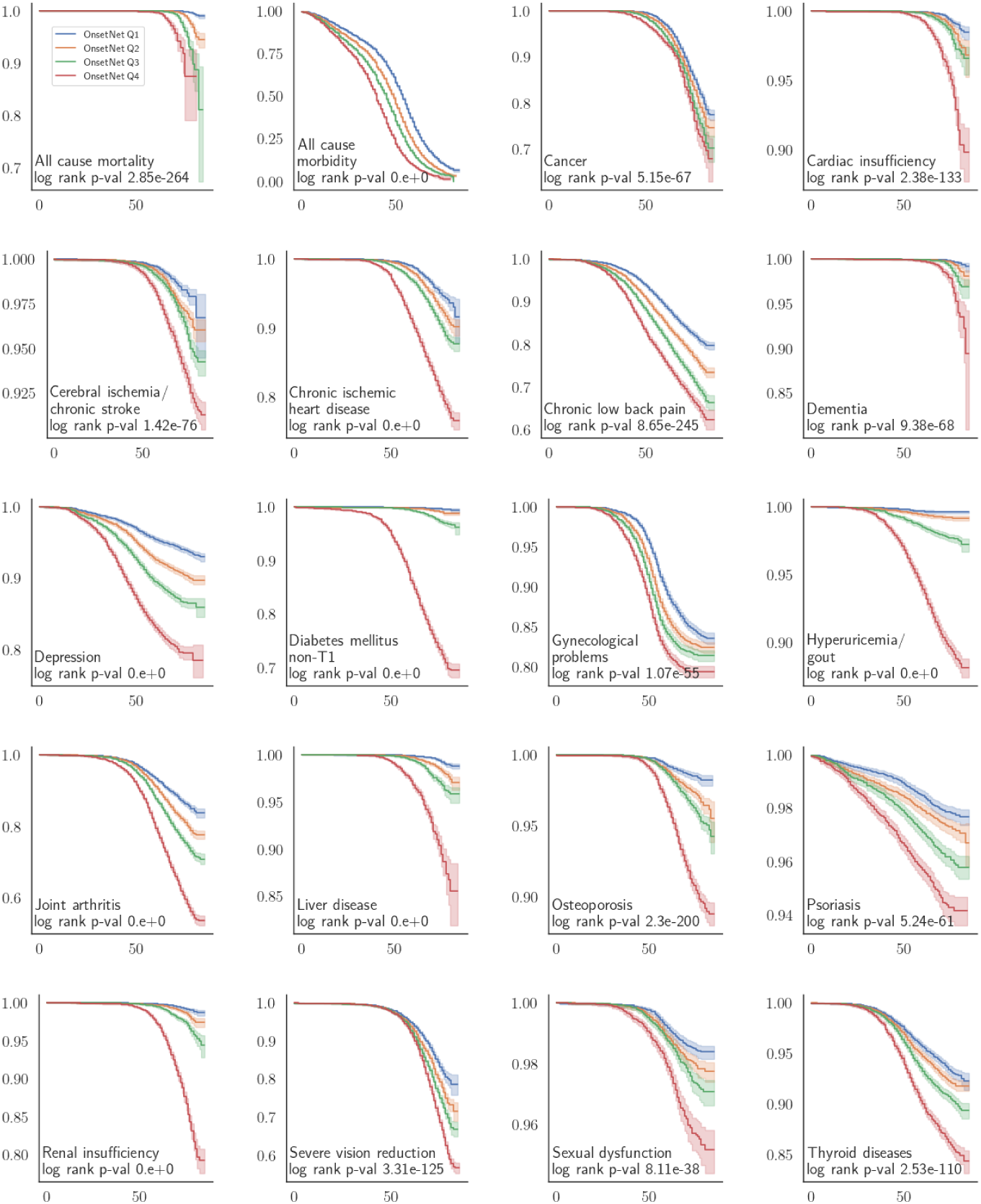
Kaplan-Meier survival curves of age in years (horizontal axis) against survival probability (vertical axis) for OnsetNet risk quartiles. All conditions and at-risk counts are given in Appendix C.

### 3.2 Inter-disease onset acceleration analysis

Pearson’s *r* was computed between OnsetNet score quartiles for each pair of 38 conditions where both conditions had internal test C-index performance ≥ 0.6 (Figure 5, Figure 6). 1420 out of 1444 correlations were significant at *p* ≤ 3.46*E* −5 level (Bonferroni corrected, *p* = 5.87*E* − 9 ± 9.81*E* − 8) including all strong correlations (*r* ≥ 0.6). Disease clusters and strong correlations were reproduced in sensitivity analysis without Bonferroni correction (*p* ≤ 0.05) or with inclusion of statin users (Appendix C).

**Fig. 5:**
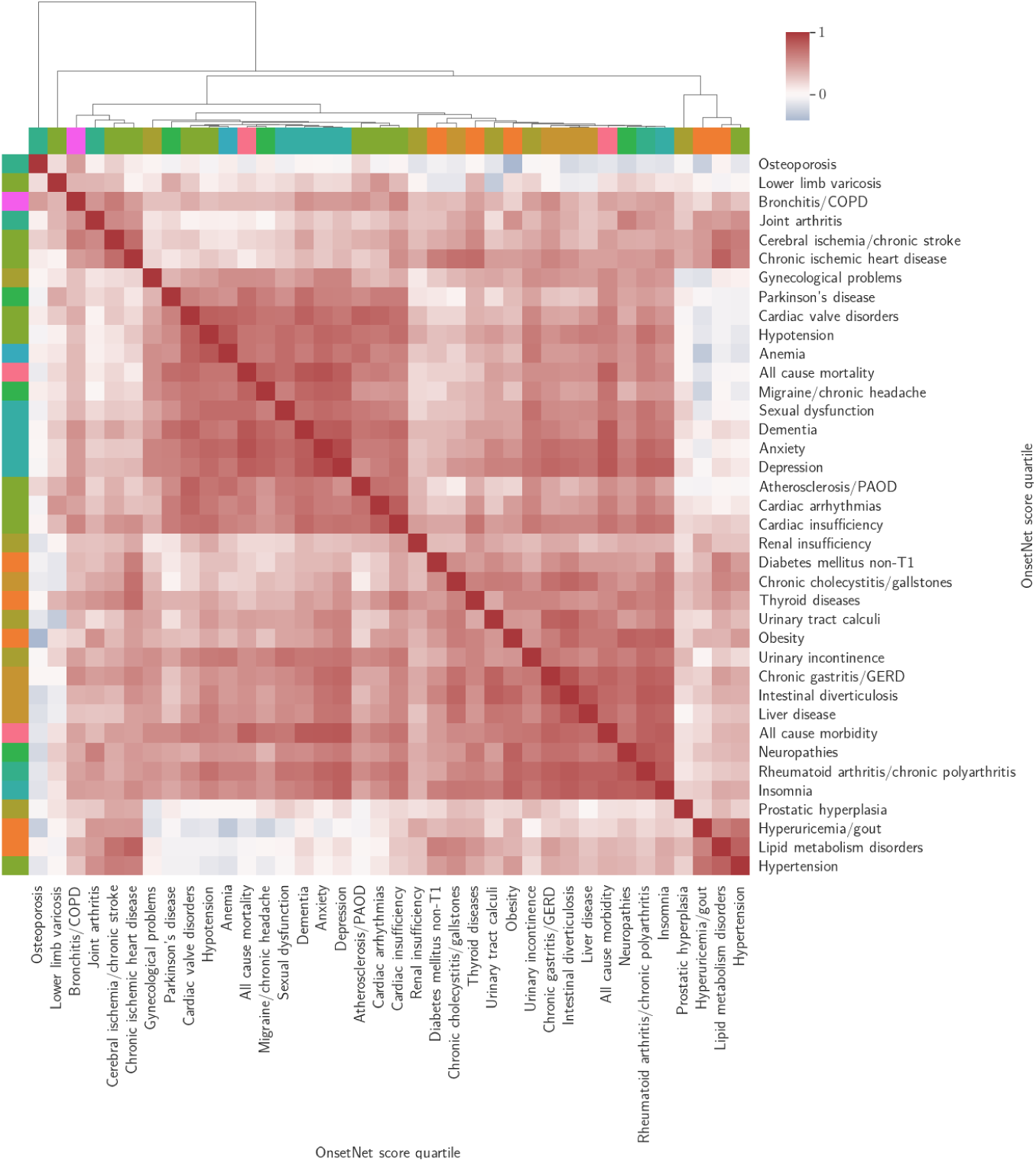
Clusterplot illustrating structure in the onset acceleration risk of age-associated diseases. Onset acceleration risk quartile correlations for conditions with internal test C-index ≥ 0.6 were clustered by hierarchical agglomerative clustering with cosine distance between correlation vectors as the clustering metric. Color denotes risk quartile correlation strength. Statin users were removed and 24 out of 1444 entries with *p >* 3.46*E* − 5 (Bonferroni corrected) were set to 0 for cosine distance. Row and column colours indicate condition types.

**Fig. 6:**
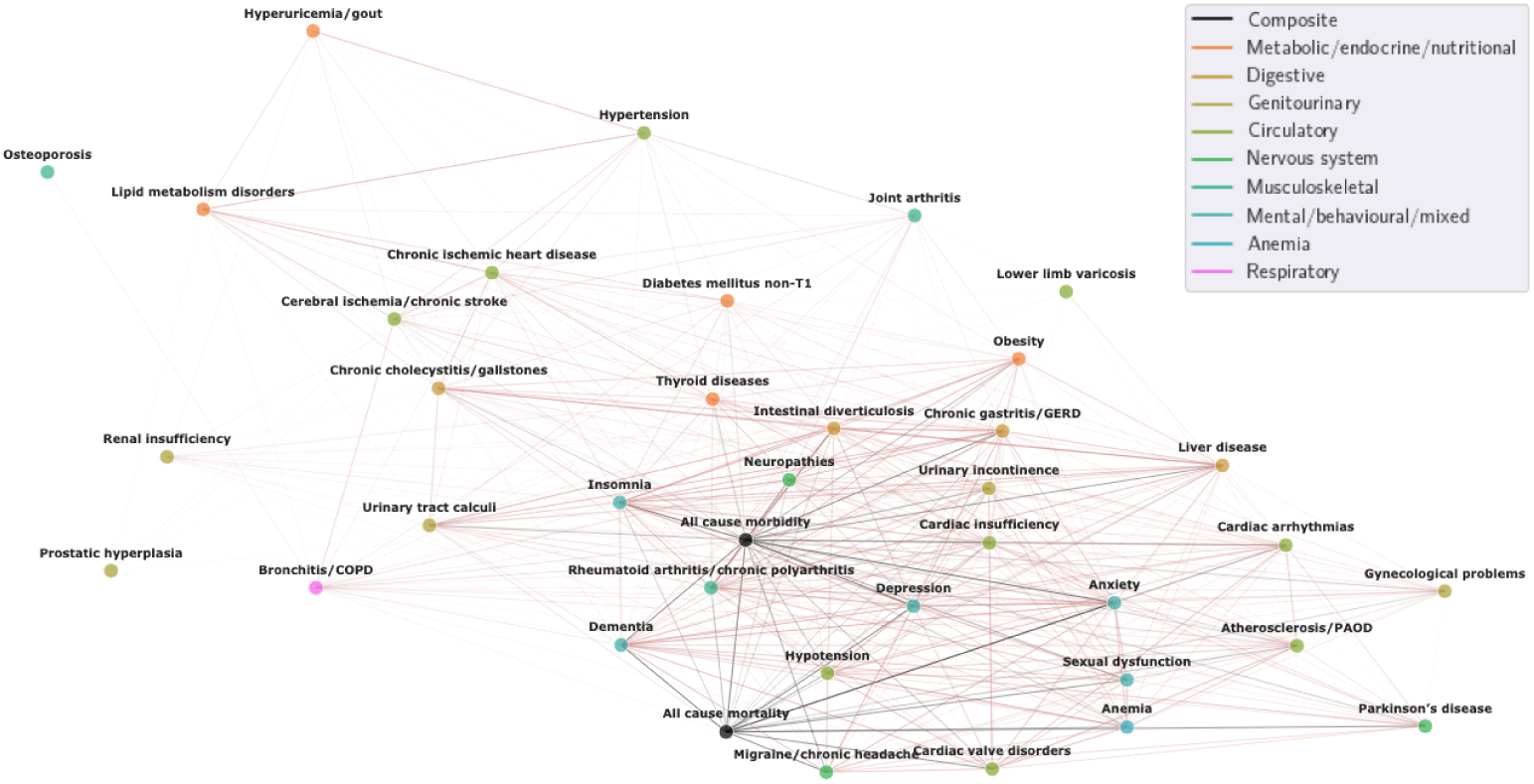
Graph of inter-disease OnsetNet risk quartile correlations for conditions with internal test C-index ≥ 0.6 and correlations with *r* ≥ 0.3 and *p* ≤ 3.46*E*−5 (Bonferroni corrected). Statin users were removed and *r* scales with edge width.

Onset acceleration risk quartile for all cause mortality and all cause morbidity were strongly correlated with each other (*r* = 0.78) and jointly strongly correlated with anxiety (*r* = 0.86, 0.82), dementia (*r* = 0.84, 0.82), depression (*r* = 0.80, 0.83), migraine/chronic headache (*r* = 0.77, 0.69), cardiac valve disorders (*r* = 0.75, 0.65), sexual dysfunction (*r* = 0.73, 0.67), hypotension (*r* = 0.69, 0.63), cardiac arrhythmias (*r* = 0.64, 0.63), and rheumatoid arthritis/chronic polyarthritis (*r* = 0.63, 0.79). Insomnia (*r* = 0.81), chronic gastritis/GERD (*r* = 0.78), intestinal diverticulosis (*r* = 0.75), liver disease (*r* = 0.72), cardiac insufficiency (*r* = 0.69), neuropathies (*r* = 0.68), urinary incontinence (*r* = 0.68), obesity (*r* = 0.67), and urinary tract calculi (*r* = 0.63) were strongly correlated with all cause morbidity but not with all cause mortality. Correlation between onset acceleration risk in obesity and all cause mortality was *r* = 0.43. Conversely, all cause mortality was strongly correlated with Parkinson’s disease (*r* = 0.72), anemia (*r* = 0.68), and atherosclerosis/PAOD (*r* = 0.62), which were not strongly correlated with all cause morbidity. The two strongest onset acceleration correlations with Parkinson’s disease were dementia (*r* = 0.73) and all cause mortality (*r* = 0.72).

Within non-composite conditions, onset acceleration risk clusters were identified by cosine distance between correlation vectors. A cardiometabolic cluster (corners of Figure 5) was identified where lipid metabolism disorders were associated with chronic ischemic heart disease (*r* = 0.79), hypertension (*r* = 0.79), cerebral ischemia/chronic stroke (*r* = 0.70), diabetes mellitus non-T1 (*r* = 0.63), hyperuricemia/gout (*r* = 0.61), and chronic cholecystitis/gallstones (*r* = 0.60) in a cluster that did not include obesity (*r* = 0.35).

Second, chronic gastritis/GERD was associated with a mixed digestive and neuropsychiatric cluster (lower right, Figure 5) containing intestinal diverticulosis (*r* = 0.85), rheumatoid arthritis/chronic polyarthritis (*r* = 0.79), all cause morbidity (*r* = 0.78), urinary tract calculi (*r* = 0.77), insomnia (*r* = 0.77), depression (*r* = 0.75), anxiety (*r* = 0.73), liver disease (*r* = 0.73), neuropathies (*r* = 0.71), chronic cholecystitis/gallstones (*r* = 0.71), urinary incontinence (*r* = 0.66), obesity (*r* = 0.63), dementia (*r* = 0.63), chronic ischemic heart disease (*r* = 0.60), cardiac insufficiency (*r* = 0.60), and thyroid diseases (*r* = 0.60).

Third, a vascular-neuropsychiatric cluster was identified (upper left, Figure 5), where onset acceleration risk in cardiac insufficiency was strongly associated with cardiac arrhythmias (*r* = 0.80), dementia (*r* = 0.75), hypotension (*r* = 0.73), rheumatoid arthritis/chronic polyarthritis (*r* = 0.71), cardiac valve disorders (*r* = 0.71), depression (*r* = 0.70), all cause morbidity (*r* = 0.69), atherosclerosis/PAOD (*r* = 0.69), insomnia (*r* = 0.68), thyroid diseases (*r* = 0.68), anxiety (*r* = 0.68), urinary incontinence (*r* = 0.67), parkinson’s disease (*r* = 0.64), anemia (*r* = 0.62), neuropathies (*r* = 0.61), chronic gastritis/GERD (*r* = 0.60), and bronchitis/COPD (*r* = 0.60).

Notable associations within the vascular-neuropsychiatric cluster included anxiety and depression (*r* = 0.9) which demonstrated the strongest correlation in onset acceleration risk overall, with top shared correlations for all cause mortality (*r* = 0.86, 0.80), all cause morbidity (*r* = 0.82, 0.83), dementia (*r* = 0.81, 0.81), migraine/chronic headache (*r* = 0.78, 0.77), rheumatoid arthritis/chronic polyarthritis (*r* = 0.77, 0.79), hypotension (*r* = 0.76, 0.73), sexual dysfunction (*r* = 0.74, 0.71), anemia (*r* = 0.74, 0.67), chronic gastritis/GERD (*r* = 0.73, 0.75), cardiac valve disorders (*r* = 0.71, 0.67), insomnia (*r* = 0.71, 0.79), intestinal diverticulosis (*r* = 0.69, 0.73), liver disease (*r* = 0.69, 0.69), cardiac insufficiency (*r* = 0.68, 0.70), urinary incontinence (*r* = 0.67, 0.69), cardiac arrhythmias (*r* = 0.64, 0.63), neuropathies (*r* = 0.63, 0.66), parkinson’s disease (*r* = 0.62, 0.62), gynecological problems (*r* = 0.61, 0.60), and urinary tract calculi (*r* = 0.60, 0.62). The strongest association with anemia was hypotension and vice versa (*r* = 0.84).

### 3.3 Prognostic survival analysis

Adjusted hazard ratios (*aHR*) for OnsetNet Q4 (Q1 as reference) for 47 input conditions and 44 outcome event conditions were rendered (Figure 7). 1334 out of 2068 (64.5%) hazard ratios were significant at *p* = 0.05 level after primary adjustment without Bonferroni correction, of which Schoenfeld residuals were insignificant (*p >* 0.05) for 1238 or 92.8% (*aHR* = 3.67 ± 5.78), and 1037 or 77.7% remained statistically significant after secondary adjustment. 435 out of 2068 (21.0%) hazard ratios were significant at *p* = 9.05*E* − 7 level (Bonferroni corrected *p* = 0.05) after primary adjustment (*aHR* = 6.11±9.00), where all Schoenfeld residuals were insignificant (*p >* 0.05), and 184 (42.3%) remained statistically significant after secondary adjustment. With Bonferroni correction, 25 out of 44 (56.8%) input conditions yielded statistically significant Q4 hazard ratios for the same outcome condition (*aHR* = 11.09 ± 17.8), and 27 statistically significant Q4 hazard ratios were observed for all cause mortality (*aHR* = 6.26 ± 9.33) of which 23 were strong with at least threefold increase in mortality risk compared to Q1 (*aHR* ≥ 3, *aHR* = 6.85 ± 9.98). Using binarized OnsetNet score instead of quartiles (above median vs. below median, Appendix C) yielded 23 statistically significant adjusted hazard ratios for all cause mortality with Bonferroni correction (*aHR* = 2.24 ± 0.408) of which only 2 were strong (all cause mortality and dementia, *aHR* = 3.39 ± 0.222), suggesting that onset acceleration risk Q4 was disproportionately associated with mortality hazard compared to lower quartiles.

**Fig. 7:**
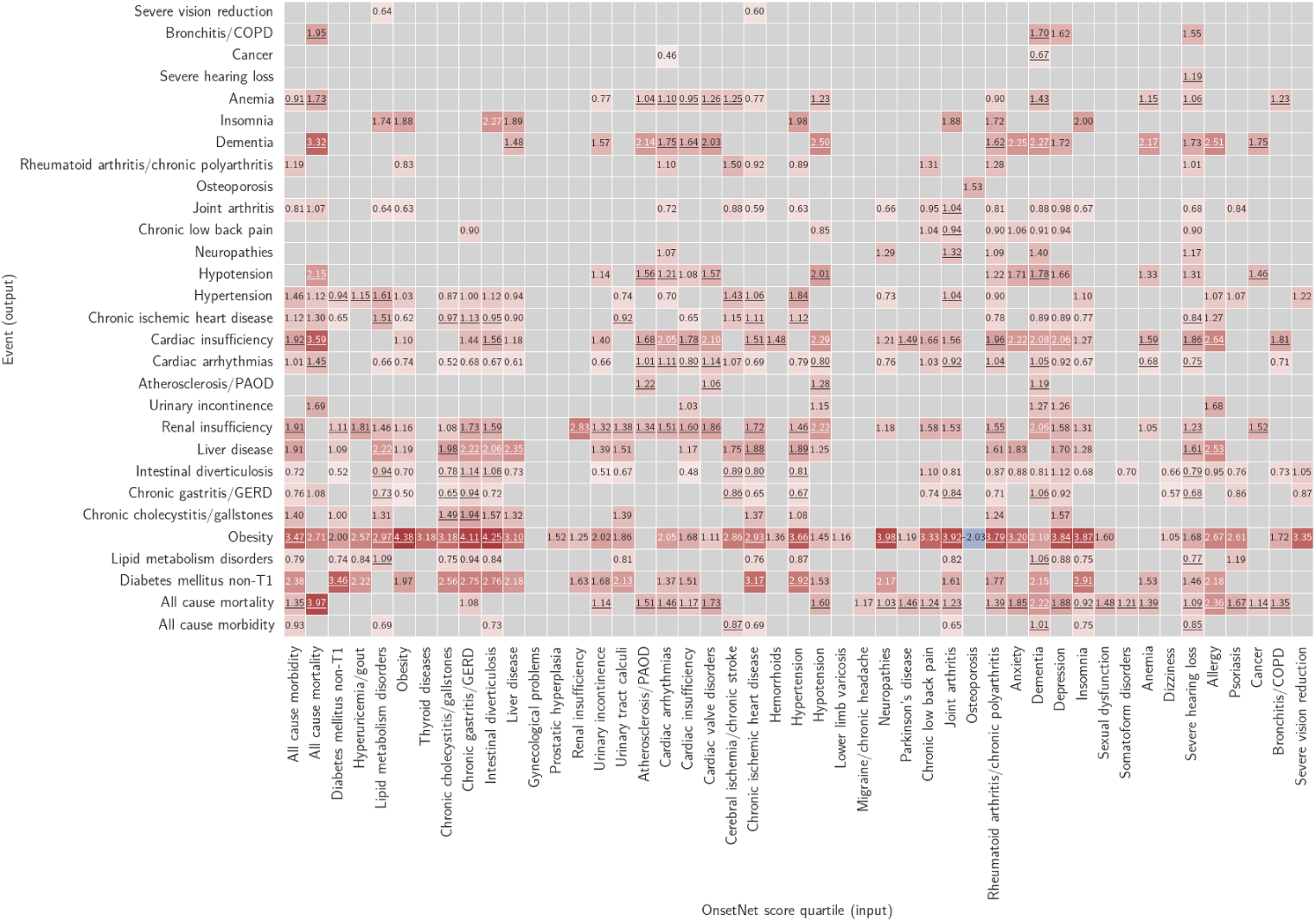
Adjusted log hazard ratios for OnsetNet risk Q4 (Q1 as reference) given primary adjustment with demographic variables. Color red (blue) indicates positive (negative) log hazard ratios and color intensity scales with log hazard ratio magnitude. Gray cells denote log hazard ratios that were statistically insignificant following primary adjustment (*p >* 9.05*E* − 7, Bonferroni corrected) or where Schoenfeld residual *p* ≤ 0.05. Entries that remained statistically significant after secondary adjustment with basic body traits are underlined to denote robustness to change in adjustment.

### 3.4 Biomarker saliency

To investigate the relationship learned by the neural network between input traits and onset acceleration risk of conditions, we computed sex-specific normalized absolute partial derivative of network input variables with respect to each output [44, 45]. Univariate traits (Figure 8) and multivariate spatial body composition (Figure 9) were normalized separately.

**Fig. 8:**
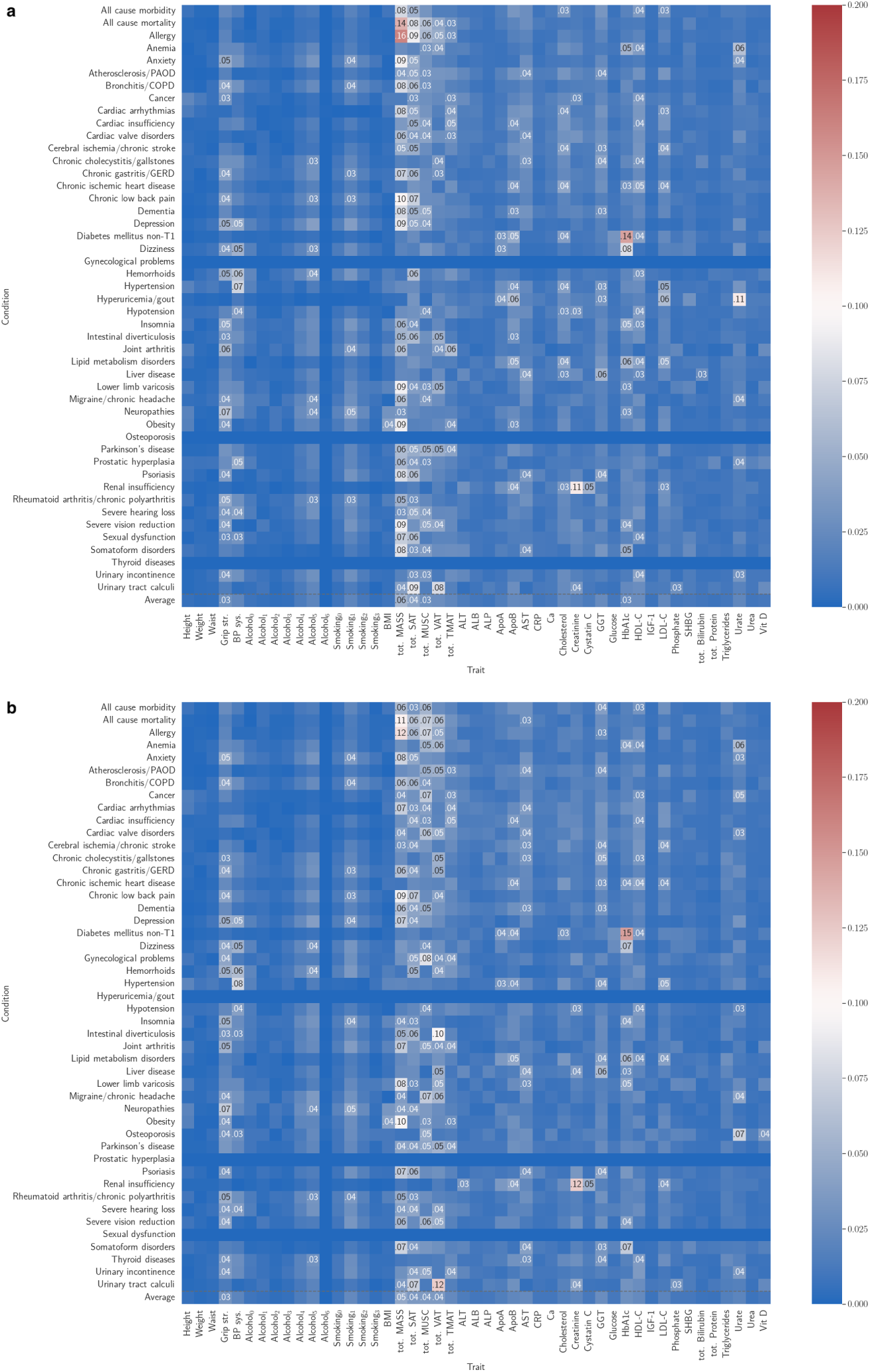
Saliency map for univariate traits for (a) male and (b) female participants. Top 5 salient traits per condition are labelled. Average saliency across conditions given in the bottom row.

**Fig. 9:**
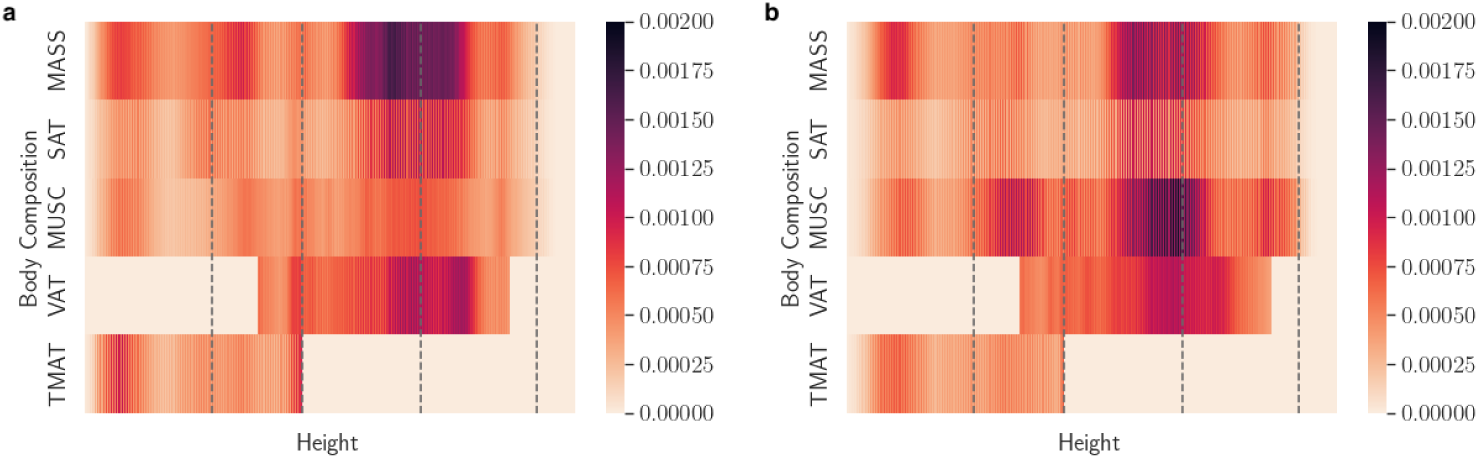
Spatial body composition saliency maps averaged over 47 conditions for (a) male and (b) female participants. Condition-specific maps are given in Appendix C.5.

Known relationships inferred by saliency without a-priori knowledge include creatinine and cystatin C for renal insufficiency, HbA1c for diabetes mellitus non-T1, urate for hyperuricemia/gout, GGT for liver disease, total MASS for obesity, systolic blood pressure for hypertension and hypotension. Notably, BMI and total MASS were not in the top 5 traits for onset acceleration risk of diabetes mellitus non-T1, which was dominated by HbA1c. The 10 most important individual traits on average over conditions for the full traits OnsetNet model were total MASS, total SAT, total MUSC, total VAT, grip strength, GGT, HbA1c, HDL-C for both males and females, ApoB and cholesterol for males, and AST and total TMAT for females. Additional traits in the top 10 for the full univariate traits model included smoking, total TMAT, and urate for males, and alcohol, smoking and urate for females (Appendix C). The most salient spatial body composition regions were VAT and MASS located at the abdomen for males, and muscle and MASS around the abdomen for females on average over conditions. Less overt relationships inferred for both sexes included HBA1c, HDL-C and urate for anemia; GGT, ApoB, AST and body composition for dementia; urate for osteoporosis; total MASS, HBA1c and grip strength for insomnia. Total MASS was highly salient for chronic low back pain, and total TMAT was highly salient for cardiac conditions (arrhythmias, insufficiency, valve disorders). For both sexes, body composition traits were more salient than blood biomarkers for predicting onset acceleration of dementia and Parkinson’s disease; the top five univariate biomarkers for Parkinson’s disease were basic body composition traits. ALT was salient for liver disease but not in the top 5 traits, which featured AST and GGT.

## 4 Discussion

For the methodological contribution of the study, we validated that onset acceleration risk of diverse age-associated diseases can be modelled using neural network and linear Cox architectures, with neural networks outperforming linear models on in-distribution and out-of-distribution test data. While there was high variability in predictive performance across conditions, the final model selected by cross-validation utilized all input traits and attained internal test C-index ≥ 0.6 on 38 out of 47 conditions (80.9%). Model applicability was not limited by temporal differences in trait measurement times (11.3 ± 3.03 years between blood samples and imaging scans for individuals in this study) since the relevant period is lifespan instead of post-measurement time. Inclusion of body composition was found to be important for predictive performance across disease conditions, with performance gain from adding body composition metrics to basic demographic and anthropomorphic traits being comparable to the performance gain from adding blood biomarkers.

In post-hoc analysis, a number of connections between diseases with existing support in the literature were inferred without a-priori knowledge, such as associations in onset correlation risk between Parkinson’s disease, dementia and all cause mortality, cardiovascular and cerebrovascular conditions [16, 39, 46]. Strong correlations in onset acceleration risk were inferred for cardiometabolic, digestive-neuropsychiatric, and vascular-neuropsychiatric condition groups, partially corroborating a previous review of cross-sectional multimorbidity studies across different populations which identified cardiometabolic, neuropsychiatric-gastrointestinal, and musculoskeletal-vascular-neuropsychiatric-gastrointestinal co-occurrence clusters [15] despite the use of different methodology that associated diseases by co-occurrence. Automated inference of associations between biomarkers and disease identified pre-supported associations in a precise manner, including biomarkers for renal insufficiency, diabetes and liver disease, adding to previous work that used partial derivative based saliency to identify biologically salient factors for disease [45]. Saliency analysis validated the importance of grip strength and body composition traits [47].

This study is subject to several limitations. Firstly, existing work showed that the UK Biobank cohort contains a healthy participant bias and is predominantly composed of individuals from White ethnic backgrounds [48]. Although models adjusted for sociodemographic factors, conclusions drawn may not hold for more representative or ethnically diverse populations. All conditions except all cause mortality were reliant on the timing and definition of diagnoses, which depends on factors such as access to healthcare services and diagnostic criteria. Underestimation of disease prevalence in the UK Biobank is known to occur secondary to underdiagnosis [49, 50]. While Cox models are the dominant survival analysis approach in medical research and clinical trials [51, 52] and have been used for time-to-death analysis of aging clocks [22], it is an approximate model of survival as the proportional hazards assumption generally does not hold perfectly. In this exploratory analysis, results from evaluation against ground truth event times (Table 2, Figures 3, 4, 7) indicated that the Cox PH objective was capable of training models to predict onset acceleration risk, but alternative models may also be applied in future work. Additionally, models were assessed on held-out in-distribution and out-of-distribution test sets but are yet to be evaluated on an entirely independent cohort, and direct clinical utility and explicit causal relationships were not assessed. Prospective and intervention based studies are required to show clinical utility and support the validity of associations inferred in this study.

In summary, we introduced and evaluated the use of Cox models for modelling onset acceleration risk in the context of multimorbid aging, and performed analysis on inter-disease relationships, within-disease heterogeneity across individuals, and biomarker saliency for onset acceleration risk prediction. This work opens up novel approaches for the analysis of age-related diseases and associated biomarker predictor traits, with potential clinical applicability to early identification of high-risk groups in the population.

## Declarations

## Data Availability

Participant data from the UK Biobank cohort was obtained through UK Biobank Access Application number 44584. The UK Biobank has approval from the North West Multi-center Research Ethics Committee (REC reference: 11/NW/0382). All methods were performed in accordance with the relevant guidelines and regulations, and informed consent was obtained from all participants. Researchers may apply to use the UK Biobank data resource by submitting a health-related research proposal that is in the public interest. More information may be found on the UK Biobank researchers and resource catalogue pages (www.ukbiobank.ac.uk).

## Acknowledgements

The authors would like to thank the UK Biobank Reading team, Madeleine Cule, Ramprakash Srinivasan, Adam Kosiorek, and Martin Tobin for their support.

## Author statement

M. X. Ji conducted literature search, study design, data curation and verification, model training and data analysis, results interpretation, figures, writing first draft, and writing review and edits. M. Thanaj conducted literature search, study design, data curation and verification, results interpretation, and writing review and edits. L. Néhale-Ezzine, B. Whitcher, E. L. Thomas, and J. D. Bell conducted results interpretation, and writing review and edits.

## Data access statement

Participant data from the UK Biobank cohort was obtained through UK Biobank Access Application number 44584. The UK Biobank has approval from the North West Multi-center Research Ethics Committee (REC reference: 11/NW/0382). All methods were performed in accordance with the relevant guidelines and regulations, and informed consent was obtained from all participants. Researchers may apply to use the UK Biobank data resource by submitting a health-related research proposal that is in the public interest. More information may be found on the UK Biobank researchers and resource catalogue pages (www.ukbiobank.ac.uk). No participant or public involvement occurred during the design, conduct, reporting, interpretation, or dissemination of this study.

## Appendix A Dataset

Dataset details for *n* = 60, 396 individuals are provided, including demographics, outcome counts, and biomarker descriptions.

### A.1 Demographics

**Table A1:** Alcohol intake frequency.

| Sex | Alcohol intake frequency | Percentage (%) | n |
| --- | --- | --- | --- |
| Male | Daily or almost daily | 22.83 | 6820 |
|  | Never | 5.03 | 1503 |
|  | Once or twice a week | 25.73 | 7685 |
|  | One to three times a month | 9.11 | 2722 |
|  | Prefer not to answer | 0.04 | 12 |
|  | Special occasions only | 6.44 | 1925 |
|  | Three or four times a week | 30.82 | 9206 |
|  | Total | 100.00 | 29873 |
| Female | Daily or almost daily | 15.05 | 4595 |
|  | Never | 7.22 | 2204 |
|  | Once or twice a week | 26.71 | 8152 |
|  | One to three times a month | 13.24 | 4042 |
|  | Prefer not to answer | 0.02 | 7 |
|  | Special occasions only | 13.10 | 3997 |
|  | Three or four times a week | 24.66 | 7526 |
|  | Total | 100.00 | 30523 |

**Table A2:** Smoking status.

| Sex | Smoking status | Percentage (%) | n |
| --- | --- | --- | --- |
| Male | Current | 4.96 | 1481 |
|  | Never | 57.70 | 17236 |
|  | Prefer not to answer | 0.30 | 89 |
|  | Previous | 37.05 | 11067 |
|  | Total | 100.00 | 29873 |
| Female | Current | 3.62 | 1105 |
|  | Never | 65.07 | 19860 |
|  | Prefer not to answer | 0.33 | 101 |
|  | Previous | 30.98 | 9457 |
|  | Total | 100.00 | 30523 |

**Table A3:** Race.

| **Sex** | **Race** | **Percentage (%)** | **n** |
| --- | --- | --- | --- |
| Male | Asian or Asian British | 1.54 | 461 |
|  | Black or Black British | 0.53 | 159 |
|  | Mixed | 0.32 | 95 |
|  | Other | 0.83 | 247 |
|  | White | 96.78 | 28911 |
|  | Total | 100.00 | 29873 |
| Female | Asian or Asian British | 1.22 | 371 |
|  | Black or Black British | 0.69 | 211 |
|  | Mixed | 0.63 | 193 |
|  | Other | 0.82 | 251 |
|  | White | 96.64 | 29497 |
|  | Total | 100.00 | 30523 |

**Table A4:** Education.

| **Sex** | **Education** | **Percentage (%)** | **n** |
| --- | --- | --- | --- |
| Male | A levels/AS levels or equivalent | 11.71 | 3499 |
|  | College or University degree | 47.23 | 14108 |
|  | CSEs or equivalent | 4.40 | 1314 |
|  | None of the above | 6.77 | 2022 |
|  | NVQ or HND or HNC or equivalent | 7.98 | 2383 |
|  | O levels/GCSEs or equivalent | 17.59 | 5254 |
|  | Other professional qualifications eg: nursing, teaching | 3.95 | 1179 |
|  | Prefer not to answer | 0.38 | 114 |
|  | Total | 100.00 | 29873 |
| Female | A levels/AS levels or equivalent | 14.45 | 4412 |
|  | College or University degree | 44.09 | 13457 |
|  | CSEs or equivalent | 4.25 | 1296 |
|  | None of the above | 6.21 | 1895 |
|  | NVQ or HND or HNC or equivalent | 3.28 | 1001 |
|  | O levels/GCSEs or equivalent | 21.82 | 6660 |
|  | Other professional qualifications eg: nursing, teaching | 5.62 | 1716 |
|  | Prefer not to answer | 0.28 | 86 |
|  | Total | 100.00 | 30523 |

**Table A5:** Continuous demographic traits.

|  | **Male** | **Female** |
| --- | --- | --- |
| Mean birth year | 1952.47 | 1953.92 |
| Std birth year | 7.65 | 7.42 |
| Min birth year | 1937.00 | 1937.00 |
| Max birth year | 1970.00 | 1970.00 |
| IQR birth year | 13.00 | 12.00 |
| Mean Townsend deprivation index | -1.95 | -1.86 |

**Table A6:**
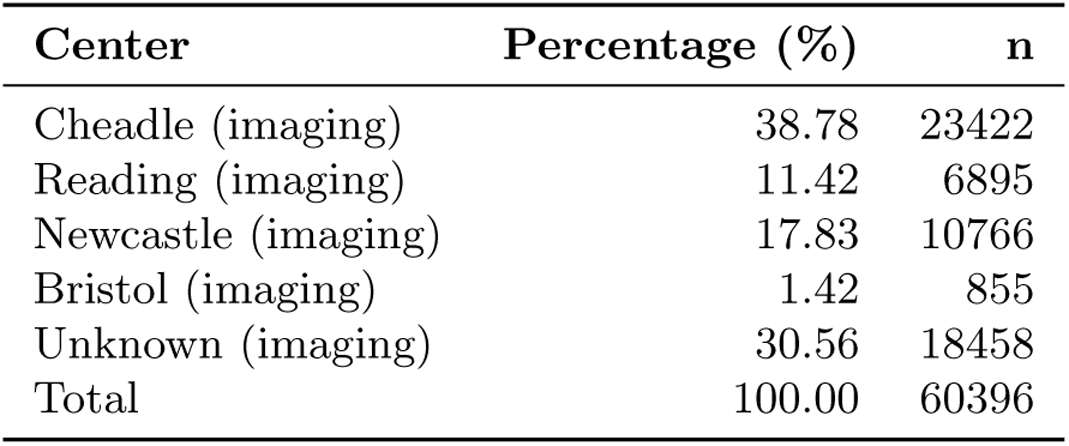
Assessment center.

| **Center** | **Percentage (%)** | **n** |
| --- | --- | --- |
| Cheadle (imaging) | 38.78 | 23422 |
| Reading (imaging) | 11.42 | 6895 |
| Newcastle (imaging) | 17.83 | 10766 |
| Bristol (imaging) | 1.42 | 855 |
| Unknown (imaging) | 30.56 | 18458 |
| Total | 100.00 | 60396 |

### A.2 Trait distributions across dataset

**Fig. A1:**
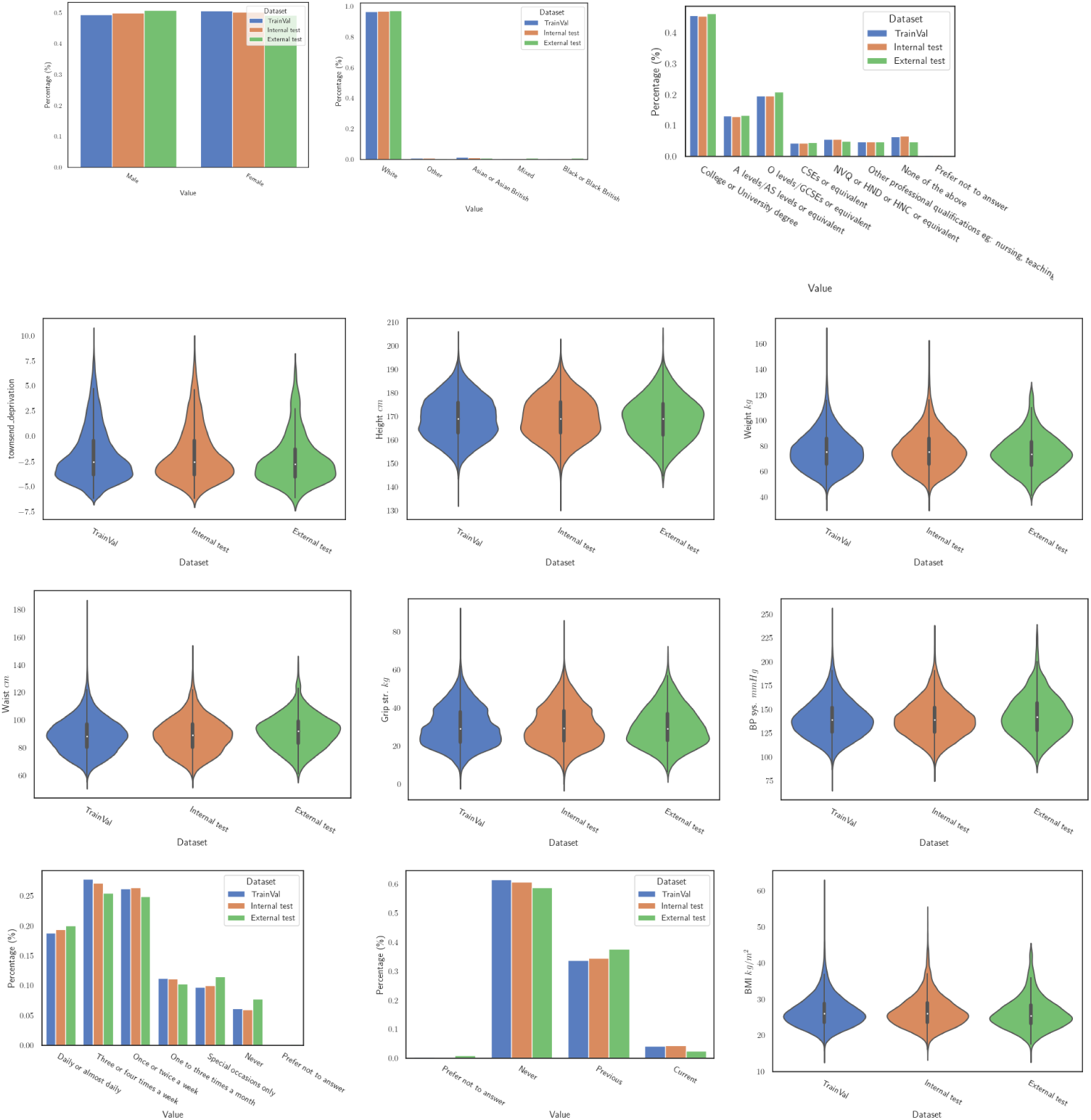
Demographic and basic body traits across dataset splits.

### A.3 Data censoring

The UK Biobank dataset used in this study contained participants of middle age and older (50.18 years old was mean age minus two standard deviations) who had medical records collected within a finite time window, therefore data was subject to left and right censoring. Cox PH survival analysis accounted for right censoring via relative risk adjustment; participants whose medical record window were not extended late enough to include future diagnoses were optimised to have higher risk scores than people with diagnoses within the window. Limitation imposed by left censoring was ameliorated by focusing on age-associated diseases which are more prevalent from middle age onwards. Finally, large dataset size assisted the model in learning robust associations inspite of limited window size.

### A.4 Disease conditions

**Table A7:** Non-composite condition definitions by ICD10 code.

| Condition group | Condition | ICD10 codes |
| --- | --- | --- |
| Metabolic/<br>endocrine/<br>nutritional | Diabetes mellitus non-T1 | E11, E12, E13, E14 |
|  | Hyperuricemia/ gout | E79, M10 |
|  | Obesity | E66 |
|  | Lipid metabolism disorders | E78 |
|  | Thyroid diseases | E01, E02, E03, E04, E05, E06, E07 |
| Genitourinary | Renal insufficiency | N18, N19 |
|  | Prostatic hyperplasia | N40 |
|  | Gynecological problems | N81, N84, N85, N86, N87, N88,<br>N89, N90, N93, N95 |
|  | Urinary incontinence | N39, R32 |

**Table A7 (continued)**
|  |  |  |
| --- | --- | --- |
|  | Urinary tract calculi | N20 |
| Circulatory | Cardiac insufficiency | I50 |
|  | Chronic ischemic heart disease | I20, I25, I21 |
|  | Hypertension | I10, I11, I12, I13, I14, I15 |
|  | Hypotension | I95 |
|  | Cerebral ischemia/ chronic stroke | I60, I61, I62, I63, I64, I69, G45 |
|  | Lower limb varicosis | I83, I87 |
|  | Cardiac arrhythmias | I44, I45, I46, I47, I48, I49 |
|  | Hemorrhoids | I84 |
|  | Cardiac valve disorders | I34, I37 |
|  | Atherosclerosis/ PAOD | I65, I66, I67, I70, I73 |
| Digestive | Liver disease | K70, K71, K72, K73, K74, K76 |
|  | Chronic cholecystitis/ gallstones | K80, K81 |
|  | Intestinal diverticulosis | K57 |
|  | Chronic gastritis/ GERD | K21, K25, K26, K27, K28, K29 |
| Mental/<br>behavioural/<br>mixed | Dementia | F00, F01, F02, F03, F05, G30, G31,<br>R54 |
|  | Sexual dysfunction | F52, N48 |
|  | Anxiety | F40, F41 |
|  | Insomnia | G47, F51 |
|  | Depression | F32, F33 |
|  | Somatoform disorders | F45 |
| Anemia | Anemia | D50, D51, D52, D53, D55, D56,<br>D57, D58, D59, D60, D61, D63, D64 |

**Table A7 (continued)**
|  |  |  |
| --- | --- | --- |
| Nervous system | Parkinson's disease | G20, G21, G22 |
|  | Migraine/ chronic headache | G43, G44 |
|  | Neuropathies | G50, G51, G52, G53, G54, G55,<br>G56, G57, G58, G59, G60, G61,<br>G62, G63, G64 |
| Respiratory | Bronchitis/ COPD | J40, J41, J42, J43, J44, J47 |
| Musculoskeletal | Osteoporosis | M80, M81, M82 |
|  | Joint arthritis | M15, M16, M17, M18, M19 |
|  | Rheumatoid arthritis/<br>chronic polyarthritis | M05, M06, M79 |
|  | Chronic low back pain | M40, M41, M42, M43, M44, M45,<br>M47, M48, M50, M51, M52, M53,<br>M54 |
| Allergy/ skin | Psoriasis | L40 |
|  | Allergy | H01, J30, L23, L27, L56, K52, K90,<br>T78, T88 |
| Eye | Severe vision reduction | H17, H18, H25, H26, H27, H28,<br>H31, H33, H34, H35, H36, H40,<br>H43, H47, H54 |
| Ear | Dizziness | H81, H82, R42 |
|  | Severe hearing loss | H90, H91 |

**Table A7 (continued)**
|  |  |  |
| --- | --- | --- |
| Cancer | Cancer | C00, C01, C02, C03, C04, C05, C06, C07, C08, C09, C10, C11, C12, C13, C14, C15, C16, C17, C18, C19, C20, C21, C22, C23, C24, C25, C26, C30, C31, C32, C33, C34, C35, C36, C37, C38, C39, C40, C41, C43, C44, C45, C46, C47, C48, C49, C50, C51, C52, C53, C54, C55, C56, C57, C58, C60, C61, C62, C63, C64, C65, C66, C67, C68, C69, C70, C71, C72, C73, C74, C75, C76, C77, C78, C79, C80, C81, C82, C83, C84, C85, C86, C87, C88, C89, C90, C91, C92, C93, C94, C95, C96, C97, D00, D01, D02, D03, D04, D05, D06, D07, D08, D09, D37, D38, D39, D40, D41, D42, D43, D44, D45, D46, D47, D48 |

**Table A8:** Condition event counts.

| Condition | Male | Female |
| --- | --- | --- |
| All cause morbidity | 26335 | 27318 |
| All cause mortality | 277 | 146 |
| Allergy | 5835 | 6380 |
| Anemia | 1559 | 3020 |
| Anxiety | 1571 | 2728 |
| Atherosclerosis/PAOD | 796 | 807 |
| Bronchitis/COPD | 1571 | 1441 |

**Table A8 (continued)**
|  |  |  |
| --- | --- | --- |
| Cancer | 3865 | 3570 |
| Cardiac arrhythmias | 3122 | 1758 |
| Cardiac insufficiency | 508 | 185 |
| Cardiac valve disorders | 317 | 219 |
| Cerebral ischemia/chronic stroke | 1130 | 676 |
| Chronic cholecystitis/gallstones | 1185 | 2177 |
| Chronic gastritis/GERD | 6472 | 6220 |
| Chronic ischemic heart disease | 3220 | 1253 |
| Chronic low back pain | 6352 | 6903 |
| Dementia | 169 | 107 |
| Depression | 2497 | 4194 |
| Diabetes mellitus non-T1 | 2215 | 1158 |
| Dizziness | 475 | 791 |
| Gynecological problems | 4 | 10193 |
| Hemorrhoids | 1925 | 1672 |
| Hypertension | 11184 | 7618 |
| Hyperuricemia/gout | 1725 | 173 |
| Hypotension | 554 | 467 |
| Insomnia | 1135 | 775 |
| Intestinal diverticulosis | 3676 | 3367 |
| Joint arthritis | 5267 | 6513 |
| Lipid metabolism disorders | 8400 | 4681 |
| Liver disease | 642 | 556 |
| Lower limb varicosis | 1124 | 1756 |
| Migraine/chronic headache | 1422 | 3249 |
| Neuropathies | 2217 | 2954 |
| Obesity | 1956 | 1823 |
| Osteoporosis | 318 | 1825 |

**Table A8 (continued)**
|  |  |  |
| --- | --- | --- |
| Parkinson’s disease | 96 | 46 |
| Prostatic hyperplasia | 4425 | 2 |
| Psoriasis | 1015 | 908 |
| Renal insufficiency | 997 | 815 |
| Rheumatoid arthritis/chronic polyarthritis | 4145 | 5153 |
| Severe hearing loss | 1294 | 865 |
| Severe vision reduction | 5048 | 5203 |
| Sexual dysfunction | 1063 | 105 |
| Somatoform disorders | 123 | 207 |
| Thyroid diseases | 1058 | 3907 |
| Urinary incontinence | 1529 | 3475 |
| Urinary tract calculi | 1016 | 418 |

### A.5 Quartile definitions

**Table A9:** OnsetNet score quartile boundaries computed for the best model (demog+bb+bbc+sbc+blood) from the train-val dataset.

| Condition | Male |  |  | Female |  |  |
| --- | --- | --- | --- | --- | --- | --- |
|  | Q1 end | Q2 end | Q3 end | Q1 end | Q2 end | Q3 end |
| All cause morbidity | 0.403 | 0.629 | 0.869 | 0.531 | 0.761 | 1.016 |
| All cause mortality | -0.289 | 1.610 | 3.678 | -0.772 | 0.963 | 2.914 |
| Allergy | 0.471 | 0.693 | 0.943 | 0.502 | 0.678 | 0.888 |
| Anemia | -1.593 | -1.263 | -0.895 | -1.077 | -0.527 | 0.102 |
| Anxiety | -0.037 | 0.415 | 0.905 | 0.582 | 1.009 | 1.523 |
| Atherosclerosis/PAOD | 0.139 | 0.511 | 0.903 | 0.121 | 0.533 | 0.958 |
| Bronchitis/COPD | 0.390 | 0.750 | 1.136 | 0.402 | 0.721 | 1.048 |
| Cancer | -0.881 | -0.788 | -0.684 | -1.011 | -0.762 | -0.373 |

Table A9 (continued)
|  |  |  |  |  |  |  |
| --- | --- | --- | --- | --- | --- | --- |
| Cardiac arrhythmias | -0.156 | 0.134 | 0.442 | -0.674 | -0.390 | -0.065 |
| Cardiac insufficiency | 0.337 | 0.918 | 1.529 | -0.601 | -0.043 | 0.580 |
| Cardiac valve disorders | -1.962 | -1.542 | -1.102 | -2.396 | -1.856 | -1.254 |
| Cerebral ischemia/chronic stroke | 1.351 | 1.683 | 2.056 | 0.788 | 1.079 | 1.420 |
| Chronic cholecystitis/gall-stones | -1.013 | -0.588 | -0.183 | -0.539 | -0.069 | 0.505 |
| Chronic gastritis/GERD | 0.110 | 0.414 | 0.734 | -0.009 | 0.291 | 0.630 |
| Chronic ischemic heart disease | 0.446 | 0.885 | 1.399 | -0.674 | -0.242 | 0.281 |
| Chronic low back pain | -0.204 | 0.018 | 0.244 | -0.122 | 0.100 | 0.332 |
| Dementia | -1.546 | -0.749 | 0.086 | -1.812 | -1.049 | -0.223 |
| Depression | -1.068 | -0.657 | -0.220 | -0.473 | -0.098 | 0.321 |
| Diabetes mellitus non-T1 | -1.534 | -0.389 | 1.001 | -2.371 | -1.346 | -0.071 |
| Dizziness | -1.028 | -0.817 | -0.611 | -0.438 | -0.198 | 0.047 |
| Gynecological problems | -2.602 | -2.414 | -2.192 | 2.871 | 3.068 | 3.244 |
| Hemorrhoids | -0.109 | 0.071 | 0.287 | -0.405 | -0.199 | 0.049 |
| Hypertension | 2.360 | 2.897 | 3.462 | 1.762 | 2.289 | 2.892 |
| Hyperuricemia/gout | 1.538 | 2.436 | 3.370 | -1.022 | -0.314 | 0.569 |
| Hypotension | -2.655 | -2.119 | -1.492 | -3.003 | -2.327 | -1.525 |
| Insomnia | 0.593 | 1.034 | 1.574 | 0.349 | 0.654 | 1.042 |
| Intestinal diverticulosis | -0.645 | -0.328 | 0.035 | -0.769 | -0.478 | -0.138 |
| Joint arthritis | -0.260 | 0.026 | 0.338 | -0.043 | 0.272 | 0.622 |
| Lipid metabolism disorders | 1.175 | 1.696 | 2.277 | 0.407 | 0.931 | 1.495 |
| Liver disease | -2.166 | -1.368 | -0.404 | -2.284 | -1.524 | -0.610 |
| Lower limb varicosis | -0.014 | 0.257 | 0.519 | 0.562 | 0.820 | 1.080 |
| Migraine/chronic headache | -0.332 | -0.071 | 0.201 | 0.468 | 0.774 | 1.075 |
| Neuropathies | 0.229 | 0.518 | 0.848 | 0.485 | 0.773 | 1.102 |

**Table A9 (continued)**
|  |  |  |  |  |  |  |
| --- | --- | --- | --- | --- | --- | --- |
| Obesity | -0.204 | 0.776 | 1.834 | -0.821 | 0.193 | 1.460 |
| Osteoporosis | -1.091 | -0.697 | -0.260 | 0.695 | 1.149 | 1.619 |
| Parkinson's disease | 1.555 | 2.370 | 3.106 | 0.437 | 1.174 | 1.921 |
| Prostatic hyperplasia | 3.460 | 3.740 | 4.024 | -1.954 | -1.698 | -1.408 |
| Psoriasis | 1.084 | 1.387 | 1.668 | 0.769 | 1.079 | 1.401 |
| Renal insufficiency | -2.061 | -1.331 | -0.523 | -1.946 | -1.279 | -0.528 |
| Rheumatoid arthri-<br>tis/chronic polyarthritis | -0.549 | -0.311 | -0.061 | -0.359 | -0.098 | 0.196 |
| Severe hearing loss | -0.498 | -0.349 | -0.192 | -0.934 | -0.781 | -0.610 |
| Severe vision reduction | 1.281 | 1.457 | 1.609 | 1.390 | 1.551 | 1.715 |
| Sexual dysfunction | 1.134 | 1.380 | 1.652 | -1.461 | -1.139 | -0.658 |
| Somatoform disorders | 0.152 | 0.518 | 0.839 | 0.486 | 0.931 | 1.340 |
| Thyroid diseases | -0.640 | -0.421 | -0.162 | 0.708 | 0.931 | 1.191 |
| Urinary incontinence | -0.404 | -0.240 | -0.054 | 0.419 | 0.677 | 0.980 |
| Urinary tract calculi | -0.231 | 0.077 | 0.410 | -1.140 | -0.861 | -0.542 |

### A.6 Data pre-processing

The UK Biobank study protocol is publically available [25]. The inclusion flowchart is given in A2.

**Fig. A2:**
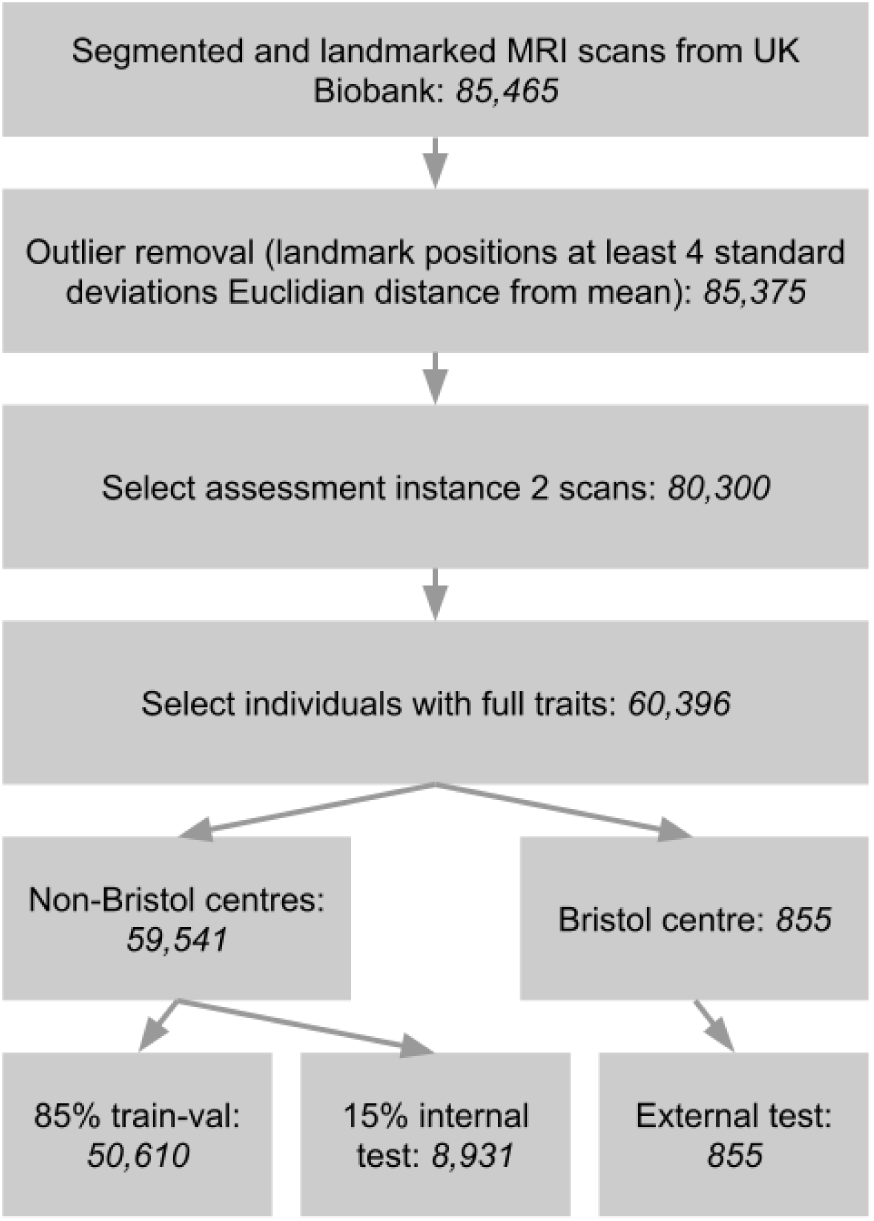
Pre-processing flowchart for data utilized in the study.

#### Body composition variables

The sbc variable with dimension [5 × 370] consisted of mass, subcutaneous adipose tissue, muscle, visceral adipose tissue, thigh intramuscular and intermuscular adipose tissue channels measured along 370 vertical slices, segmented and landmarked (at mid-thigh, hip, T12 vertebra as top of the iliopsoas muscle, and shoulder) with a previously published deep learning system [34–37]. Spatial body composition was computed from tissue segmentation maps by summing tissue volume for each vertical slice to yield a [1 × 370] vector per tissue type along the height dimension, followed by linear interpolation to register vectors to sex-specific average landmark positions and concatenation to combine vectors into a matrix with dimension [5 × 370].

## Appendix B Architecture

OnsetNet’s architecture is illustrated in Figure B3.

**Fig. B3:**
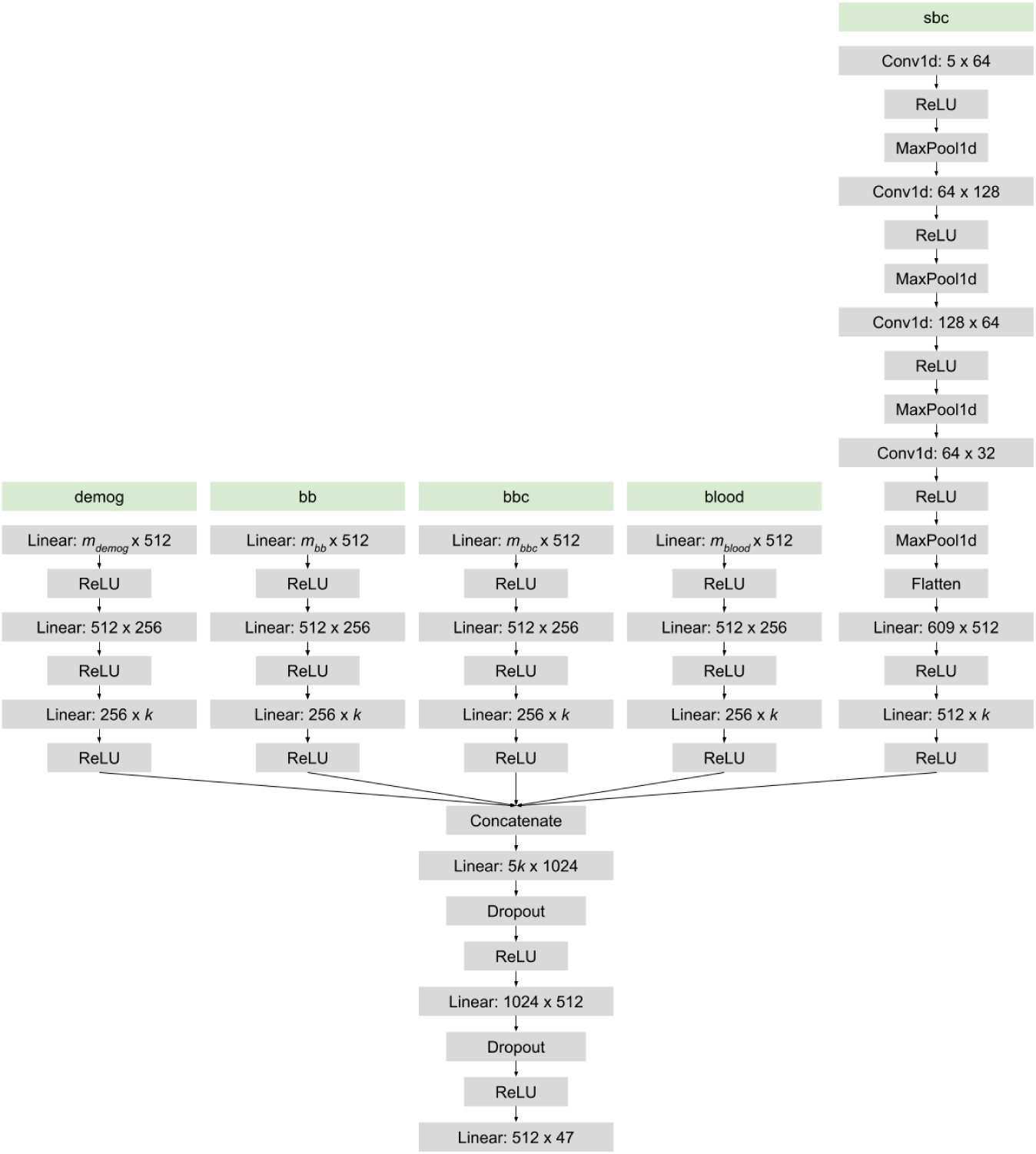
Architecture for neural network OnsetNet with channel sizes and PyTorch naming convention. Input variables shown in green. *m_g_* denotes input dimensions of trait group *g*. Encoders produce feature vectors of fixed length *k* ∈ {128, 256, 512}, where *k* is a hyperparameter.

## Appendix C Additional Results

### C.1 Addition of biomarker inputs

**Table C10:**
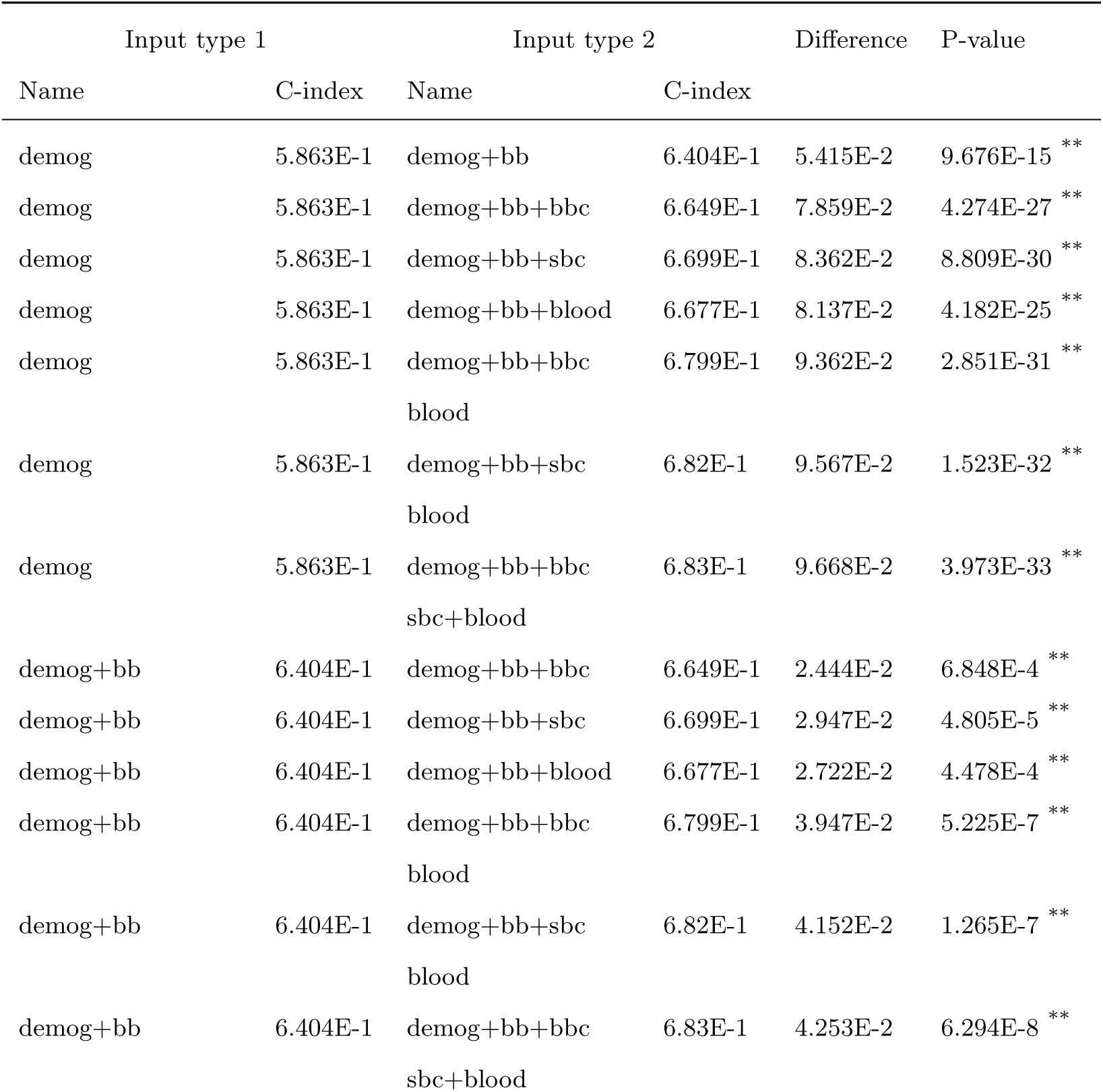
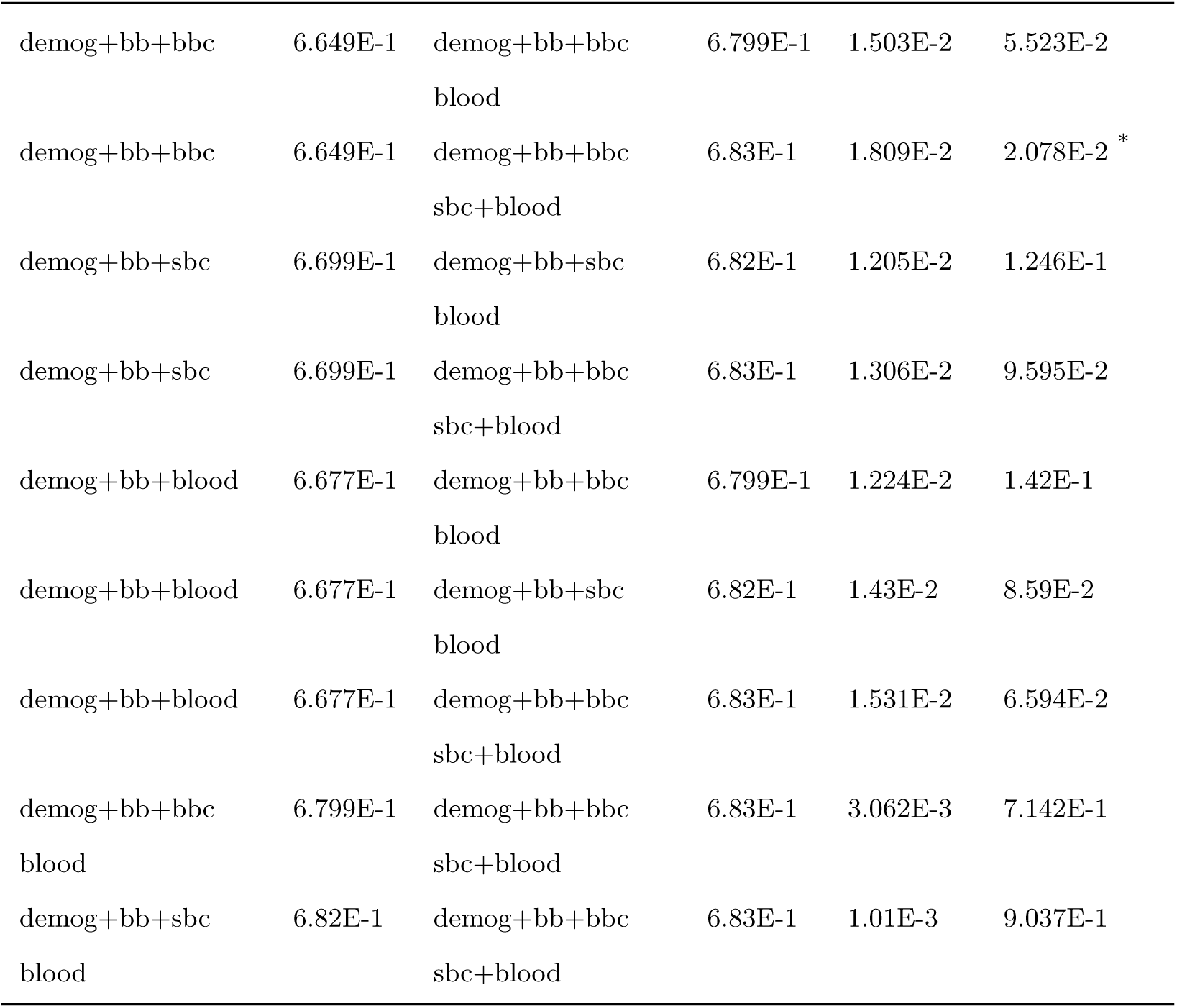
Difference in internal test C-index from adding additional input types to change from input type 1 to input type 2. C-index values denote average over 47 conditions and 5 folds. * (**) denotes significance at *p* = 0.05 (Bonferroni corrected *p* = 2.27*E* − 3) level.

### C.2 Test performance

**Fig. C4:**
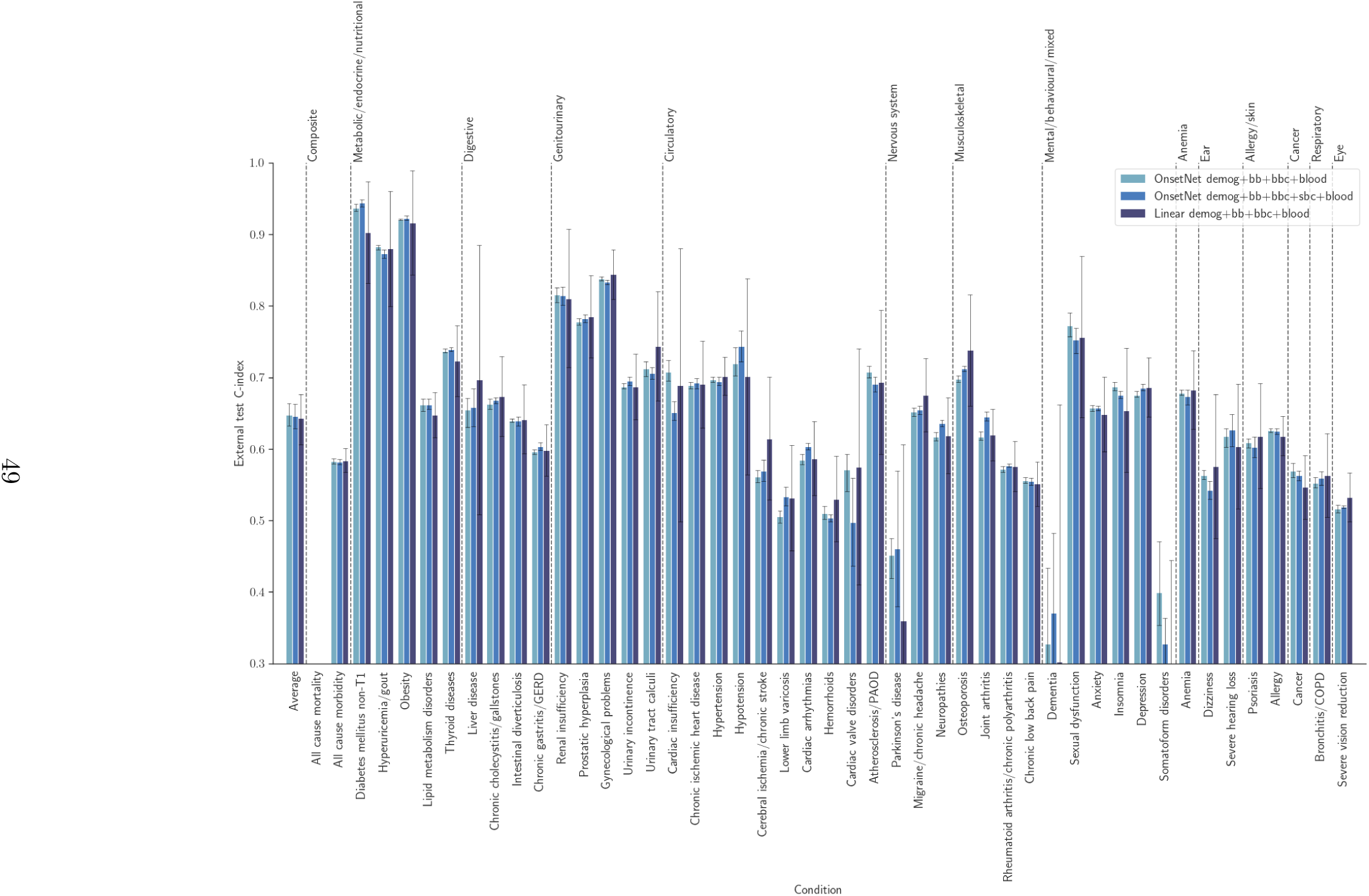
External test set C-index across disease conditions for neural network and linear architectures. Gray error bars illustrate 95% confidence intervals.

**Fig. C5:**
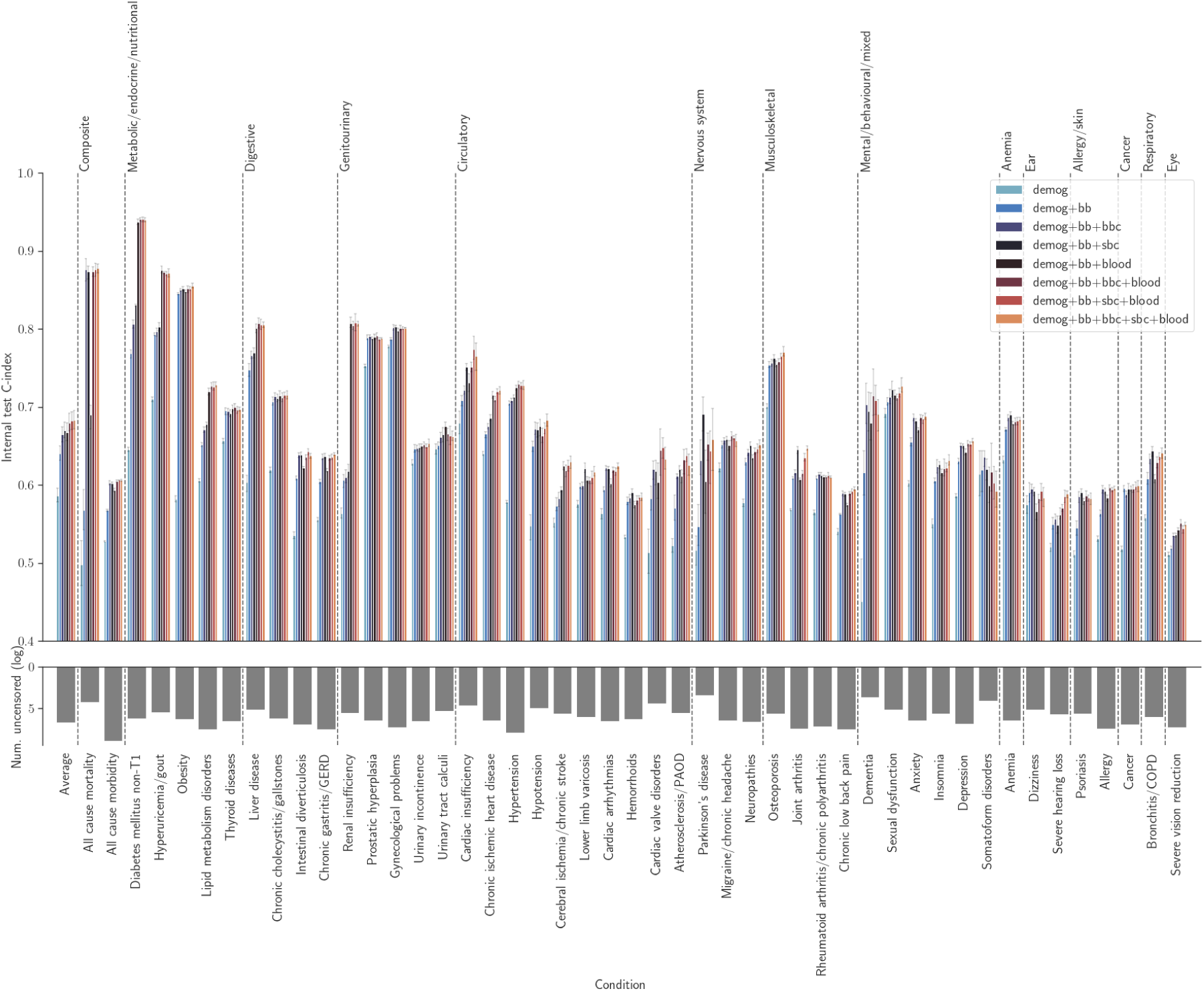
Internal test set C-index across disease conditions for neural network architectures. Gray error bars illustrate 95% confidence intervals.

**Fig. C6:**
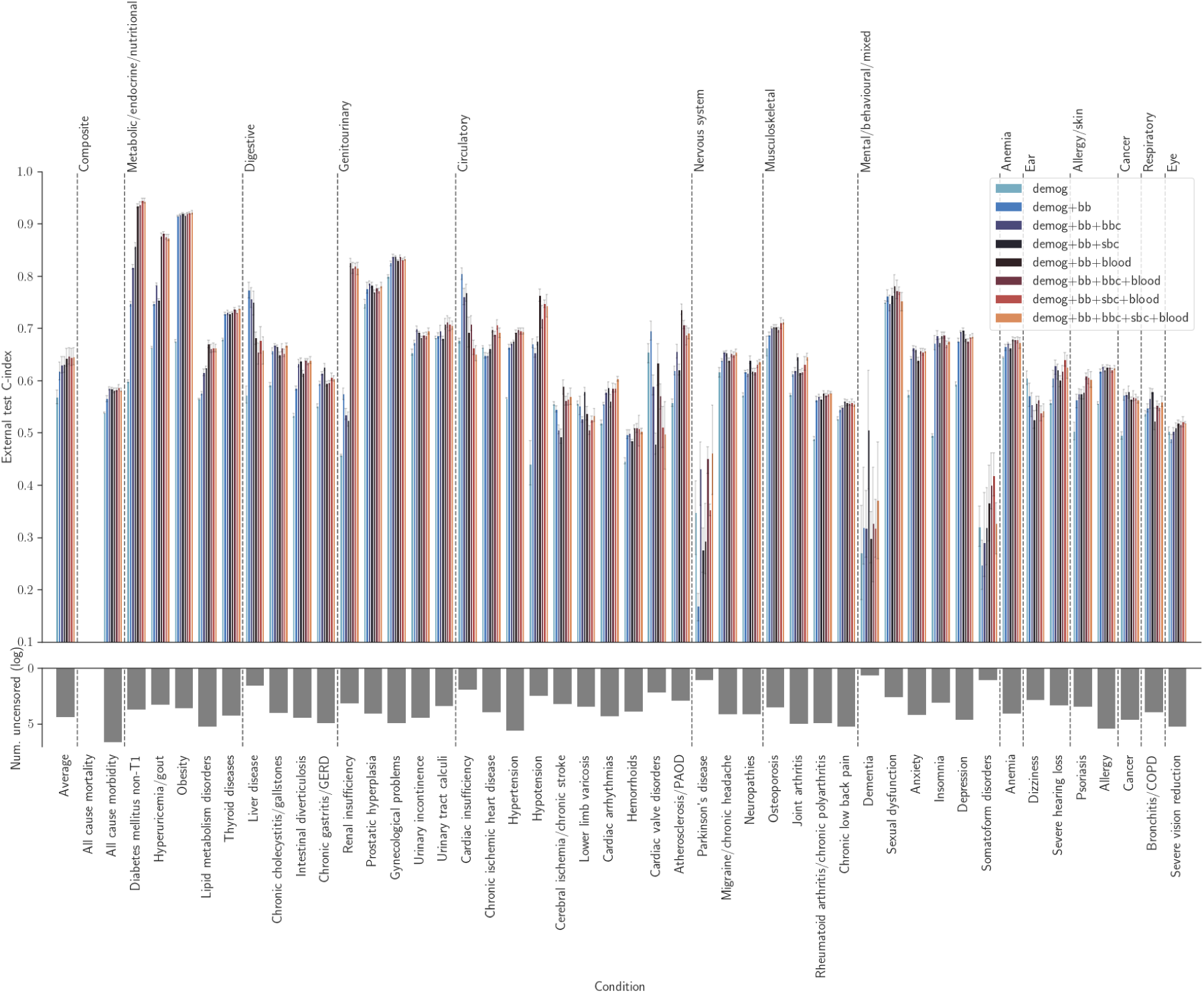
External test set C-index across disease conditions for neural network architectures. Gray error bars illustrate 95% confidence intervals.

### C.3 Kaplan-Meier curves

**Fig. C7:**
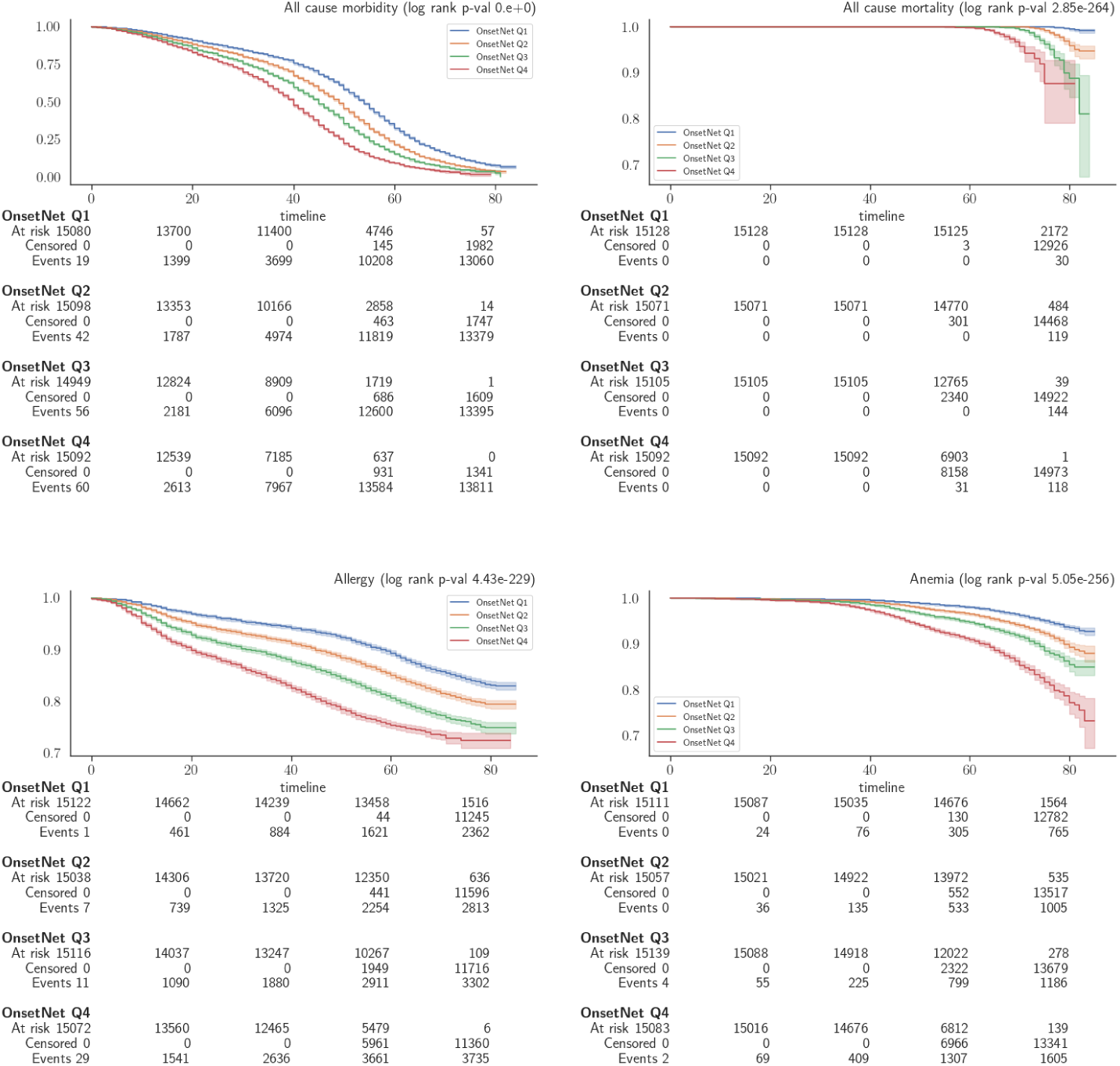
Kaplan-Meier curves plotting survival probability (vertical) against age in years (horizontal). (1/9)

**Fig. C8:**
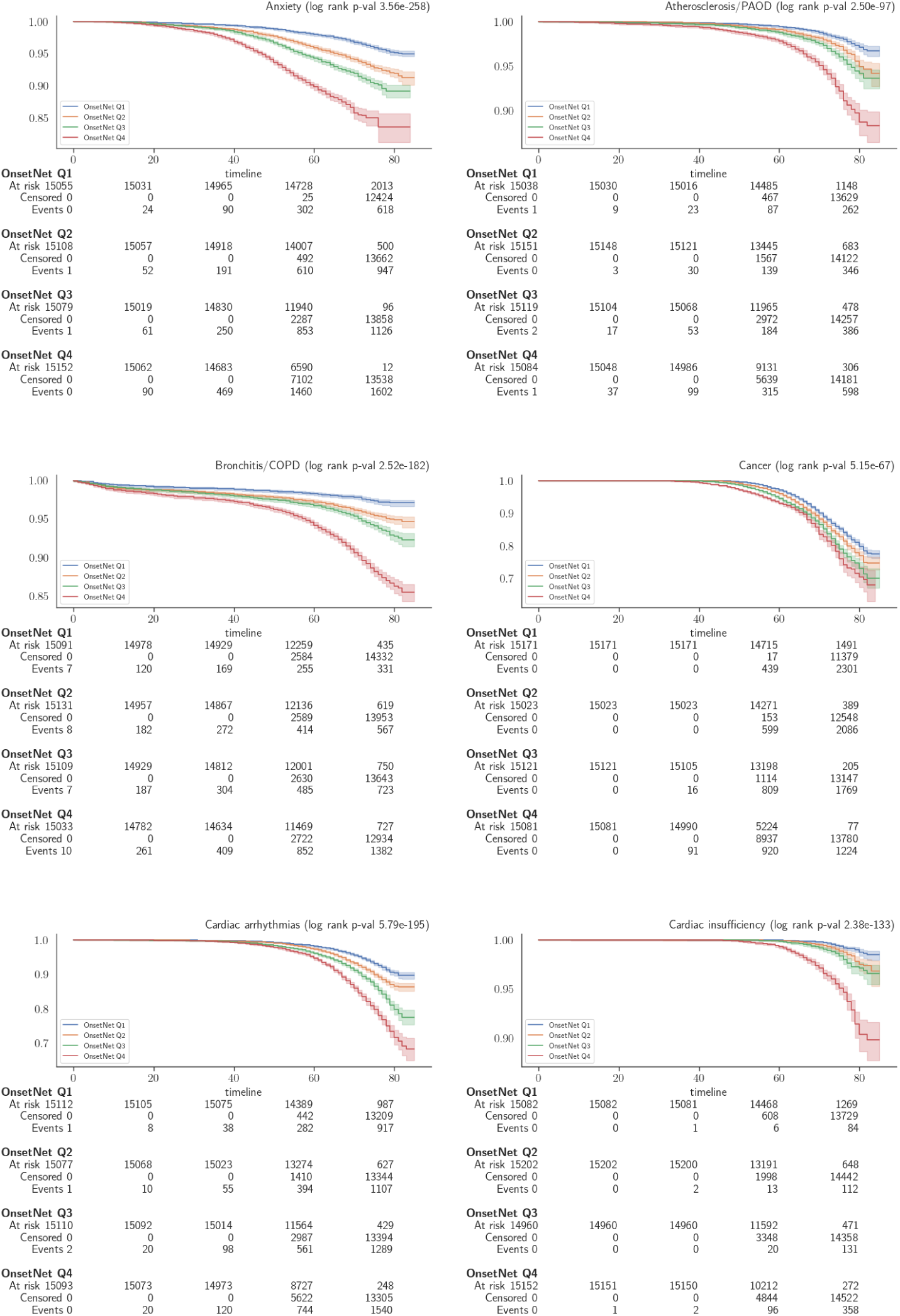
Kaplan-Meier curves plotting survival probability (vertical) against age in years (horizontal). (2/9)

**Fig. C9:**
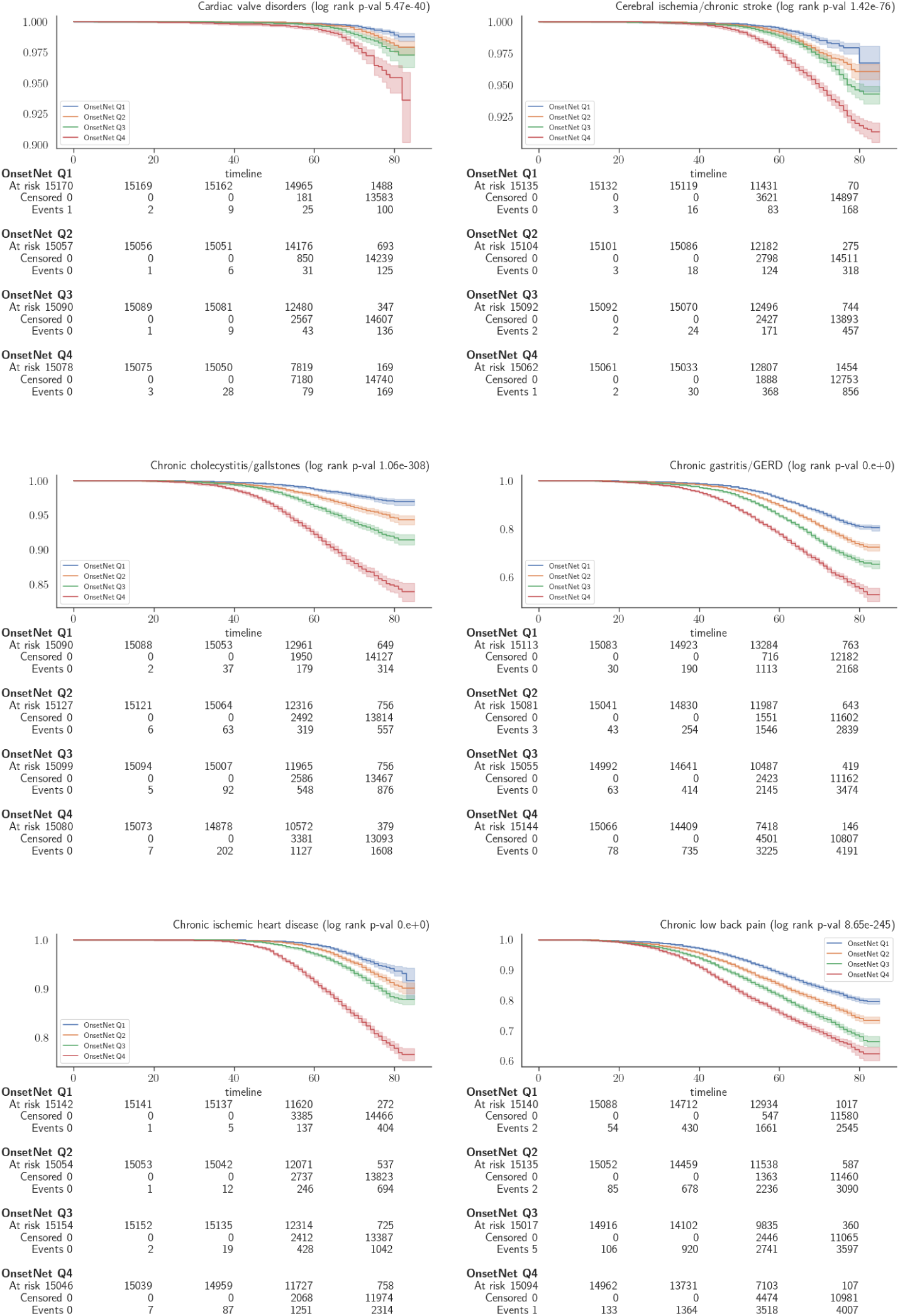
Kaplan-Meier curves plotting survival probability (vertical) against age in years (horizontal). (3/9)

**Fig. C10:**
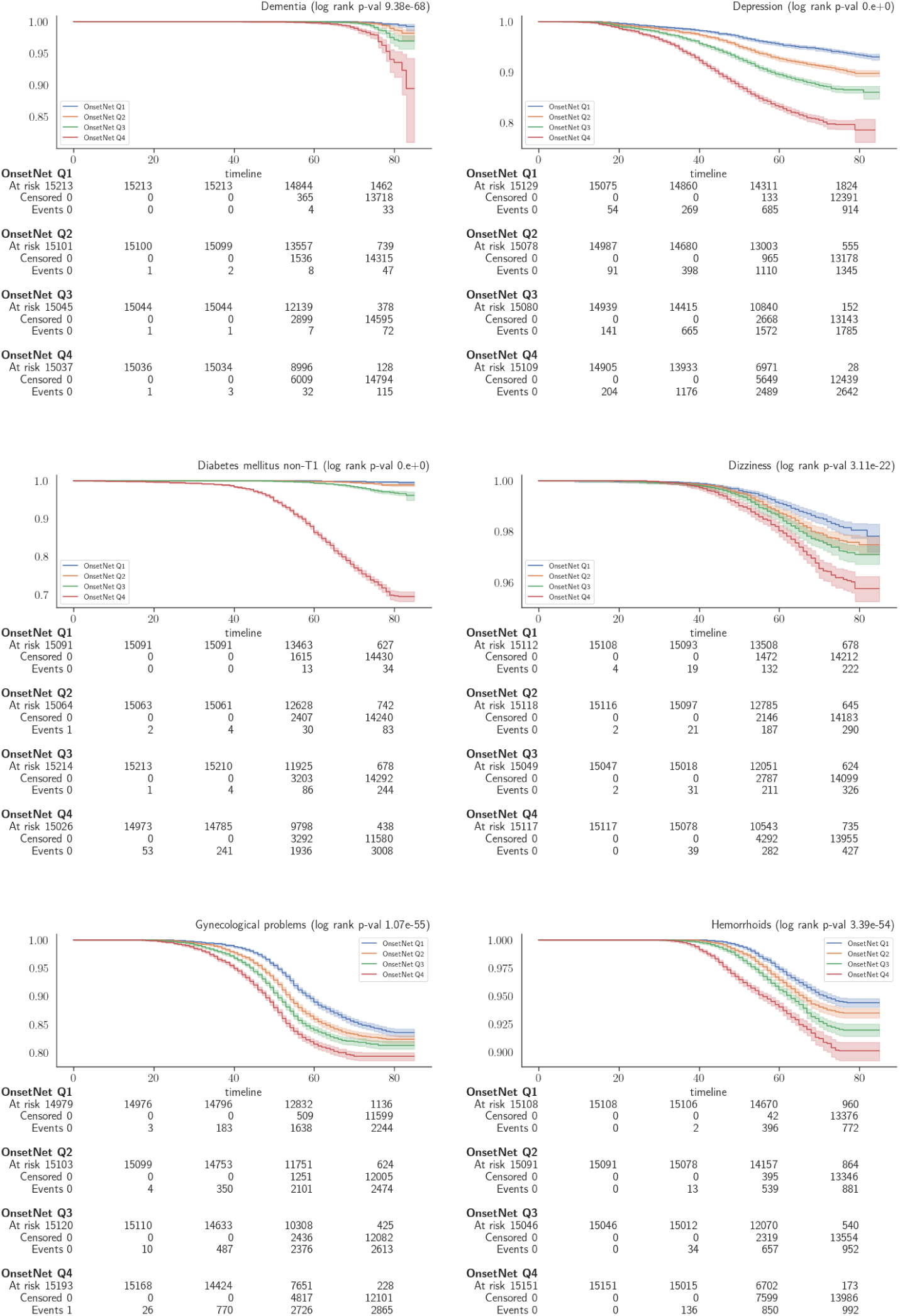
Kaplan-Meier curves plotting survival probability (vertical) against age in years (horizontal). (4/9)

**Fig. C11:**
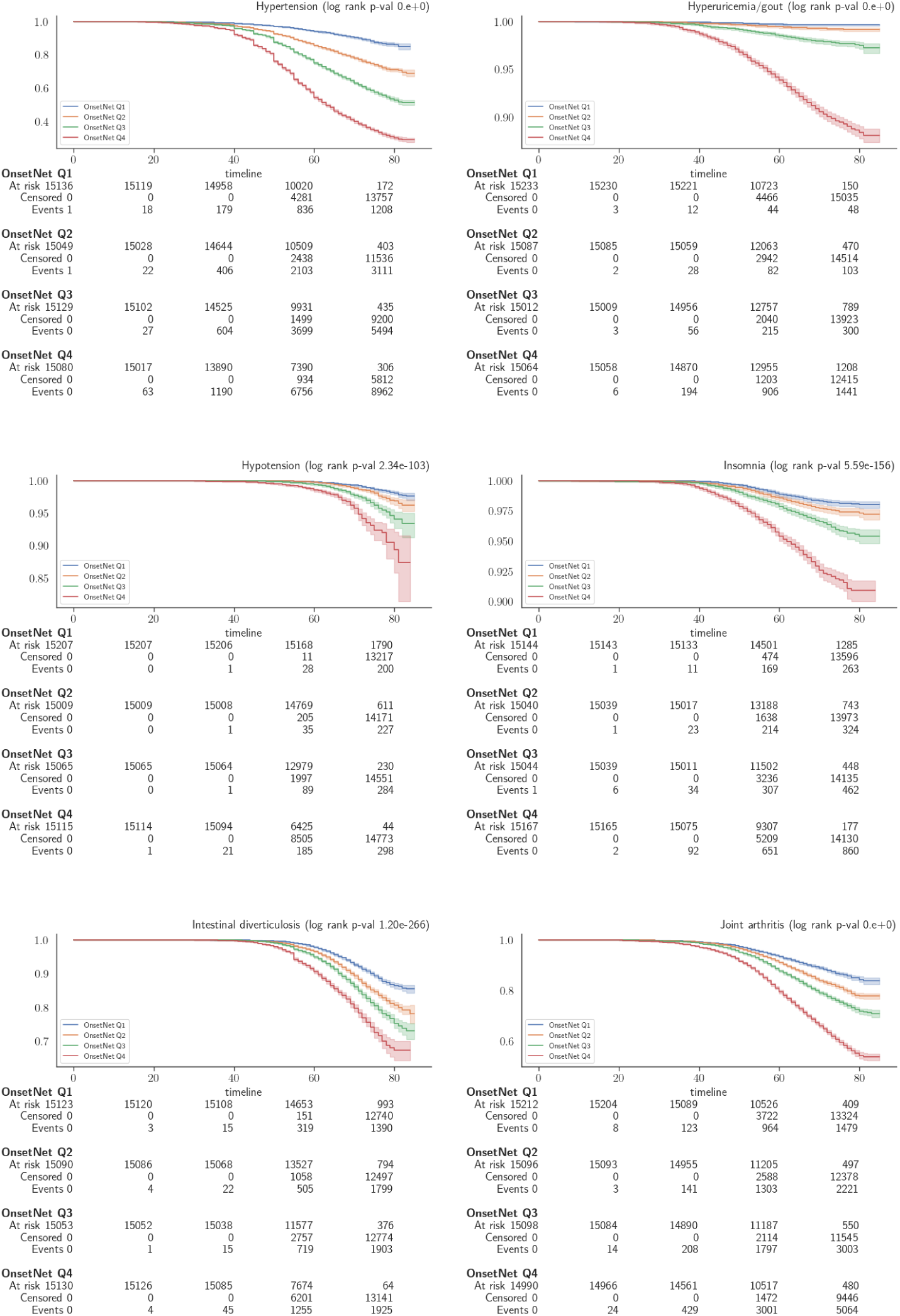
Kaplan-Meier curves plotting survival probability (vertical) against age in years (horizontal). (5/9)

**Fig. C12:**
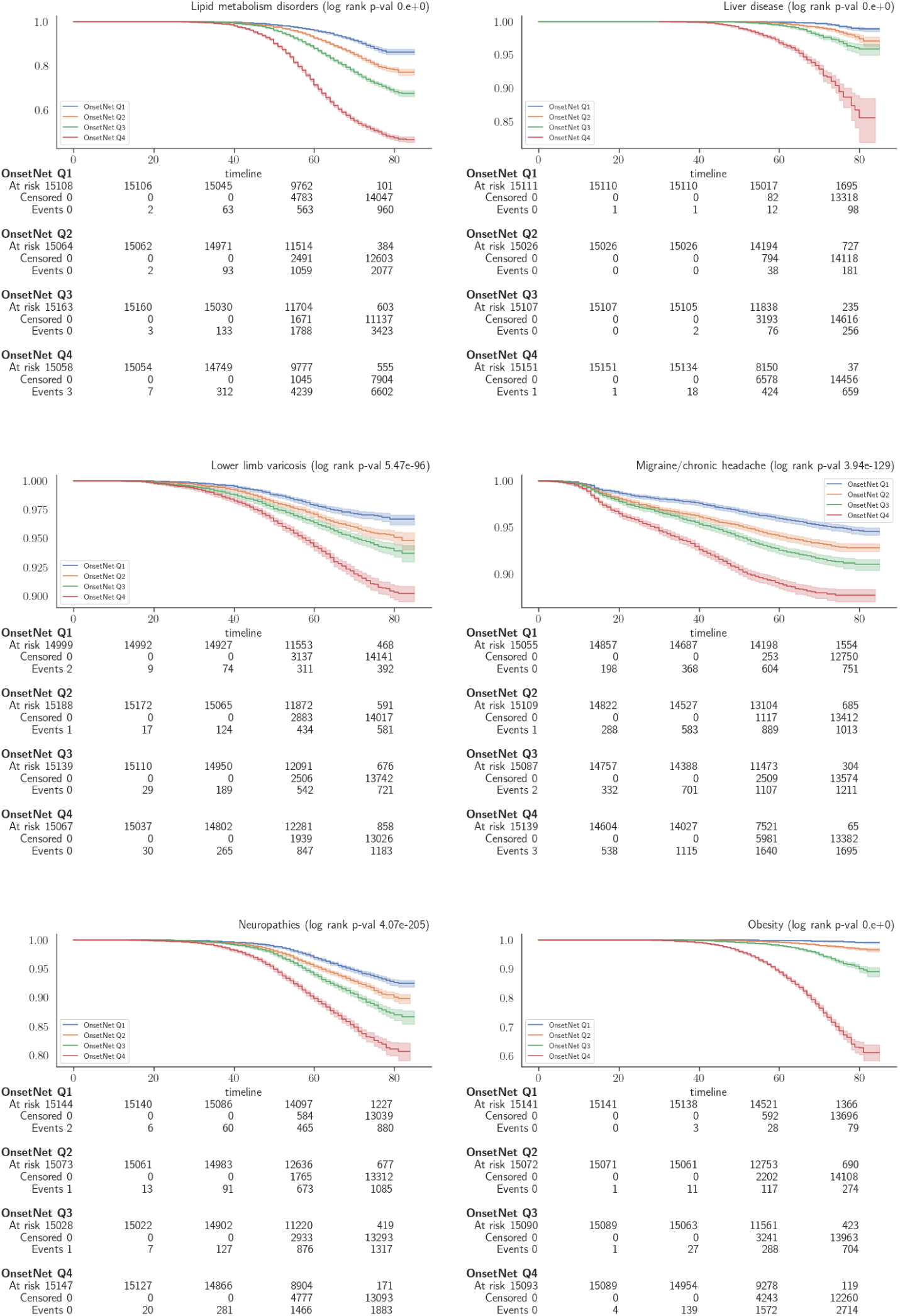
Kaplan-Meier curves plotting survival probability (vertical) against age in years (horizontal). (6/9)

**Fig. C13:**
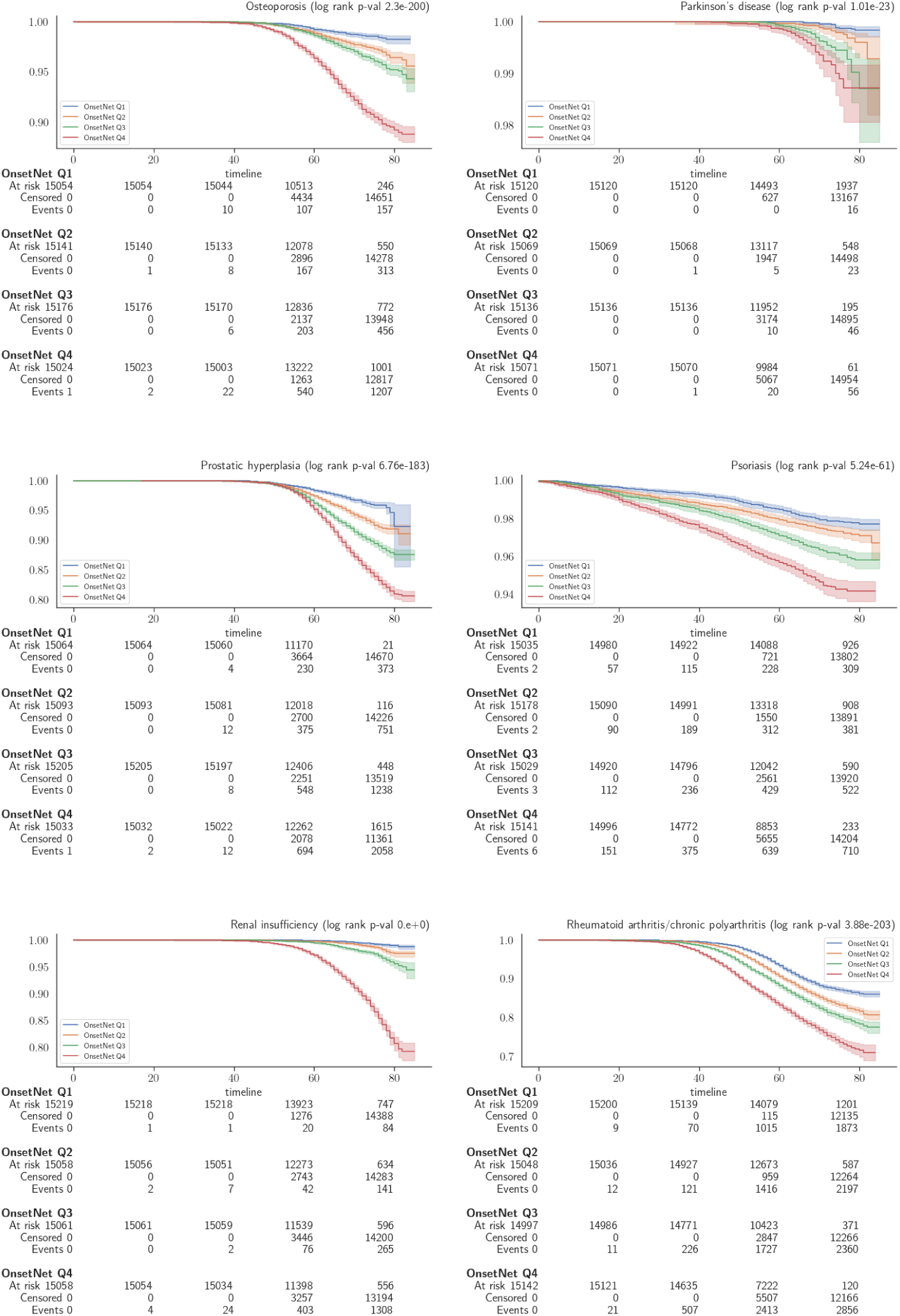
Kaplan-Meier curves plotting survival probability (vertical) against age in years (horizontal). (7/9)

**Fig. C14:**
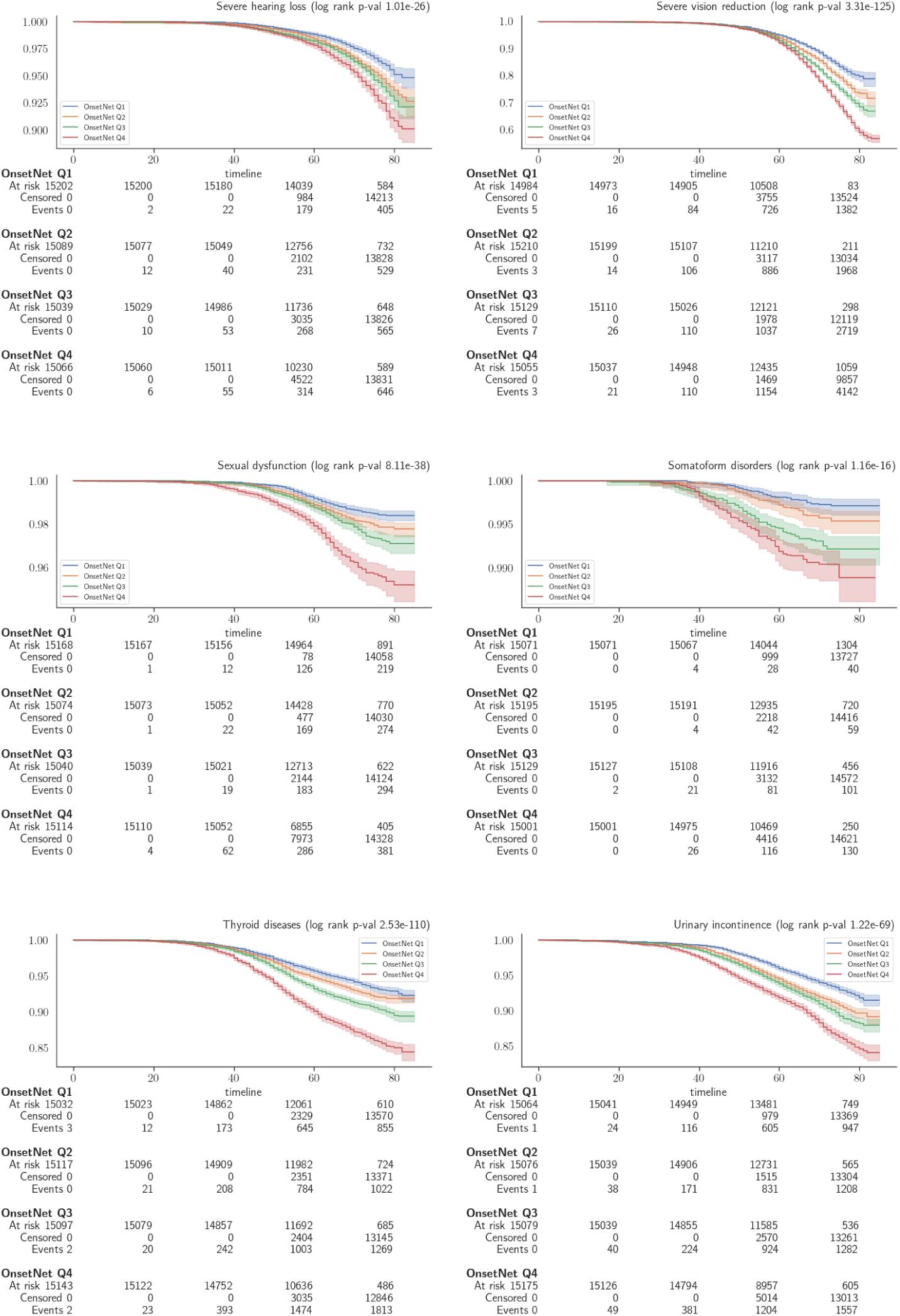
Kaplan-Meier curves plotting survival probability (vertical) against age in years (horizontal). (8/9)

**Fig. C15:**
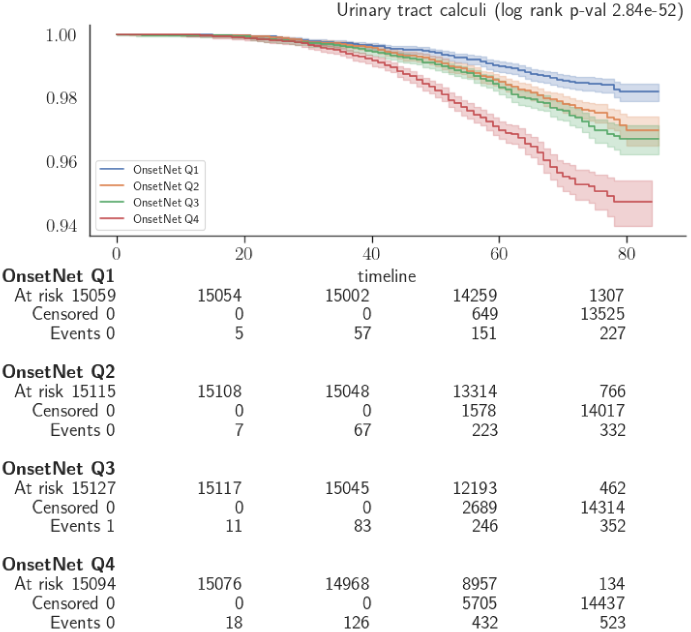
Kaplan-Meier curves plotting survival probability (vertical) against age in years (horizontal). (9/9)

### C.4 Inter-disease risk analysis

#### C.4.1 Onset acceleration correlation

The clustering dendrogram in Figure 5 utilized UPGMA (unweighted pair group method with arithmetic mean) hierarchical agglomerative clustering for the linkage function (SciPy *cluster.hierarchy.linkage*) with cosine distance between correlation vectors as the clustering metric, where 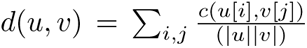 defined the linkage function between non-identical clusters *u* and *v*, *i* (*j*) indexes members of cluster *u* (*v*), and *c* denotes cosine distance. Color red (blue) indicates positive (negative)

**Fig. C16:**
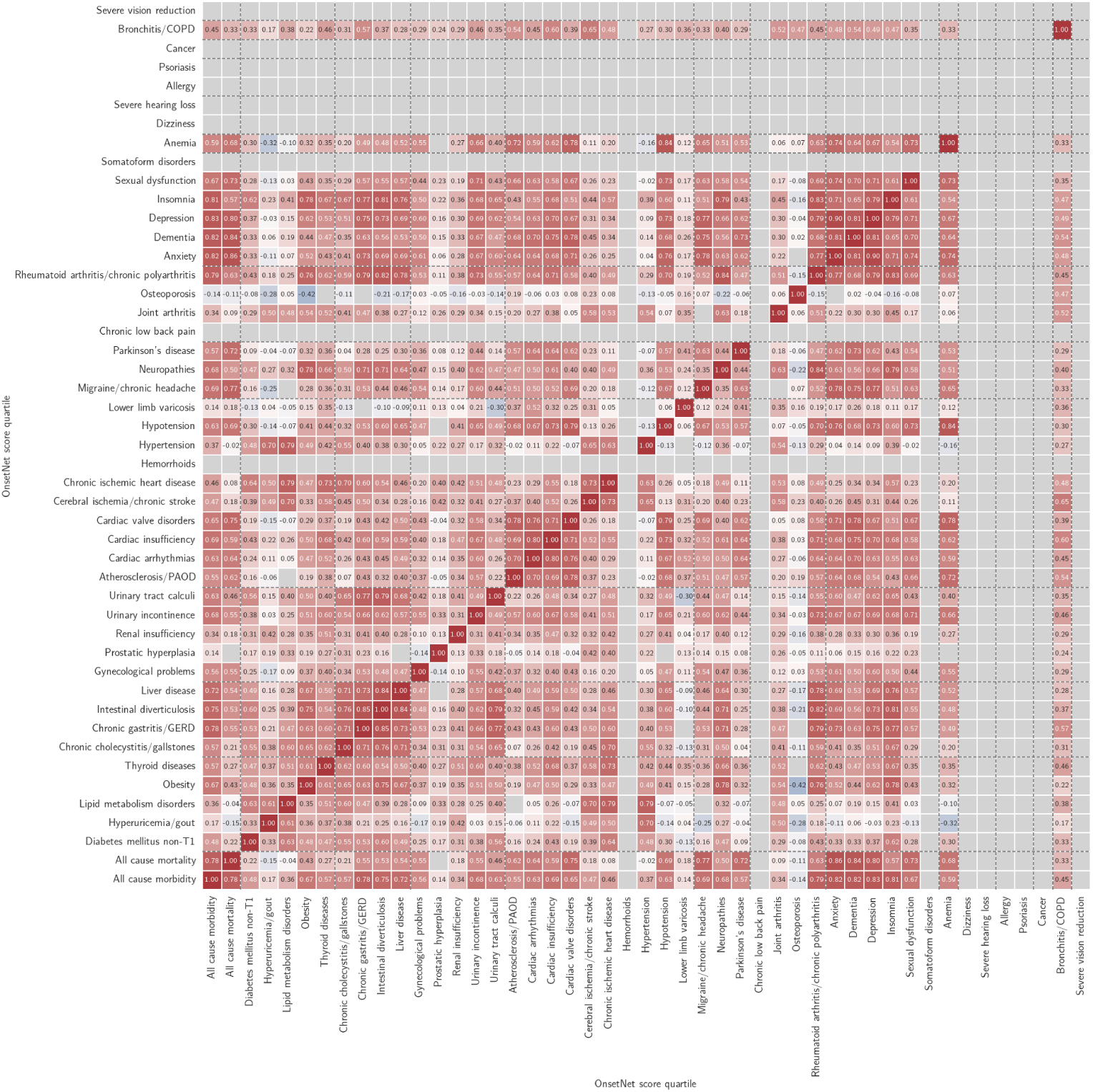
All inter-disease Pearson correlations *r* for non-statin users. Color red (blue) indicates positive (negative) correlation and color intensity scales with correlation magnitude. Conditions with internal test C-index *<* 0.6 and correlations with *p >* 3.46*E* − 5 (Bonferroni corrected) indicated in gray.

**Fig. C17:**
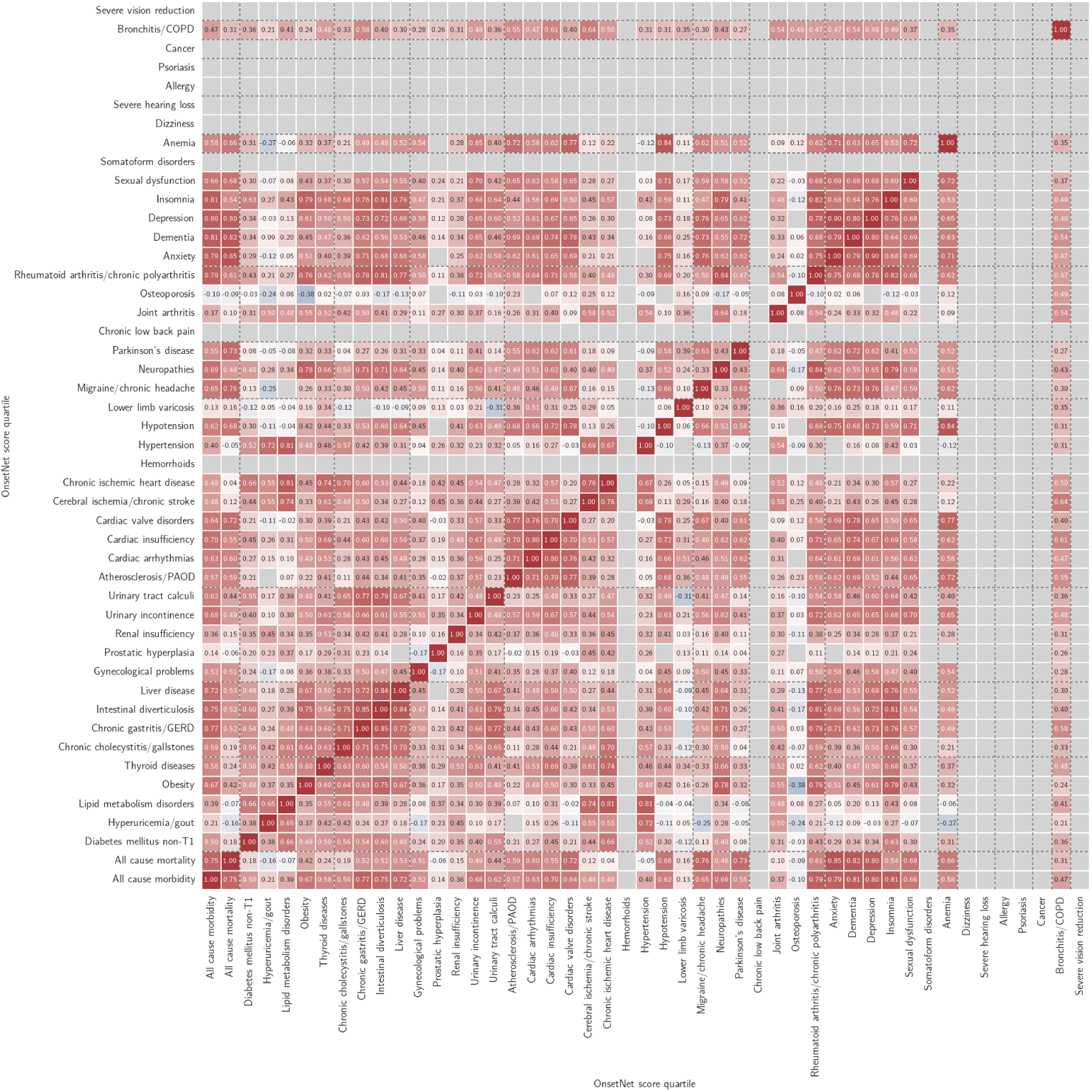
All inter-disease Pearson correlations *r*, statin users included. Color red (blue) indicates positive (negative) correlation and color intensity scales with correlation magnitude. Conditions with internal test C-index *<* 0.6 and correlations with *p >* 3.46*E* − 5 (Bonferroni corrected) indicated in gray.

**Fig. C18:**
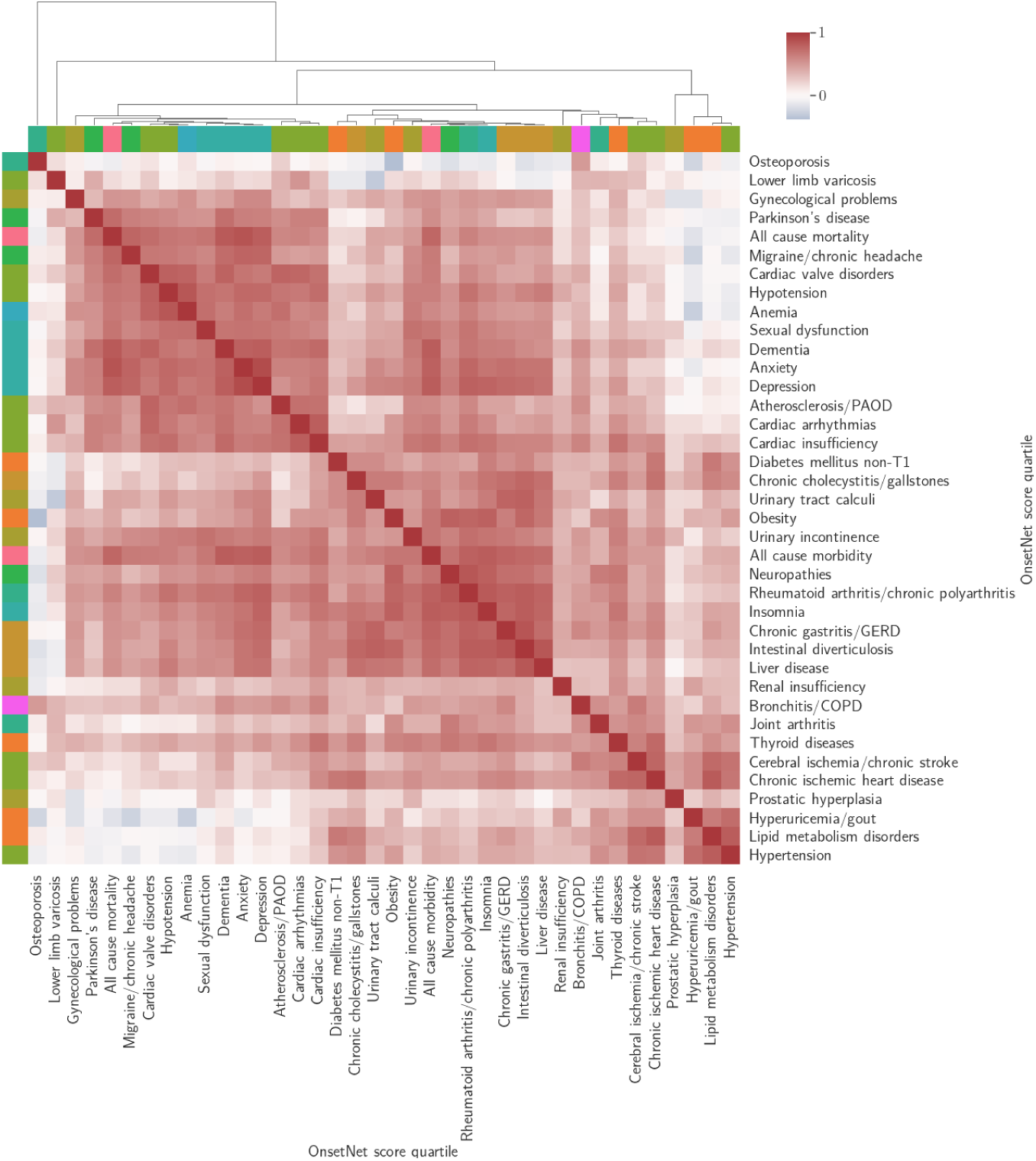
All-to-all OnsetNet score quartile correlation for conditions with internal test C-index ≥ 0.6, clustered by cosine distance between correlation vectors. Color red (blue) indicates positive (negative) correlation and color intensity scales with correlation magnitude. Statin users were included and 28 out of 1444 entries with *p >* 3.46*E* − 5 (Bonferroni corrected) were set to 0 for cosine distance computation.

**Fig. C19:**
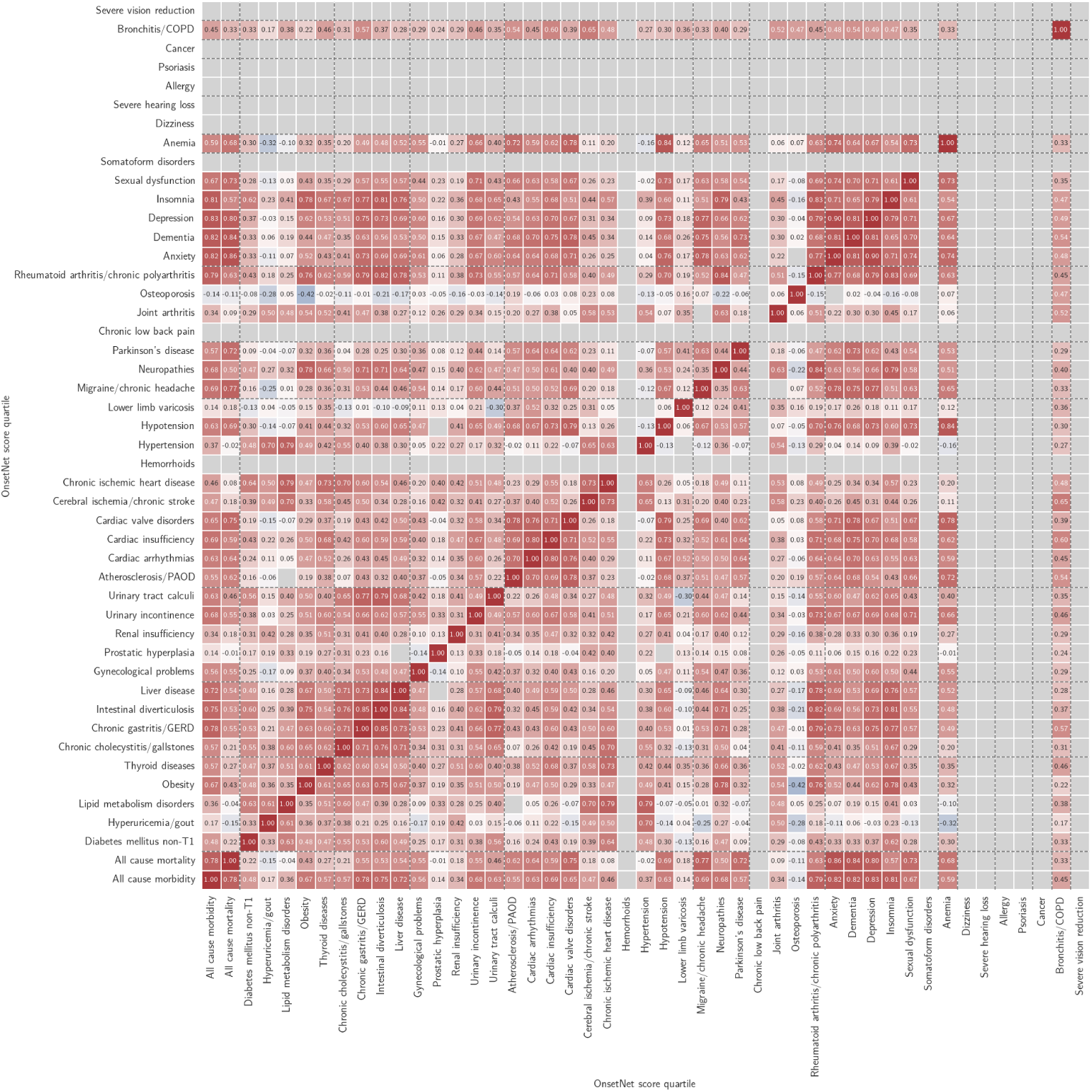
All inter-disease Pearson correlations *r* for non-statin users without Bonferroni correction. Color red (blue) indicates positive (negative) correlation and color intensity scales with correlation magnitude. Conditions with internal test C-index *<* 0.6 and correlations with *p >* 0.05 indicated in gray.

**Fig. C20:**
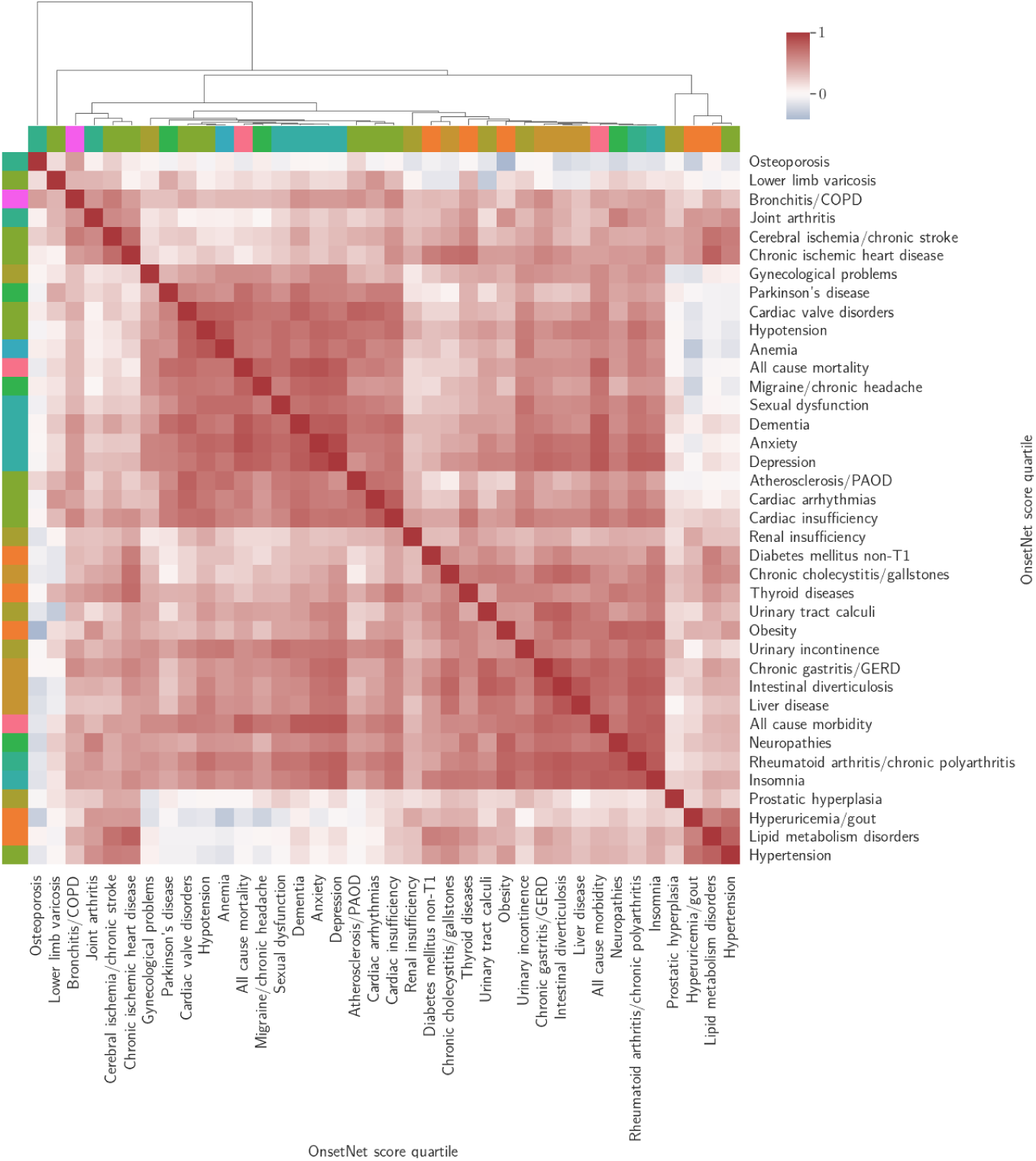
All-to-all OnsetNet score quartile correlation for non-statin users and conditions with internal test C-index ≥ 0.6, clustered by cosine distance between correlation vectors. Color red (blue) indicates positive (negative) correlation and color intensity scales with correlation magnitude. 12 out of 1444 entries with *p >* 0.05 (no Bonferroni correction) were set to 0 for cosine distance computation.

#### C.4.2 Associations in onset acceleration correlation

**Table C11:** Top 5 conditions with highest onset acceleration risk quartile correlation across non-statin users for conditions with internal test C-index ≥ 0.6. All correlations in the top 5 were statistically significant (*p* ≤ 9.41*E* − 77 ± 1.29*E* − 75).

| Condition | Top 5 conditions by onset acceleration risk correlation |
| --- | --- |
| All cause morbidity | Depression (0.83), anxiety (0.82), dementia (0.82), insomnia (0.81), rheumatoid arthritis/chronic polyarthritis (0.79) |
| All cause mortality | Anxiety (0.86), dementia (0.84), depression (0.80), all cause morbidity (0.78), migraine/chronic headache (0.77) |
| Diabetes mellitus non-T1 | Chronic ischemic heart disease (0.64), lipid metabolism disorders (0.63), insomnia (0.62), intestinal diverticulosis (0.60), urinary tract calculi (0.56) |
| Hyperuricemia/gout | Hypertension (0.70), lipid metabolism disorders (0.61), joint arthritis (0.50), chronic ischemic heart disease (0.50), cerebral ischemia/chronic stroke (0.49) |
| Lipid metabolism disorders | Chronic ischemic heart disease (0.79), hypertension (0.79), cerebral ischemia/chronic stroke (0.70), diabetes mellitus non-T1 (0.63), hyperuricemia/gout (0.61) |
| Obesity | Insomnia (0.78), neuropathies (0.78), rheumatoid arthritis/chronic polyarthritis (0.76), intestinal diverticulosis (0.75), liver disease (0.67) |
| Thyroid diseases | Chronic ischemic heart disease (0.73), cardiac insufficiency (0.68), insomnia (0.67), neuropathies (0.66), rheumatoid arthritis/chronic polyarthritis (0.62) |
| Chronic cholecystitis/ gallstones | Intestinal diverticulosis (0.76), chronic gastritis/GERD (0.71), liver disease (0.71), chronic ischemic heart disease (0.70), insomnia (0.67) |
| Chronic gastritis/GERD | Intestinal diverticulosis (0.85), rheumatoid arthritis/chronic polyarthritis (0.79), all cause morbidity (0.78), urinary tract calculi (0.77), insomnia (0.77) |

**Table C11 (continued)**
|  |  |
| --- | --- |
| Intestinal diverticu-<br>losis | Chronic gastritis/GERD (0.85), liver disease (0.84), rheumatoid arthritis/chronic polyarthritis (0.82), insomnia (0.81), urinary tract calculi (0.79) |
| Liver disease | Intestinal diverticulosis (0.84), rheumatoid arthritis/chronic polyarthritis (0.78), insomnia (0.76), chronic gastritis/GERD (0.73), all cause morbidity (0.72) |
| Gynecological prob-<br>lems | Anxiety (0.61), depression (0.60), all cause morbidity (0.56), all cause mortality (0.55), anemia (0.55) |
| Prostatic hyperpla-<br>sia | Cerebral ischemia/chronic stroke (0.42), chronic ischemic heart disease (0.40), lipid metabolism disorders (0.33), urinary incontinence (0.33), chronic cholecystitis/gallstones (0.31) |
| Renal insufficiency | Thyroid diseases (0.51), cardiac insufficiency (0.47), hyperuricemia/-gout (0.42), chronic ischemic heart disease (0.42), hypotension (0.41) |
| Urinary inconti-<br>nence | Rheumatoid arthritis/chronic polyarthritis (0.73), sexual dysfunction (0.71), depression (0.69), insomnia (0.68), all cause morbidity (0.68) |
| Urinary tract cal-<br>culi | Intestinal diverticulosis (0.79), chronic gastritis/GERD (0.77), liver disease (0.68), chronic cholecystitis/gallstones (0.65), insomnia (0.65) |
| Atherosclerosis/<br>PAOD | Cardiac valve disorders (0.78), anemia (0.72), cardiac arrhythmias (0.70), cardiac insufficiency (0.69), dementia (0.68) |
| Cardiac arrhyth-<br>mias | Cardiac insufficiency (0.80), cardiac valve disorders (0.76), dementia (0.70), atherosclerosis/PAOD (0.70), hypotension (0.67) |
| Cardiac insuffi-<br>ciency | Cardiac arrhythmias (0.80), dementia (0.75), hypotension (0.73), rheumatoid arthritis/chronic polyarthritis (0.71), cardiac valve disorders (0.71) |
| Cardiac valve disor-<br>ders | Hypotension (0.79), atherosclerosis/PAOD (0.78), dementia (0.78), anemia (0.78), cardiac arrhythmias (0.76) |

**Table C11 (continued)**
|  |  |
| --- | --- |
| Cerebral ischemia/<br>chronic stroke | Chronic ischemic heart disease (0.73), lipid metabolism disorders (0.70), bronchitis/COPD (0.65), hypertension (0.65), thyroid diseases (0.58) |
| Chronic ischemic<br>heart disease | Lipid metabolism disorders (0.79), thyroid diseases (0.73), cerebral ischemia/chronic stroke (0.73), chronic cholecystitis/gallstones (0.70), diabetes mellitus non-T1 (0.64) |
| Hypertension | Lipid metabolism disorders (0.79), hyperuricemia/gout (0.70), cerebral ischemia/chronic stroke (0.65), chronic ischemic heart disease (0.63), chronic cholecystitis/gallstones (0.55) |
| Hypotension | Anemia (0.84), cardiac valve disorders (0.79), anxiety (0.76), depression (0.73), sexual dysfunction (0.73) |
| Lower limb varico-<br>sis | Cardiac arrhythmias (0.52), parkinson's disease (0.41), atherosclerosis/PAOD (0.37), bronchitis/COPD (0.36), thyroid diseases (0.35) |
| Migraine/ chronic<br>headache | Anxiety (0.78), depression (0.77), all cause mortality (0.77), dementia (0.75), all cause morbidity (0.69) |
| Neuropathies | Rheumatoid arthritis/chronic polyarthritis (0.84), insomnia (0.79), obesity (0.78), chronic gastritis/GERD (0.71), intestinal diverticulosis (0.71) |
| Parkinson's disease | Dementia (0.73), all cause mortality (0.72), cardiac arrhythmias (0.64), cardiac insufficiency (0.64), migraine/chronic headache (0.63) |
| Joint arthritis | Neuropathies (0.63), cerebral ischemia/chronic stroke (0.58), hypertension (0.54), obesity (0.54), chronic ischemic heart disease (0.53) |
| Osteoporosis | Bronchitis/COPD (0.47), cerebral ischemia/chronic stroke (0.23), atherosclerosis/PAOD (0.19), lower limb varicosis (0.16), chronic ischemic heart disease (0.08) |
| Rheumatoid<br>arthritis/ chronic<br>polyarthritis | Neuropathies (0.84), insomnia (0.83), intestinal diverticulosis (0.82), chronic gastritis/GERD (0.79), depression (0.79) |

**Table C11 (continued)**
|  |  |
| --- | --- |
| Anxiety | Depression (0.90), all cause mortality (0.86), all cause morbidity (0.82), dementia (0.81), migraine/chronic headache (0.78) |
| Dementia | All cause mortality (0.84), all cause morbidity (0.82), depression (0.81), anxiety (0.81), cardiac valve disorders (0.78) |
| Depression | Anxiety (0.90), all cause morbidity (0.83), dementia (0.81), all cause mortality (0.80), rheumatoid arthritis/chronic polyarthritis (0.79) |
| Insomnia | Rheumatoid arthritis/chronic polyarthritis (0.83), intestinal diverticulosis (0.81), all cause morbidity (0.81), neuropathies (0.79), depression (0.79) |
| Sexual dysfunction | Anxiety (0.74), all cause mortality (0.73), anemia (0.73), hypotension (0.73), depression (0.71) |
| Anemia | Hypotension (0.84), cardiac valve disorders (0.78), anxiety (0.74), sexual dysfunction (0.73), atherosclerosis/PAOD (0.72) |
| Bronchitis/ COPD | Cerebral ischemia/chronic stroke (0.65), cardiac insufficiency (0.60), chronic gastritis/GERD (0.57), dementia (0.54), atherosclerosis/-PAOD (0.54) |

#### C.4.3 Prognostic survival analysis

##### Prognostic risk evaluation

Although commonly used exclusively for prognostic prediction in biomedical contexts [27], Cox PH models do not assume events must occur after trait measurement baseline. The Cox PH objective function specifies a well-defined optimization problem for risk prediction functions regardless of whether variable *x_i_* (Section 2.3) contains traits of the individual measured before or after the event occurred. Model *h^d^* was trained and evaluated as a general (not exclusively prognostic) risk prediction model taking variable *x_i_* containing imperfect information for event time of condition *d* whether retrospective or prospective. Prognostic value of the model was separately verified in Section 3.3 with exclusion of events that occurred before trait measurement time.

**Fig. C21:**
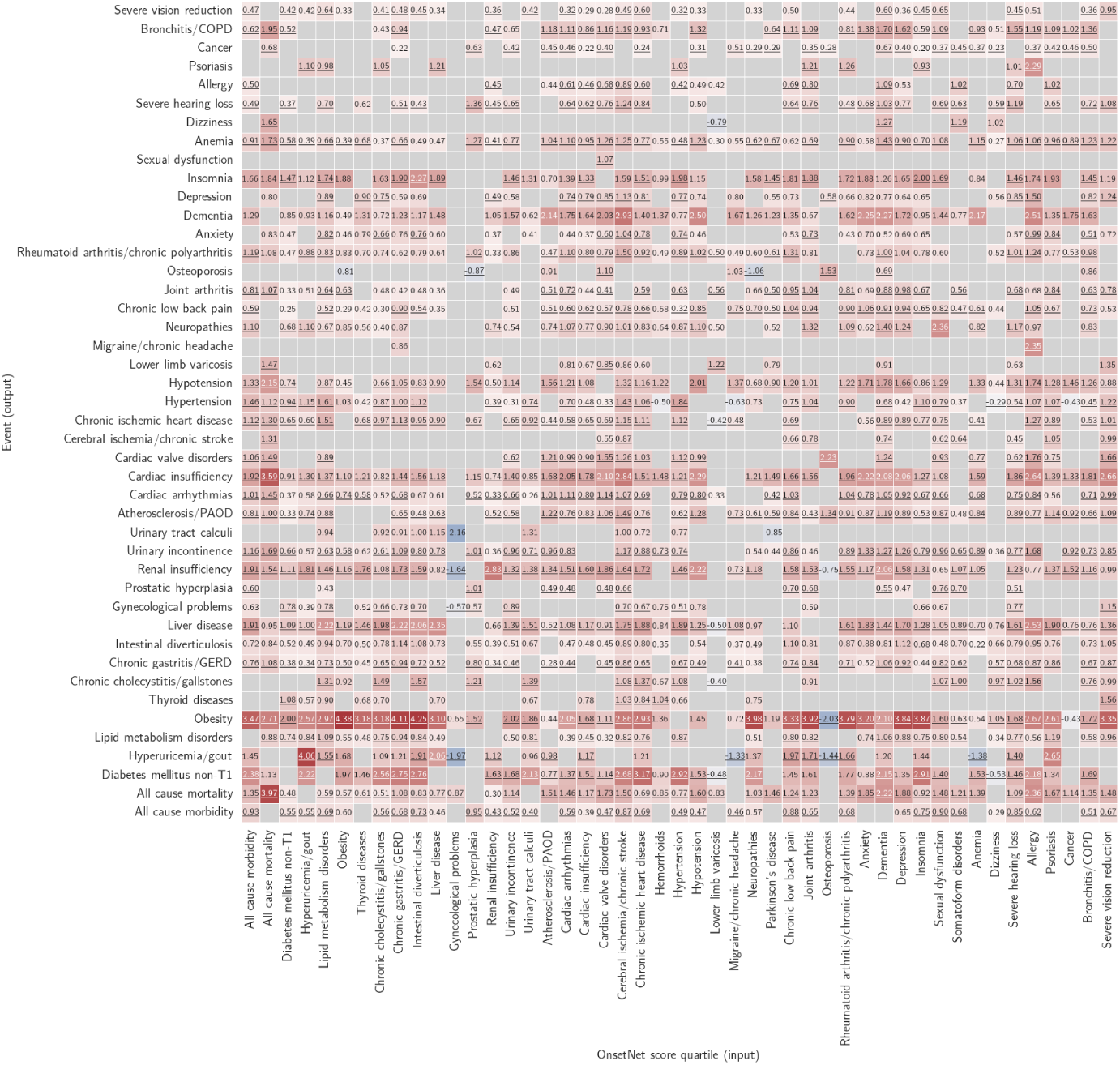
All-to-all adjusted log hazard ratios for OnsetNet risk Q4 (Q1 as reference) given primary adjustment with demographic variables. Color red (blue) indicates positive (negative) log hazard ratios and color intensity scales with log hazard ratio magnitude. Results with adjusted hazard ratio *p >* 0.05 (without Bonferroni correction) or Schoenfeld residual *p* ≤ 0.05 omitted in gray. Entries that remained statistically significant after secondary adjustment with basic body traits are underlined to denote robustness to change in adjustment.

**Fig. C22:**
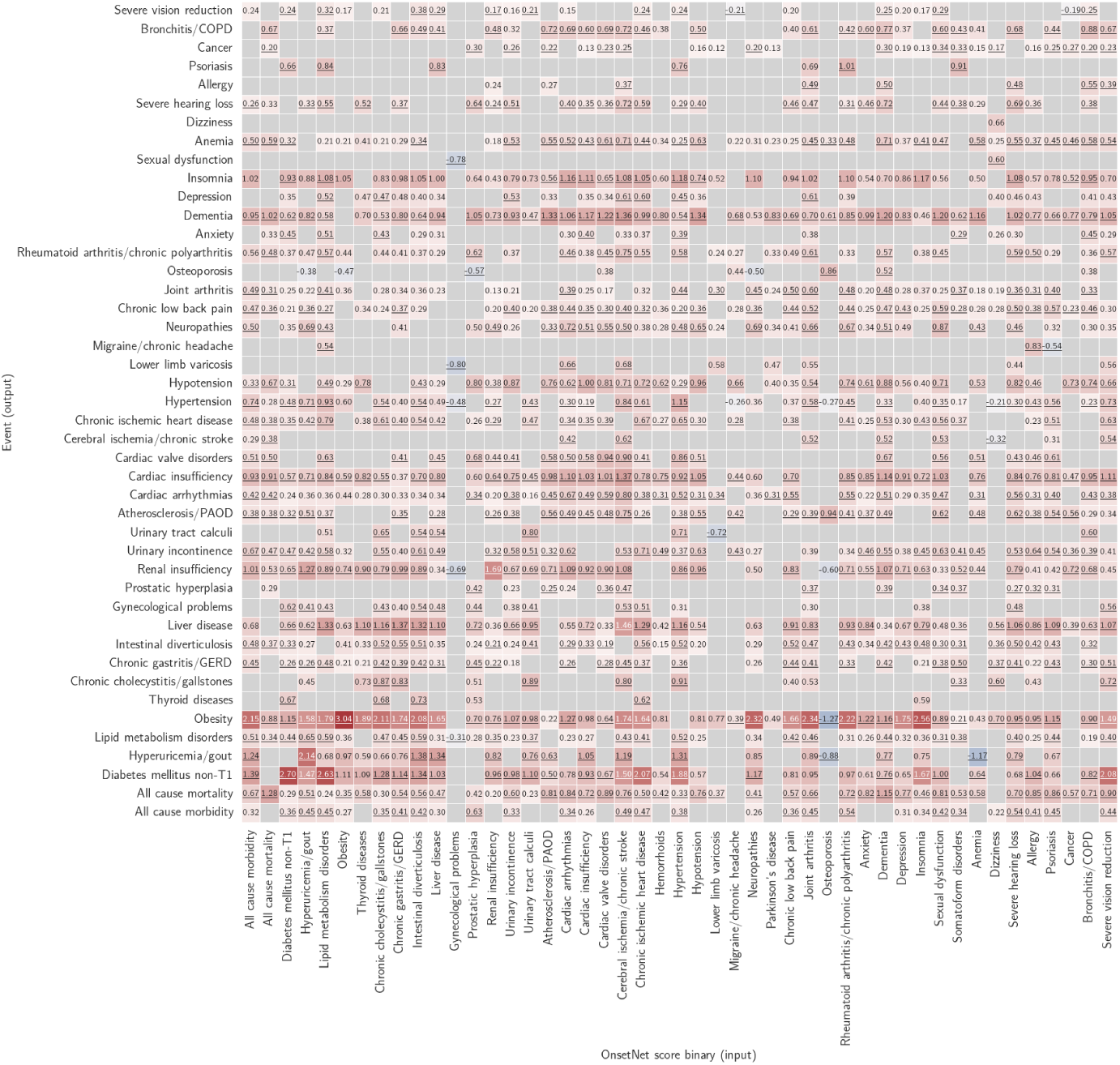
All-to-all log adjusted hazard ratios for binarized OnsetNet risk (above-median; below-median as reference) given primary adjustment with demographic variables. Color red (blue) indicates positive (negative) log hazard ratios and color intensity scales with log hazard ratio magnitude. Results with adjusted hazard ratio *p >* 0.05 (without Bonferroni correction) or Schoenfeld residual *p* ≤ 0.05 omitted in gray. Entries that remained statistically significant after secondary adjustment with basic body traits are underlined to denote robustness to change in adjustment.

**Fig. C23:**
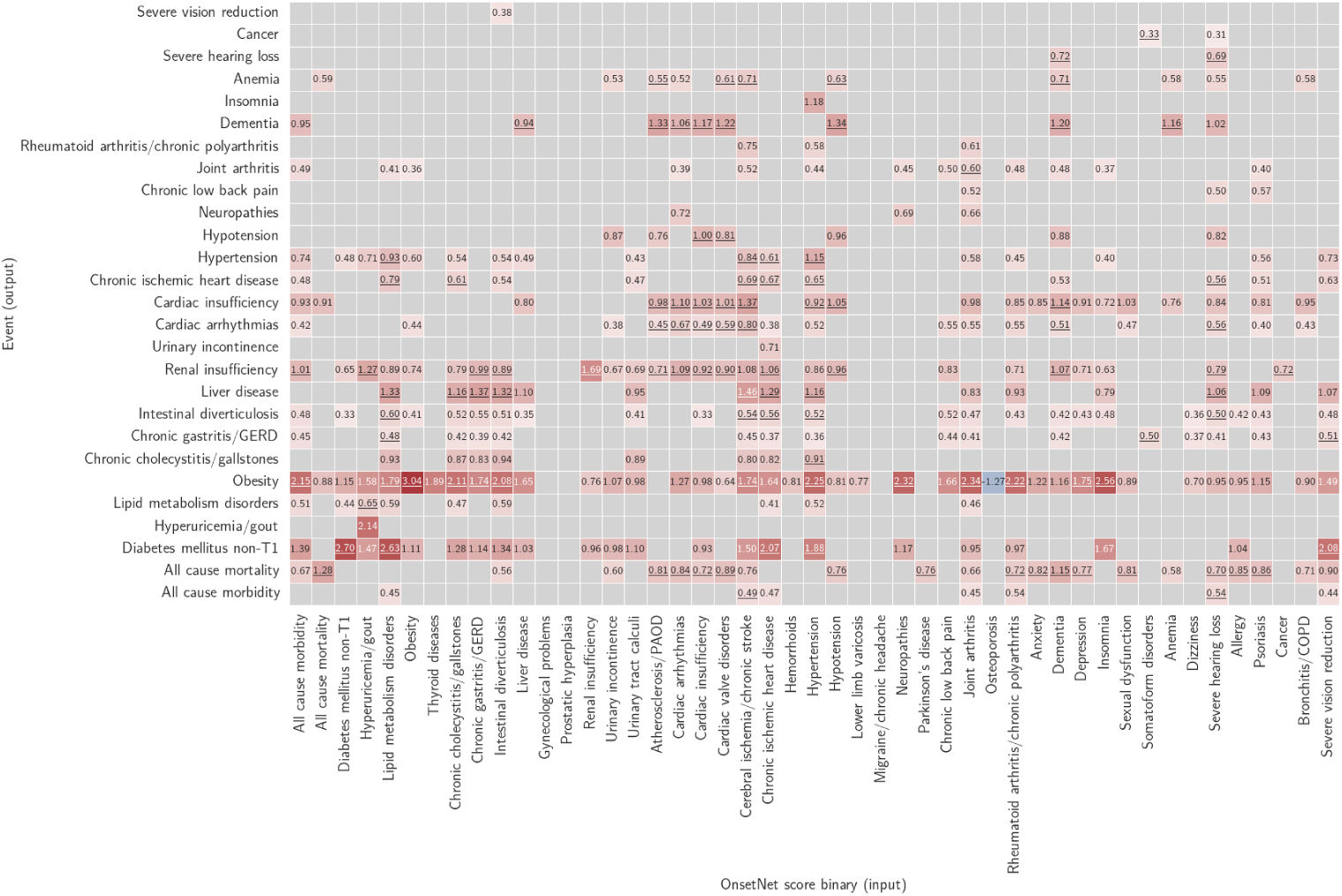
All-to-all log adjusted hazard ratios for binarized OnsetNet risk (above-median; below-median as reference) given primary adjustment with demographic variables. Color red (blue) indicates positive (negative) log hazard ratios and color intensity scales with log hazard ratio magnitude. Statistically insignificant adjusted hazard ratios with *p >* 9.05*E* − 7 (Bonferroni corrected *p >* 0.05) or Schoenfeld residual *p* ≤ 0.05 are omitted in gray. Entries that remained statistically significant after secondary adjustment with basic body traits are underlined to denote robustness to change in adjustment.

### C.5 Biomarker saliency

Saliency maps for full traits (demog+bb+bbc+sbc+blood) and full univariate traits (demog+bb+bbc+blood) OnsetNet models are provided.

#### C.5.1 Full traits model

**Fig. C24:**
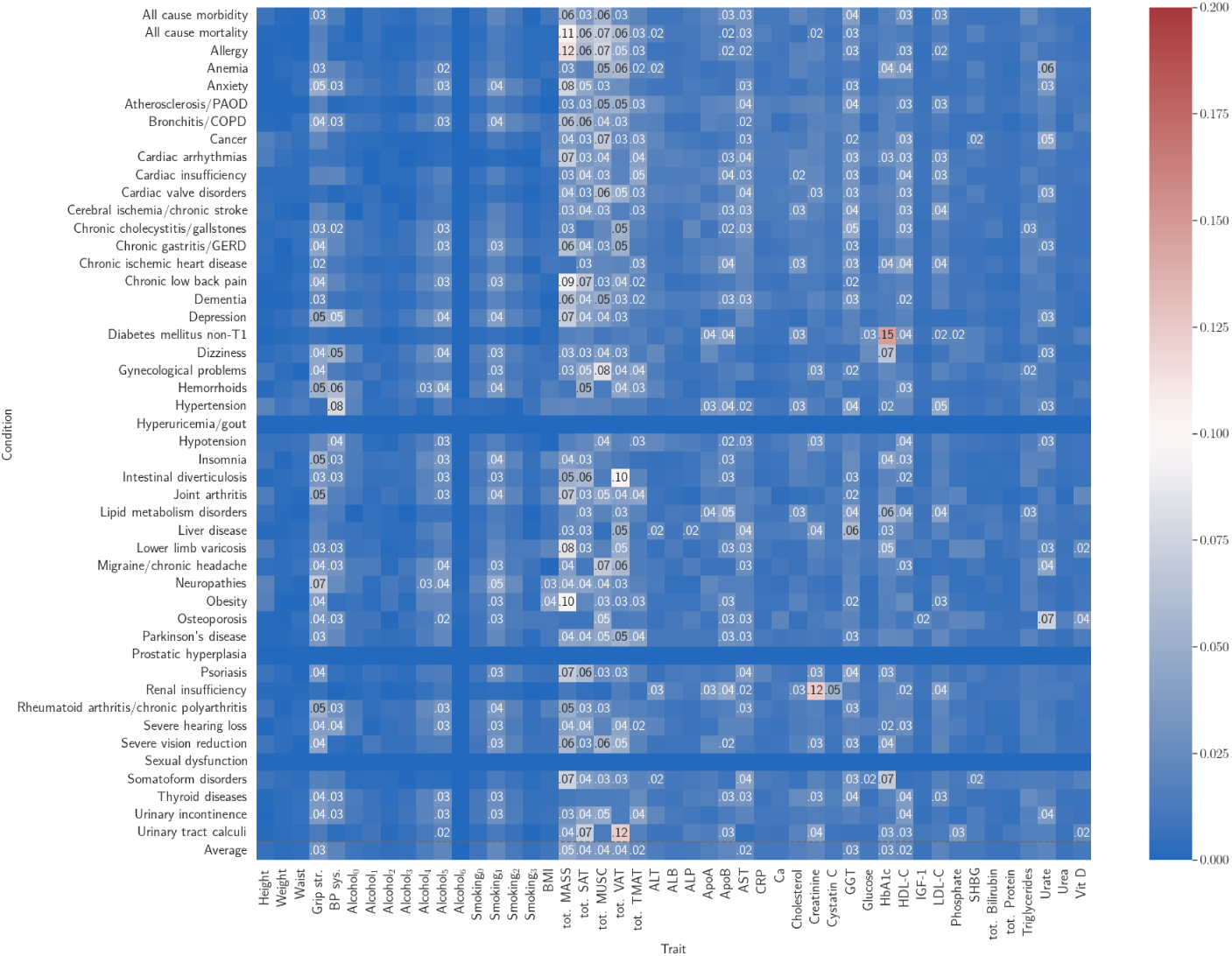
Univariate saliency for full traits model (female). Top 10 traits for each row are labelled.

**Fig. C25:**
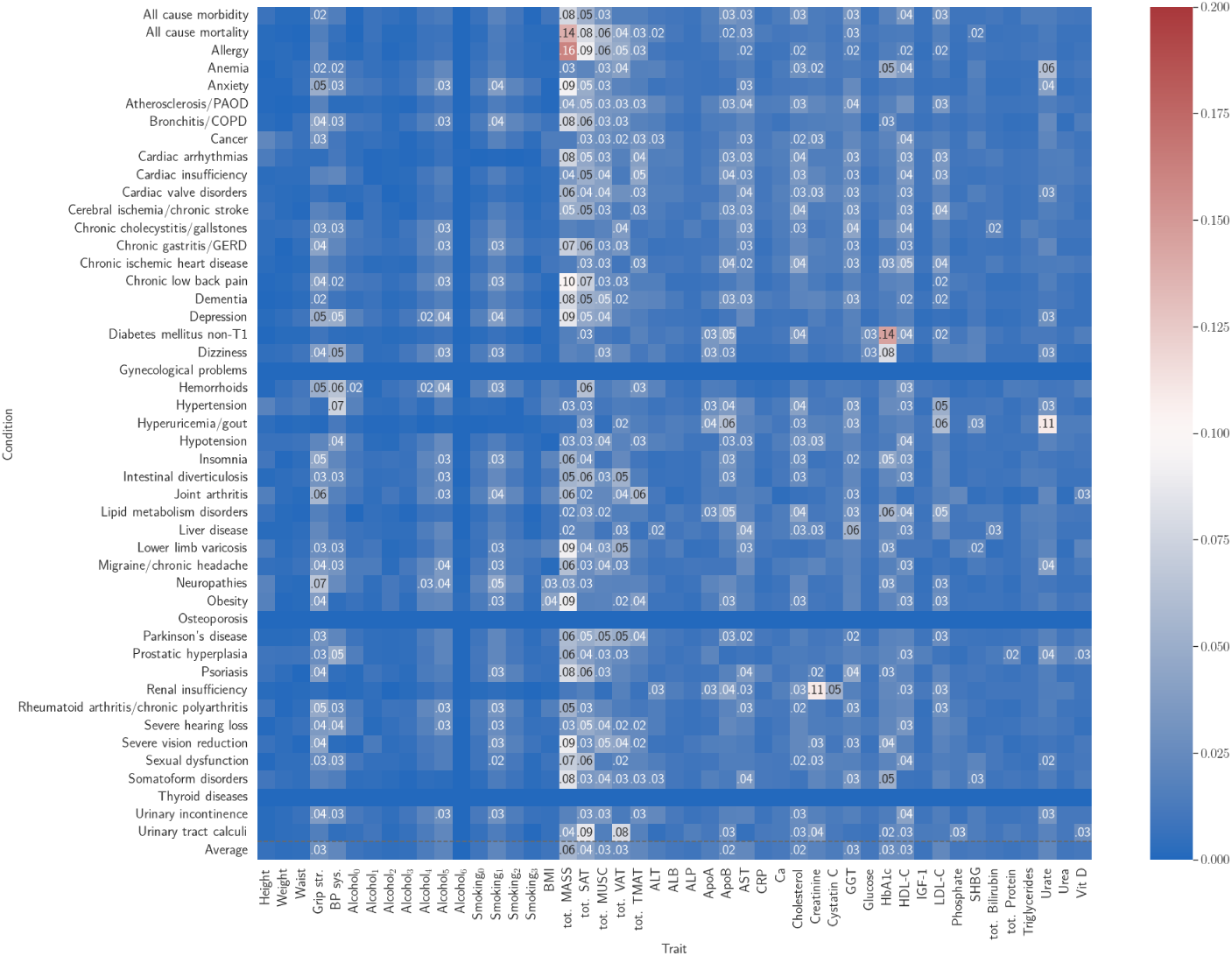
Univariate saliency for full traits model (male). Top 10 traits for each row are labelled.

**Fig. C26:**
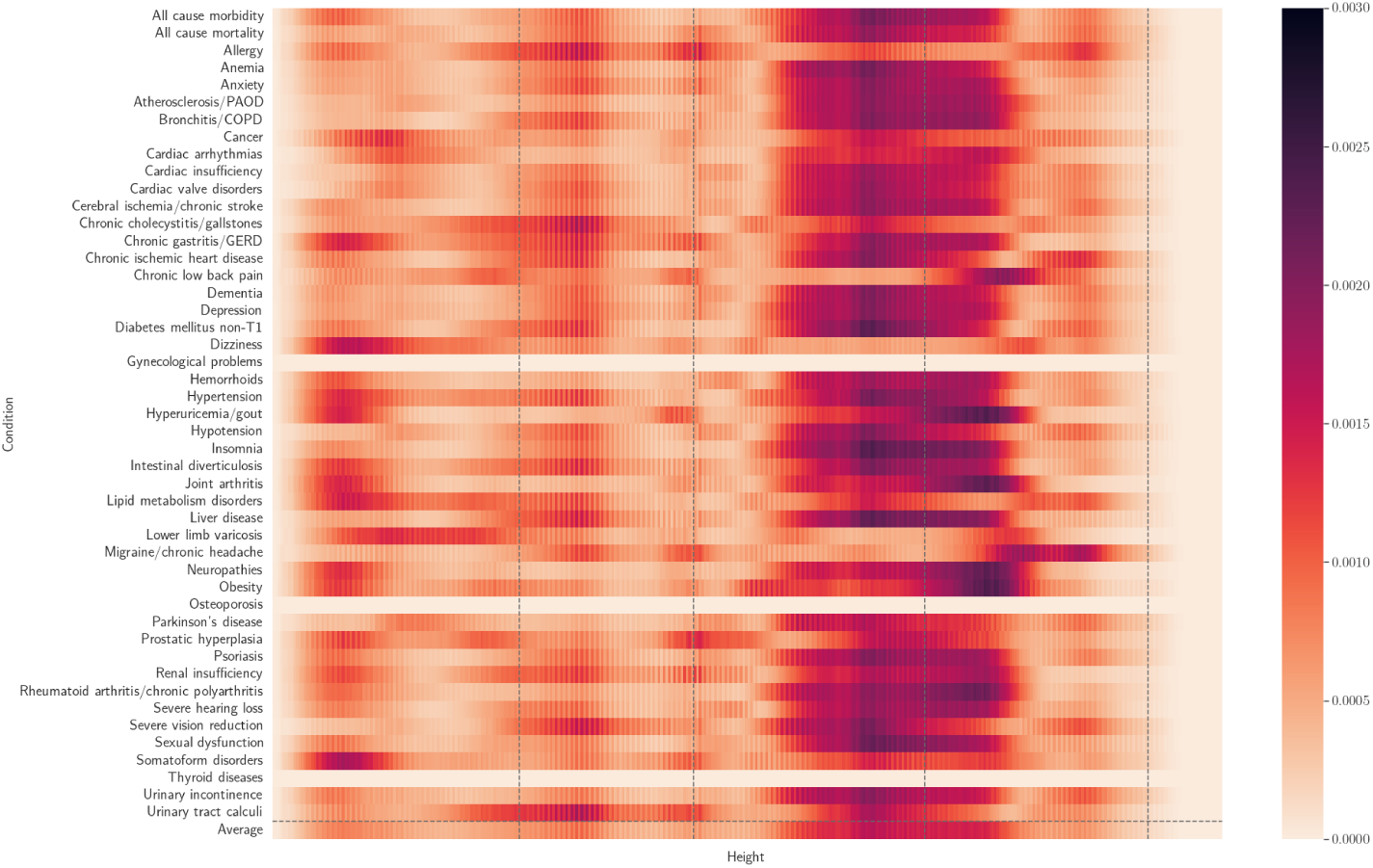
Spatial body composition saliency across conditions (male MASS).

**Fig. C27:**
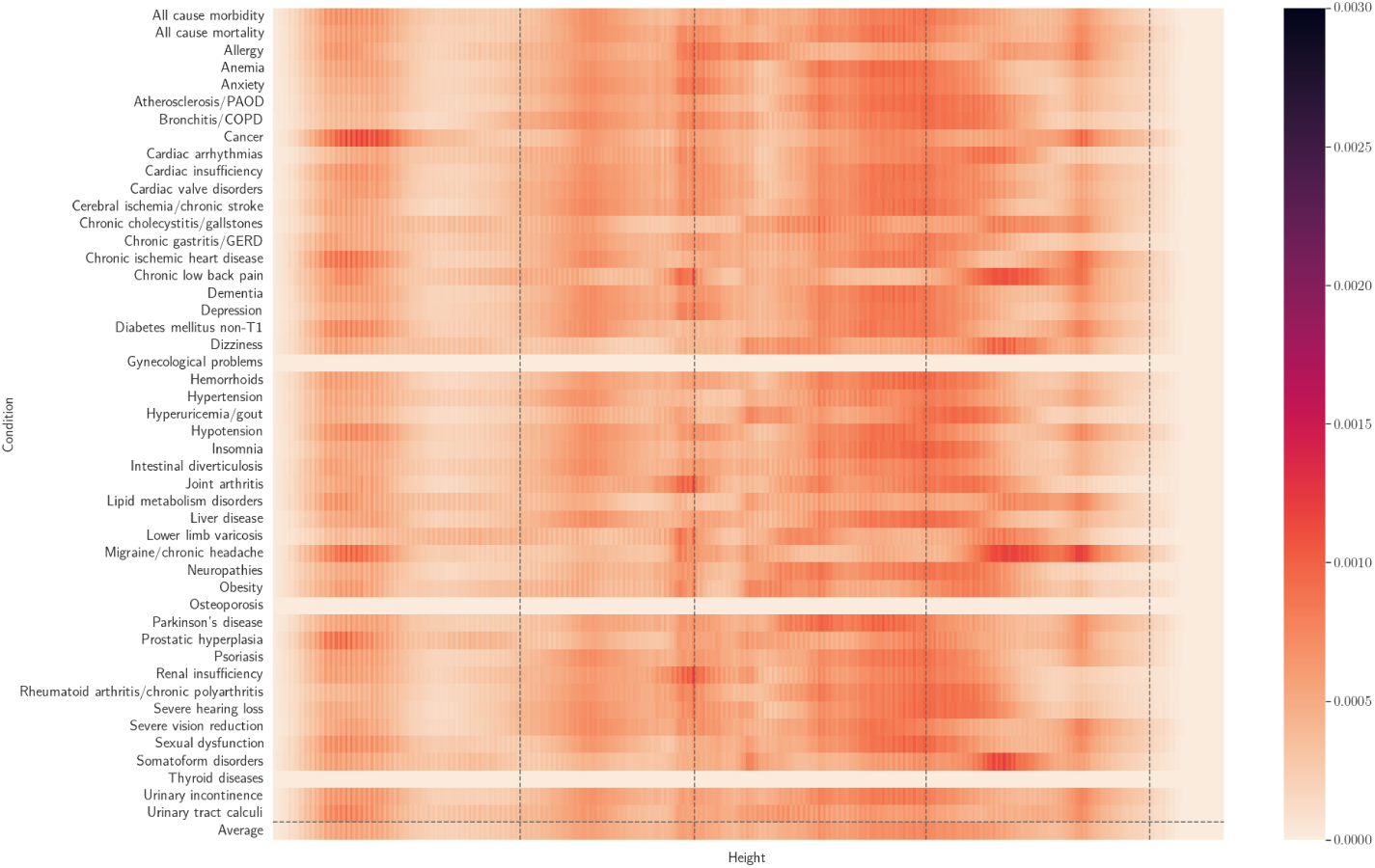
Spatial body composition saliency across conditions (male MUSC).

**Fig. C28:**
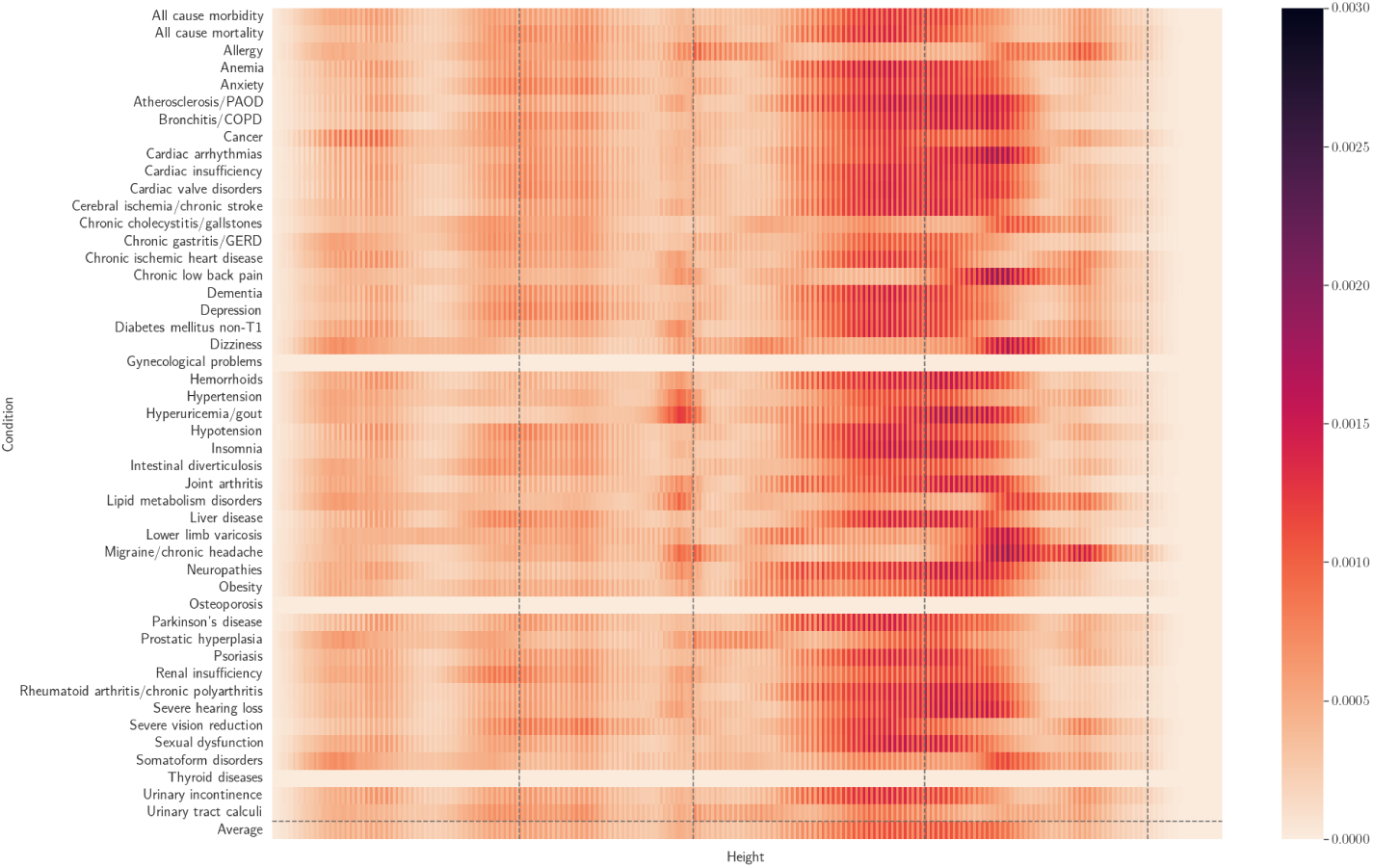
Spatial body composition saliency across conditions (male SAT).

**Fig. C29:**
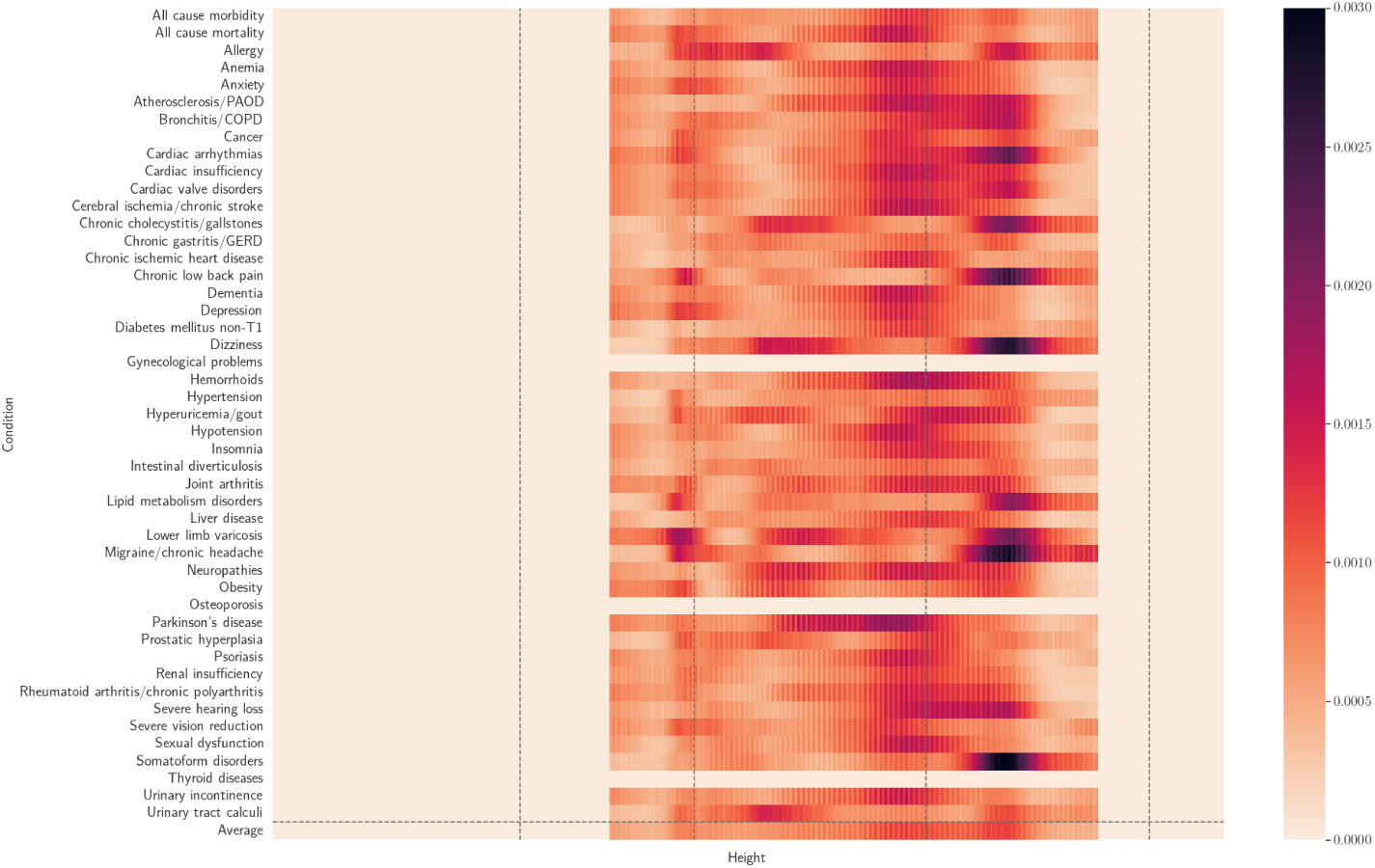
Spatial body composition saliency across conditions (male VAT).

**Fig. C30:**
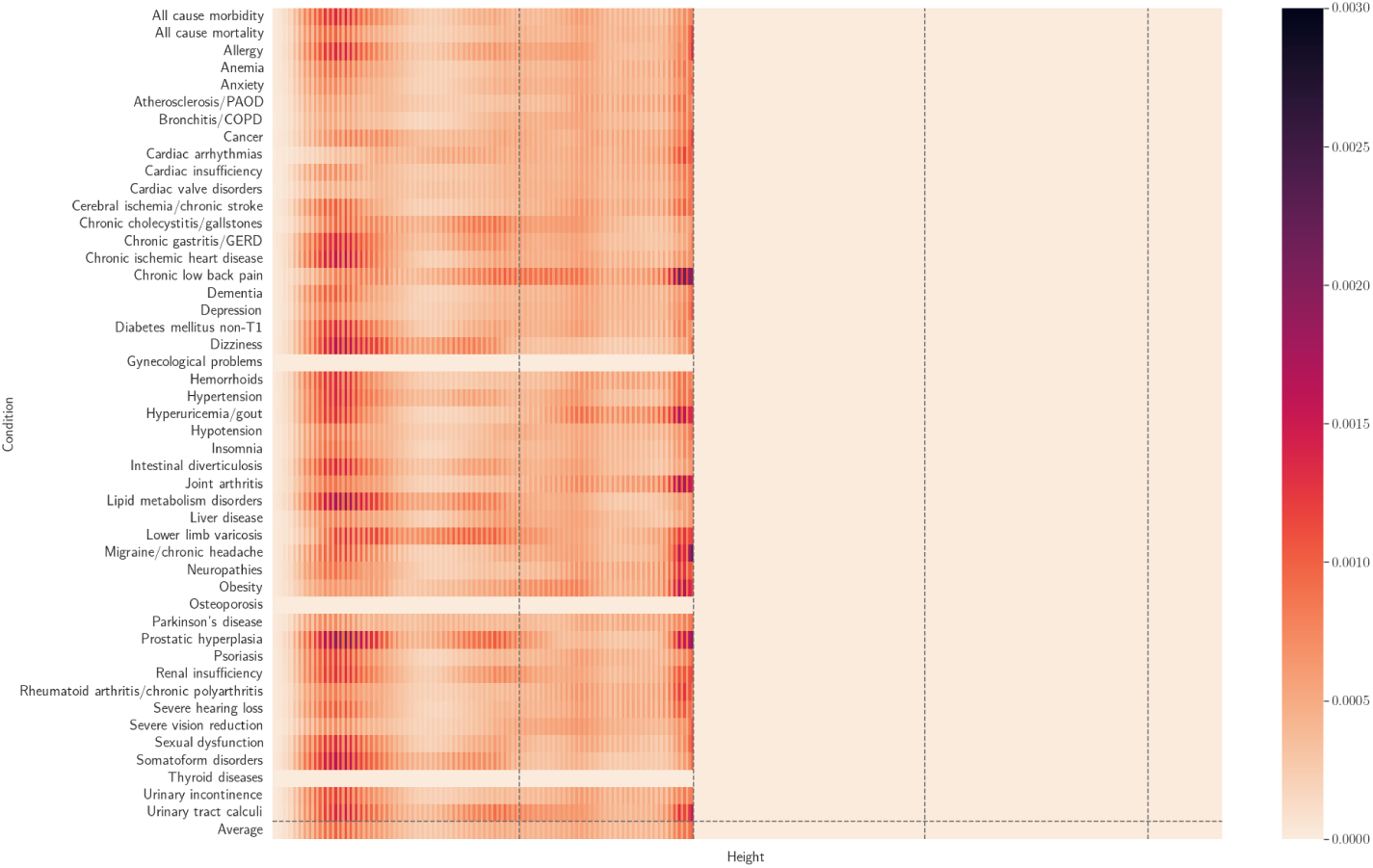
Spatial body composition saliency across conditions (male TMAT).

**Fig. C31:**
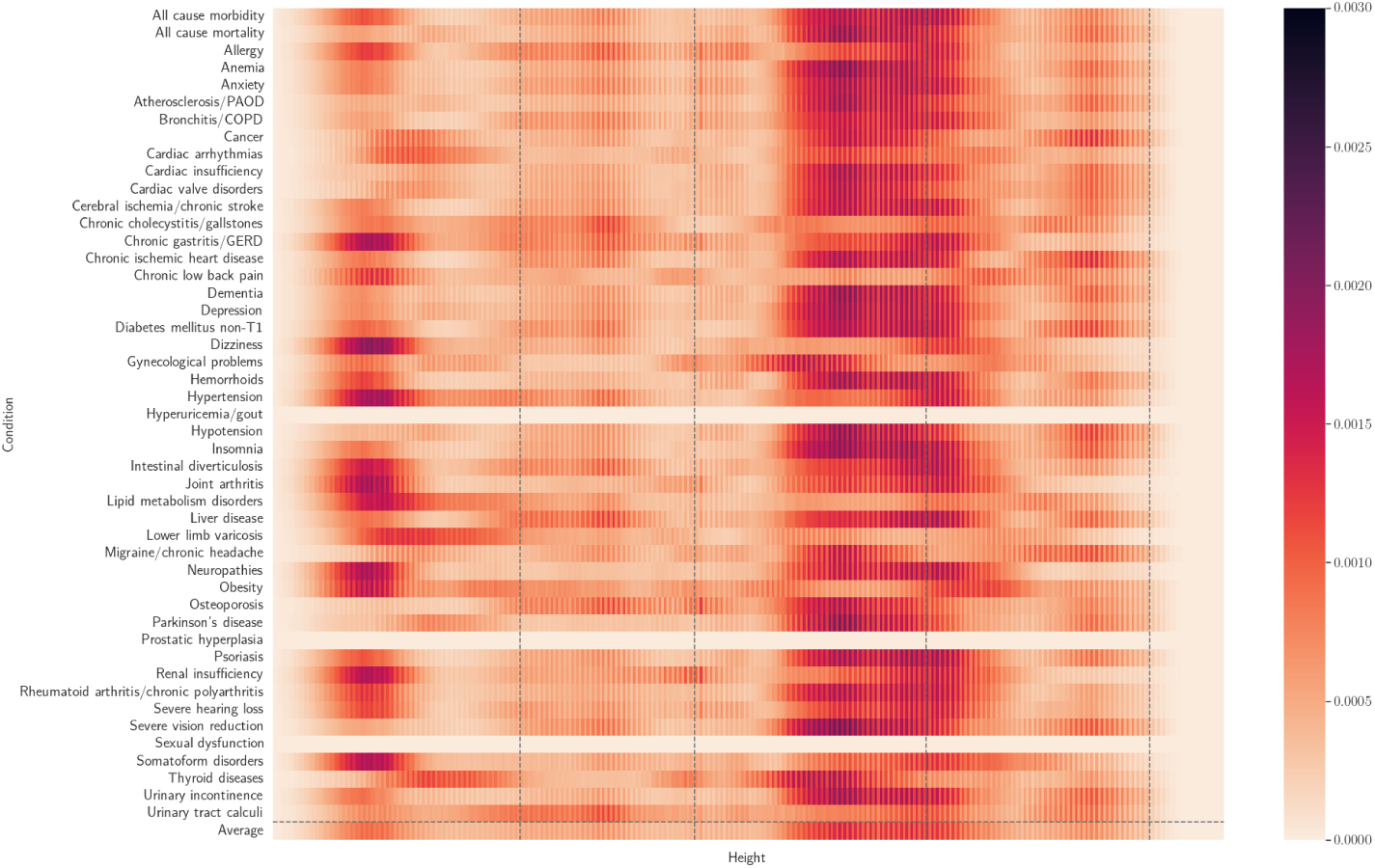
Spatial body composition saliency across conditions (female MASS).

**Fig. C32:**
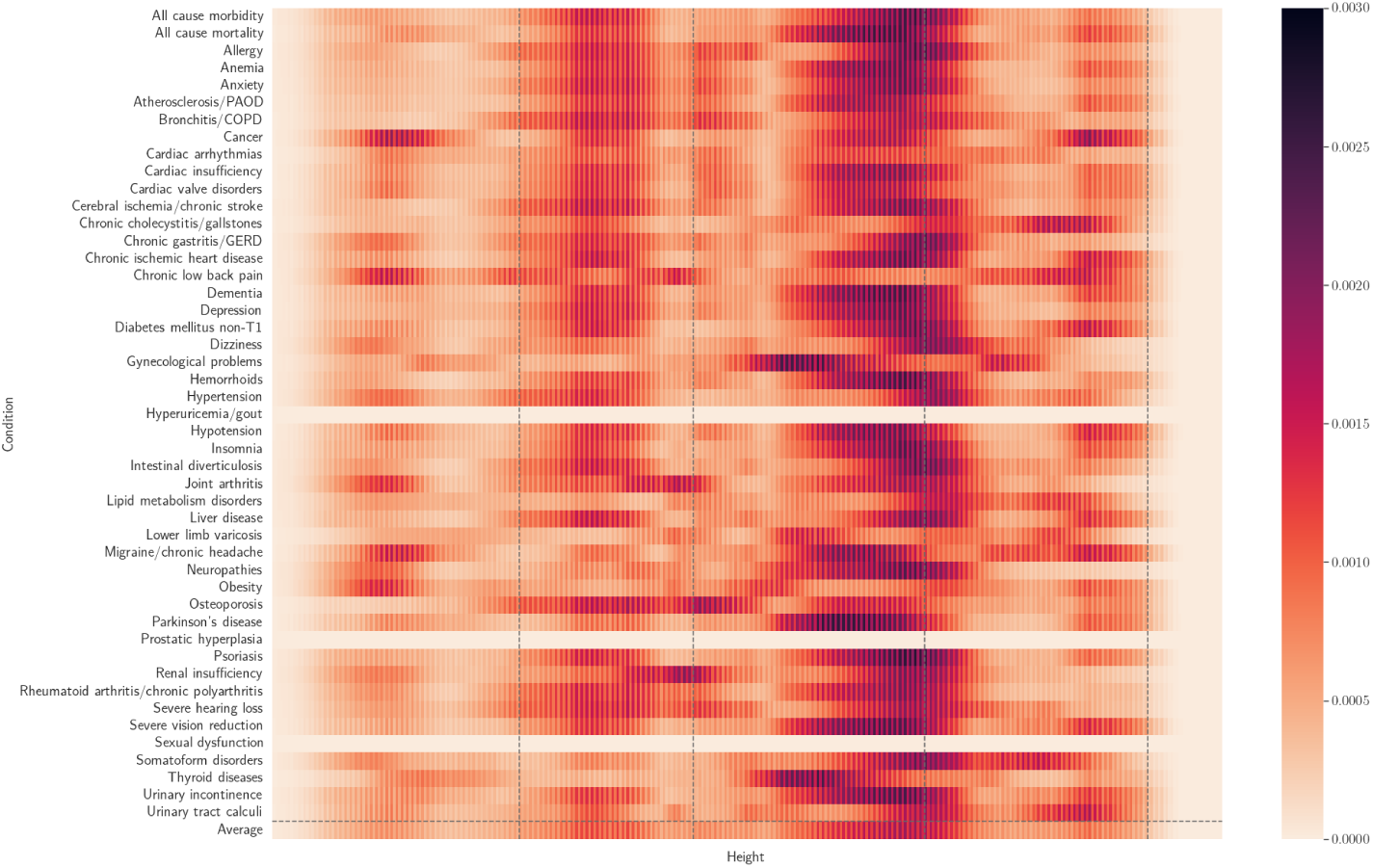
Spatial body composition saliency across conditions (female MUSC).

**Fig. C33:**
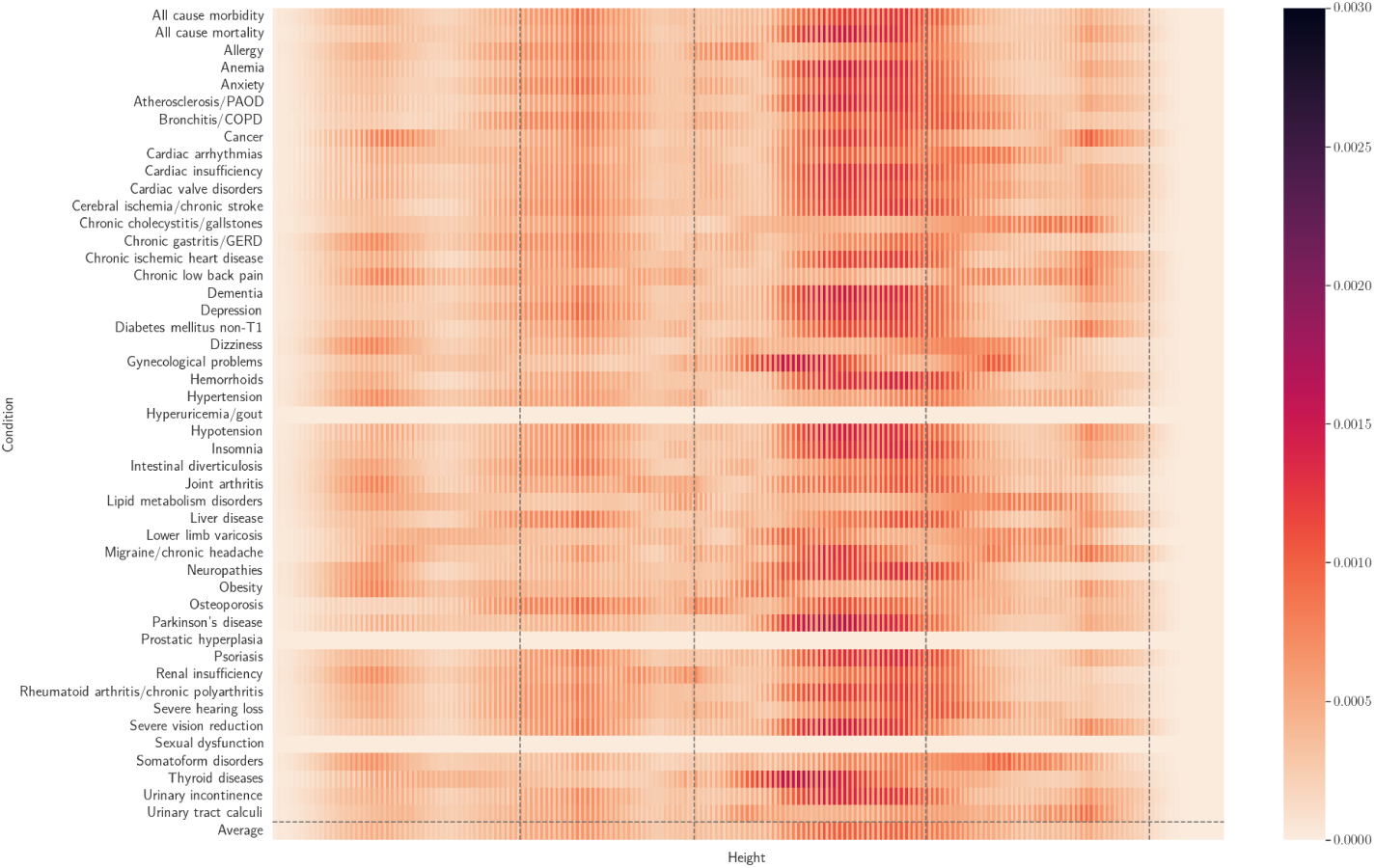
Spatial body composition saliency across conditions (female SAT).

**Fig. C34:**
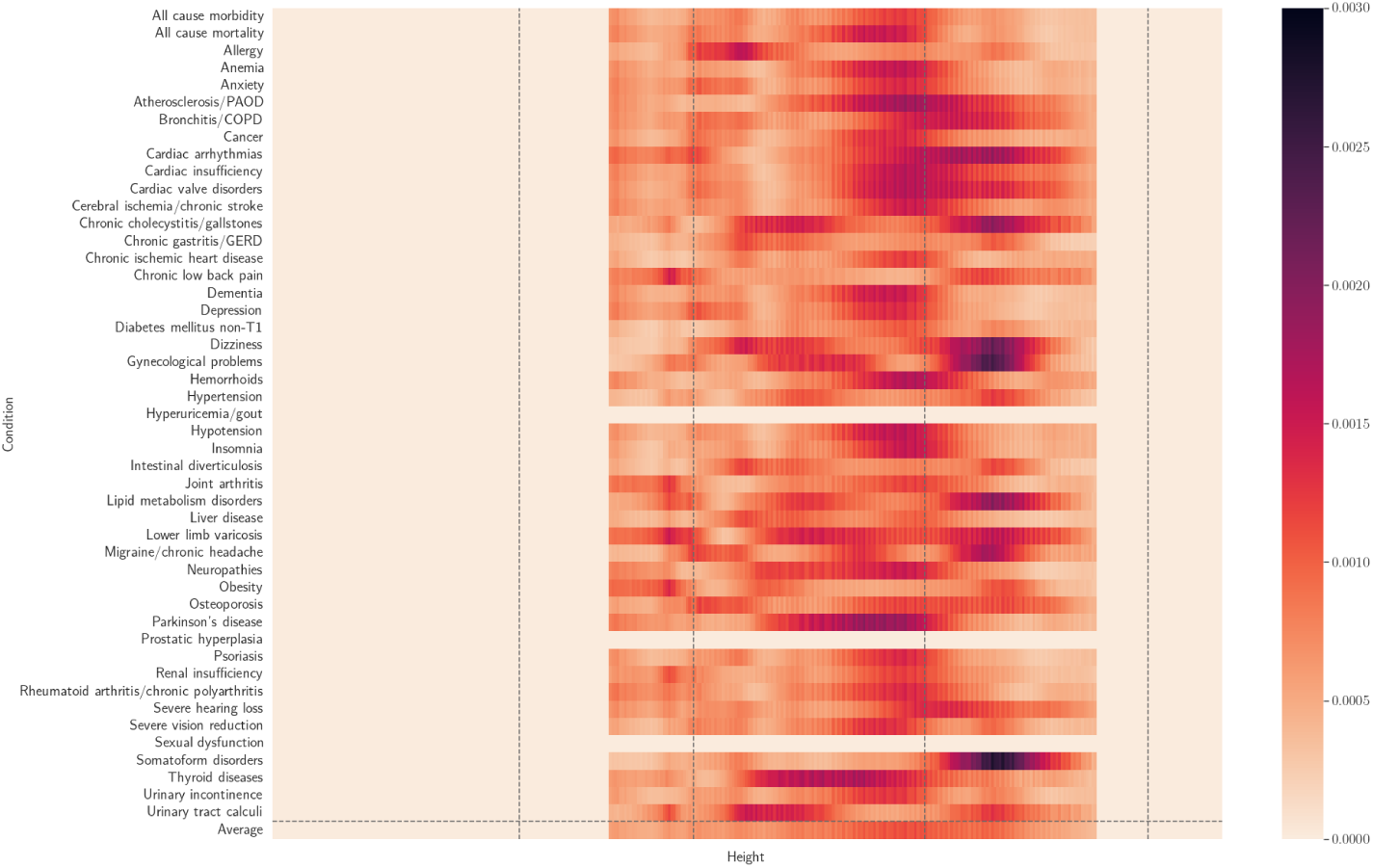
Spatial body composition saliency across conditions (female VAT).

**Fig. C35:**
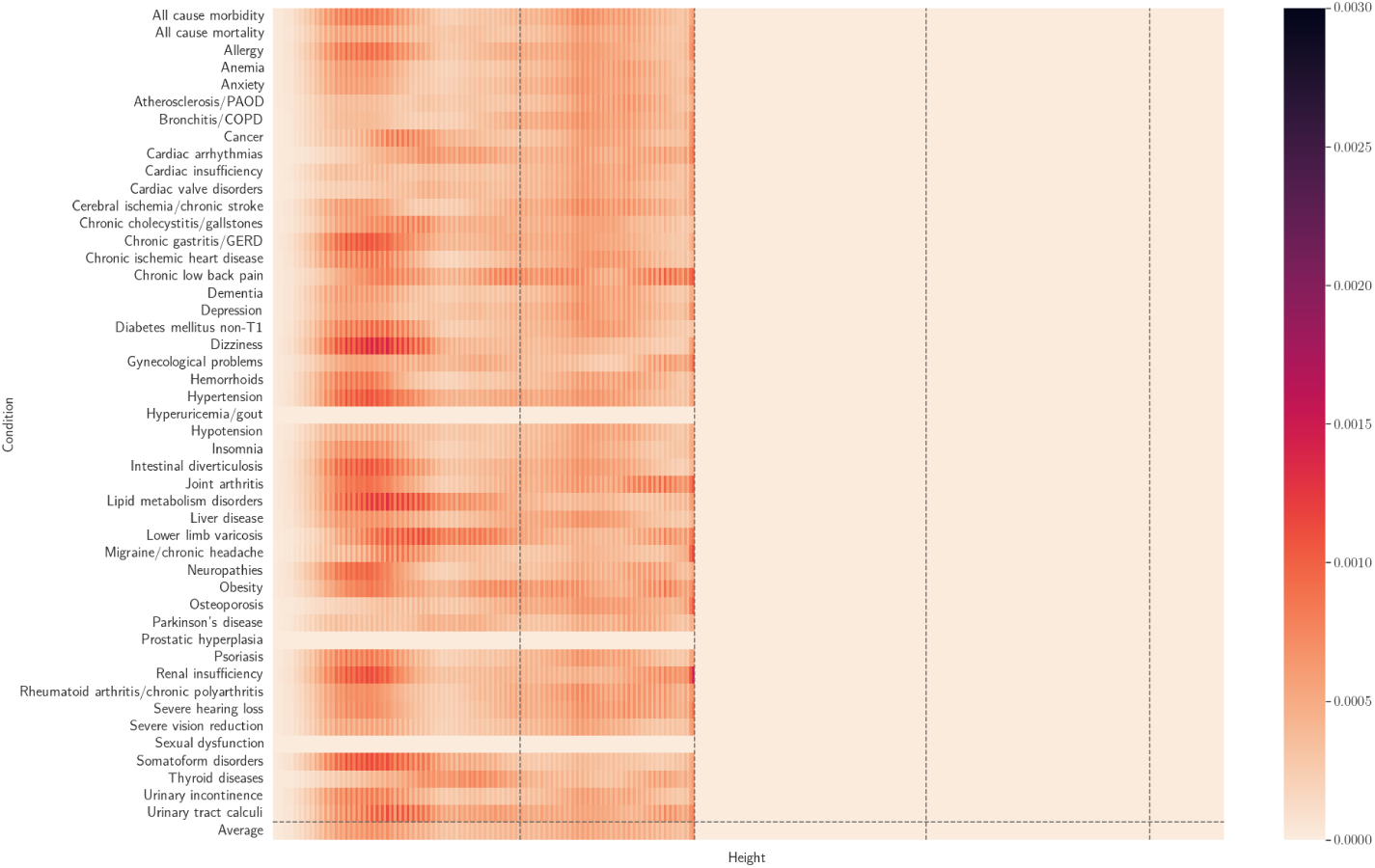
Spatial body composition saliency across conditions (female TMAT).

#### C.5.2 Full univariate traits model

**Fig. C36:**
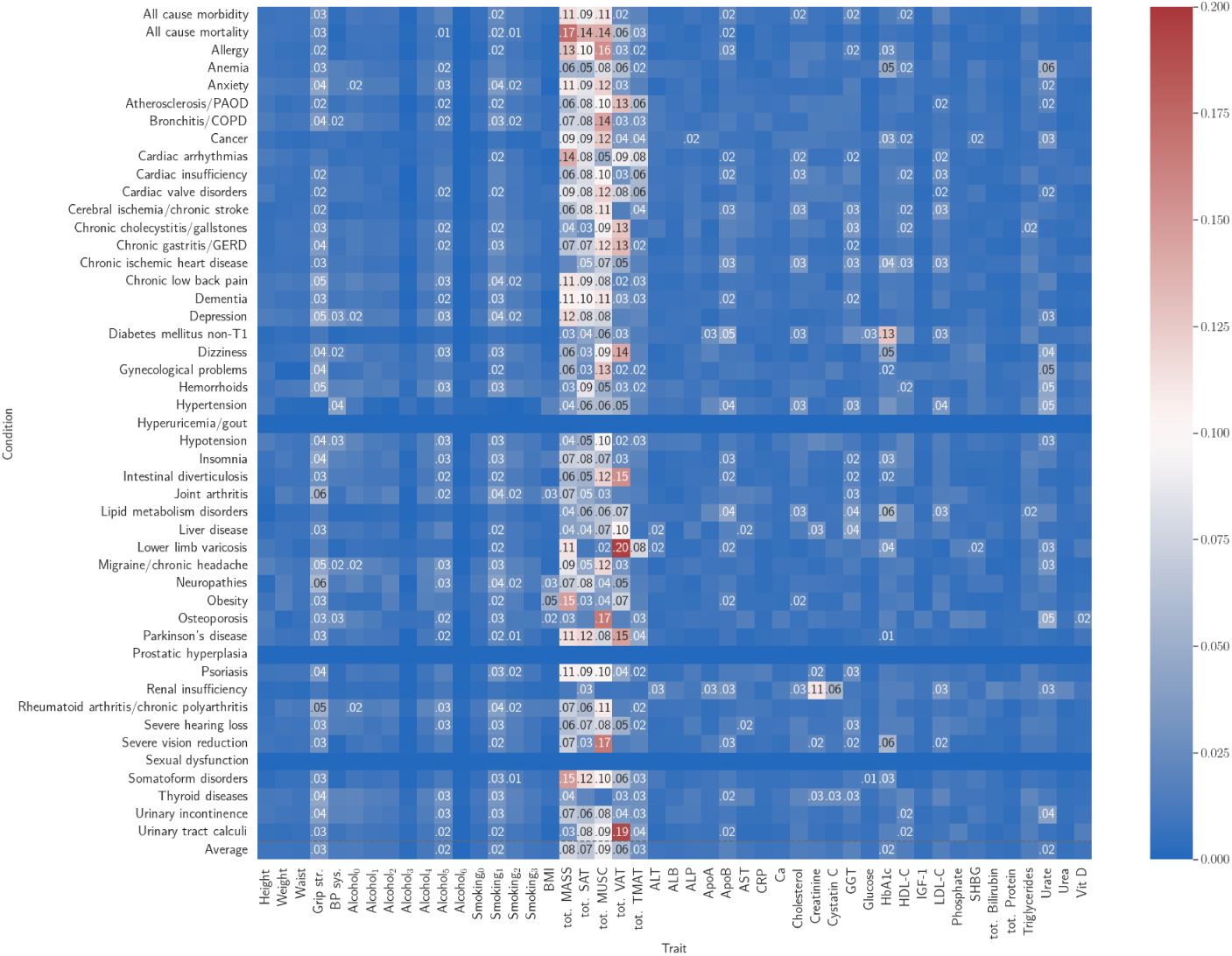
Univariate saliency for full univariate traits model (female). Top 10 traits for each row are labelled.

**Fig. C37:**
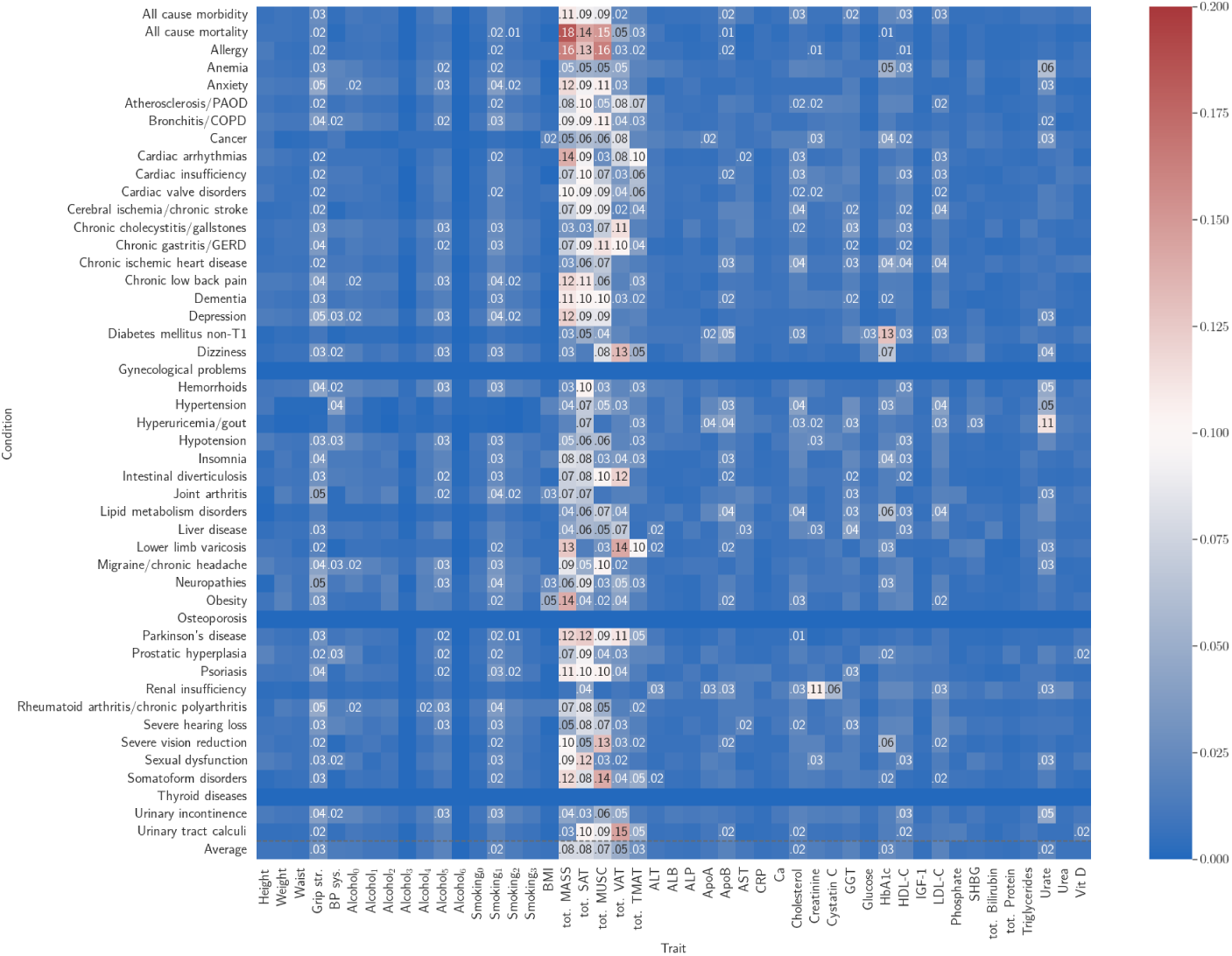
Univariate saliency for full univariate traits model (male). Top 10 traits for each row are labelled.

## Appendix D Reporting checklists

| Section and item | Checklist item |
| --- | --- |
| **Title** |  |
| 1 x | Identify the study as developing or evaluating the performance of a multivariable prediction model, the target population, and the outcome to be predicted |
| **Background** |  |
| 2 x | Provide a brief explanation of the healthcare context and rationale for developing or evaluating the performance of all models |
| **Objectives** |  |
| 3 x | Specify the study objectives, including whether the study describes model development, evaluation, or both |
| **Methods** |  |
| 4 x | Describe the sources of data |
| 5 x | Describe the eligibility criteria and setting where the data were collected |
| 6 x | Specify the outcome to be predicted by the model, including time horizon of predictions in case of prognostic models |
| 7 x | Specify the type of model, a summary of the model-building steps, and the method for internal validation† |
| 8 x | Specify the measures used to assess model performance (eg, discrimination, calibration, clinical utility) |
| **Results** |  |
| 9 x | Report the number of participants and outcome events |
| 10 x | Summarise the predictors in the final model† |
| 11 x | Report model performance estimates (with confidence intervals) |
| **Discussion** |  |
| 12 x | Give an overall interpretation of the main results |
| **Registration** |  |
| 13 N/A | Give the registration number and name of the registry or repository |

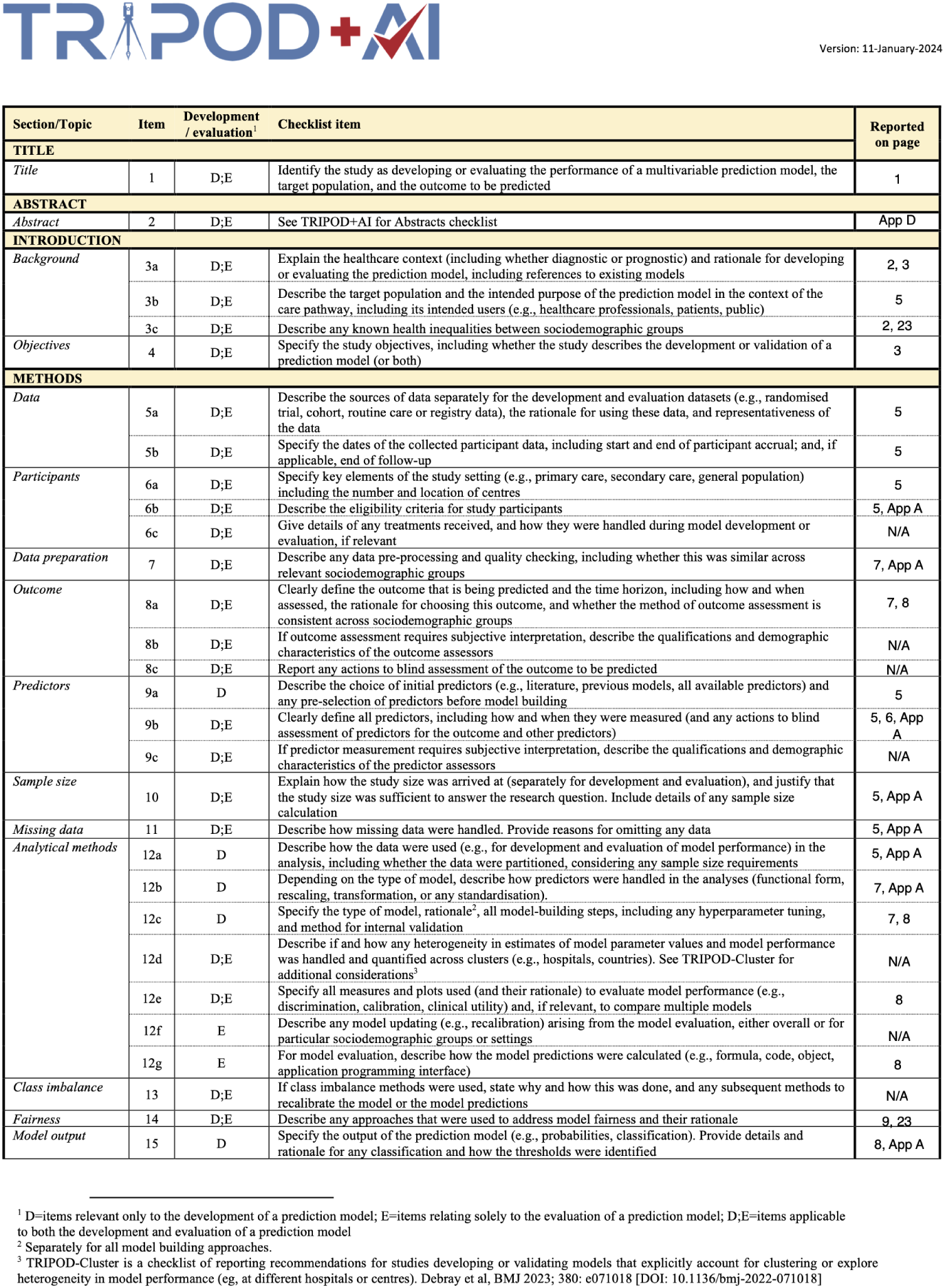

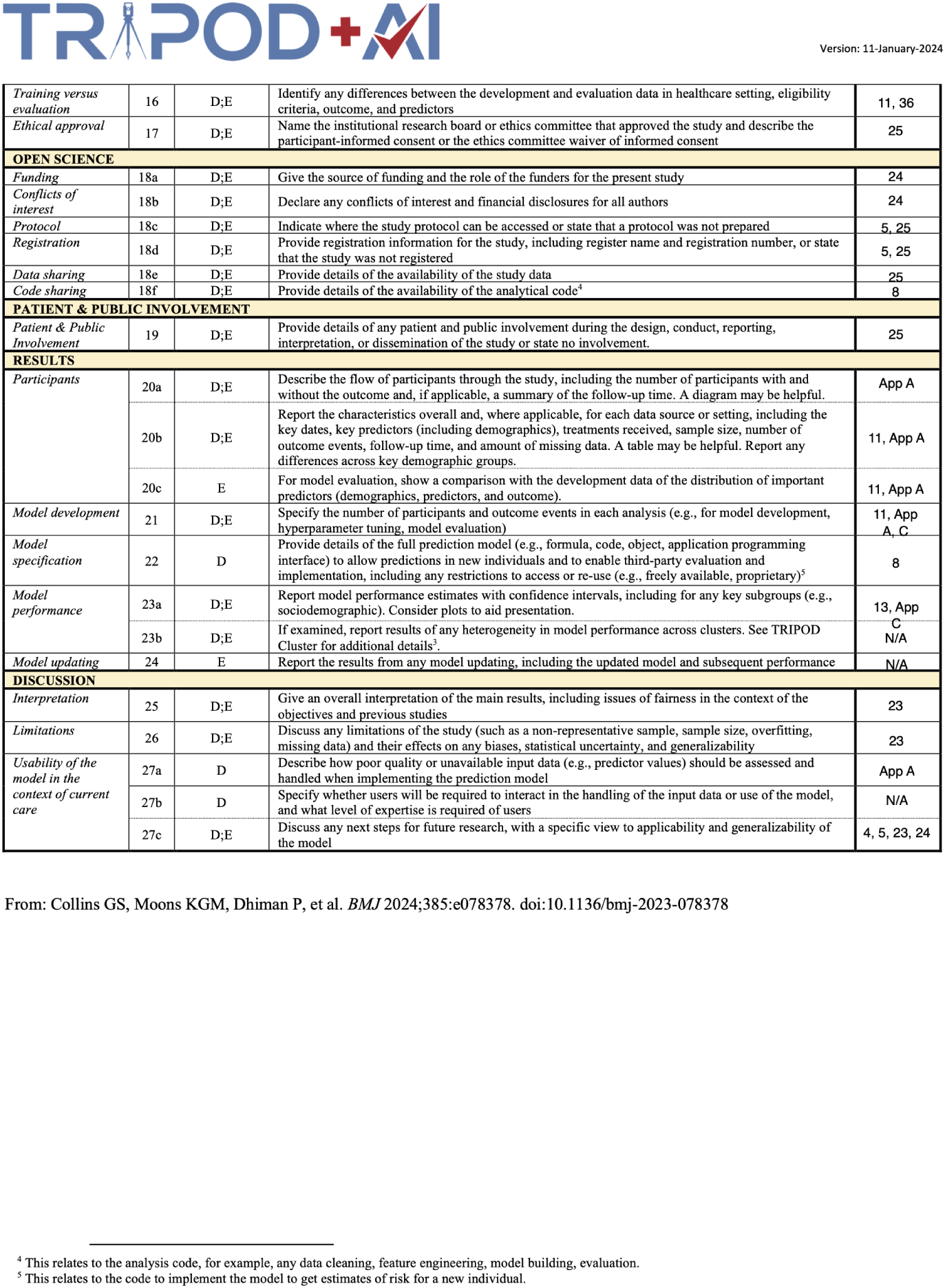

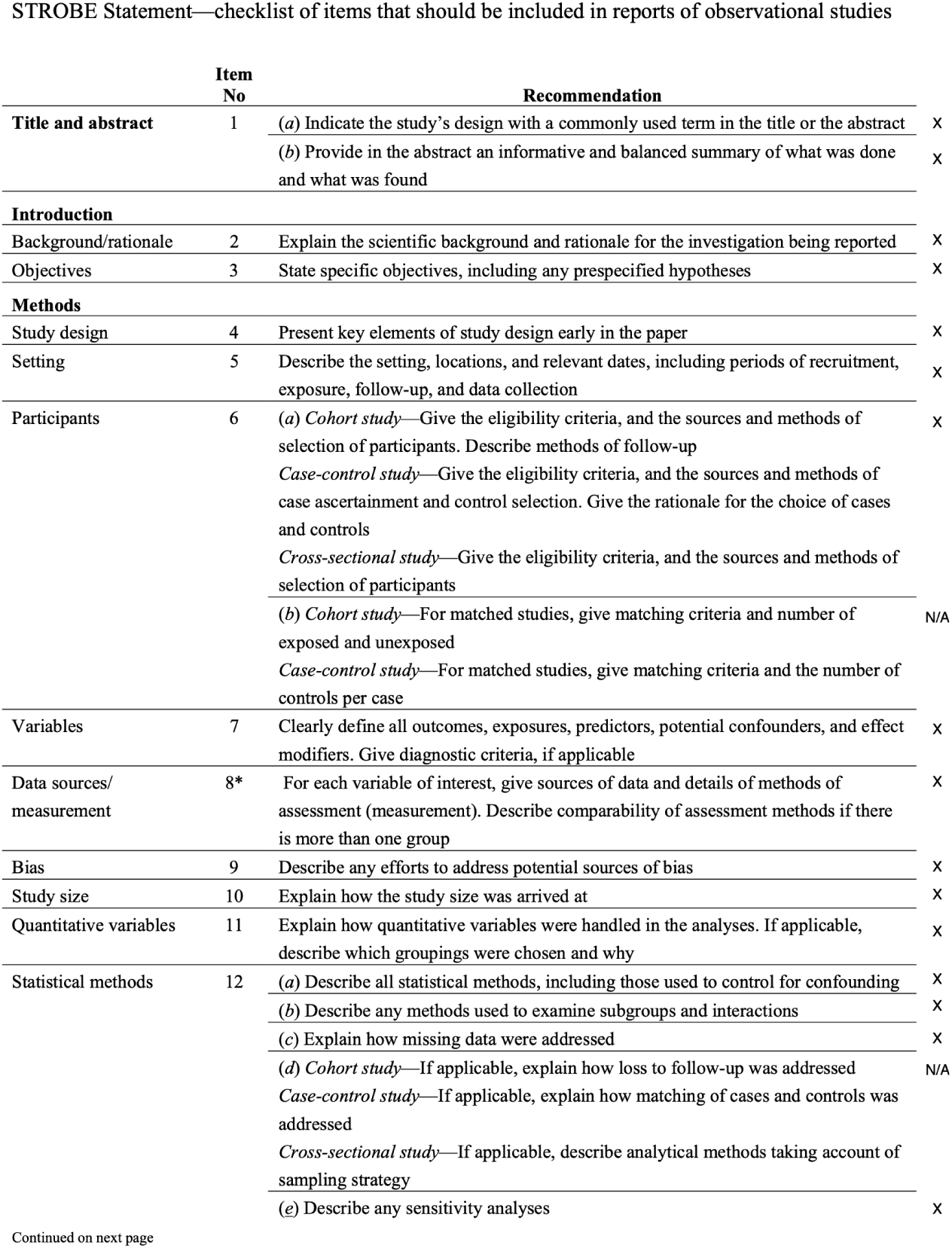

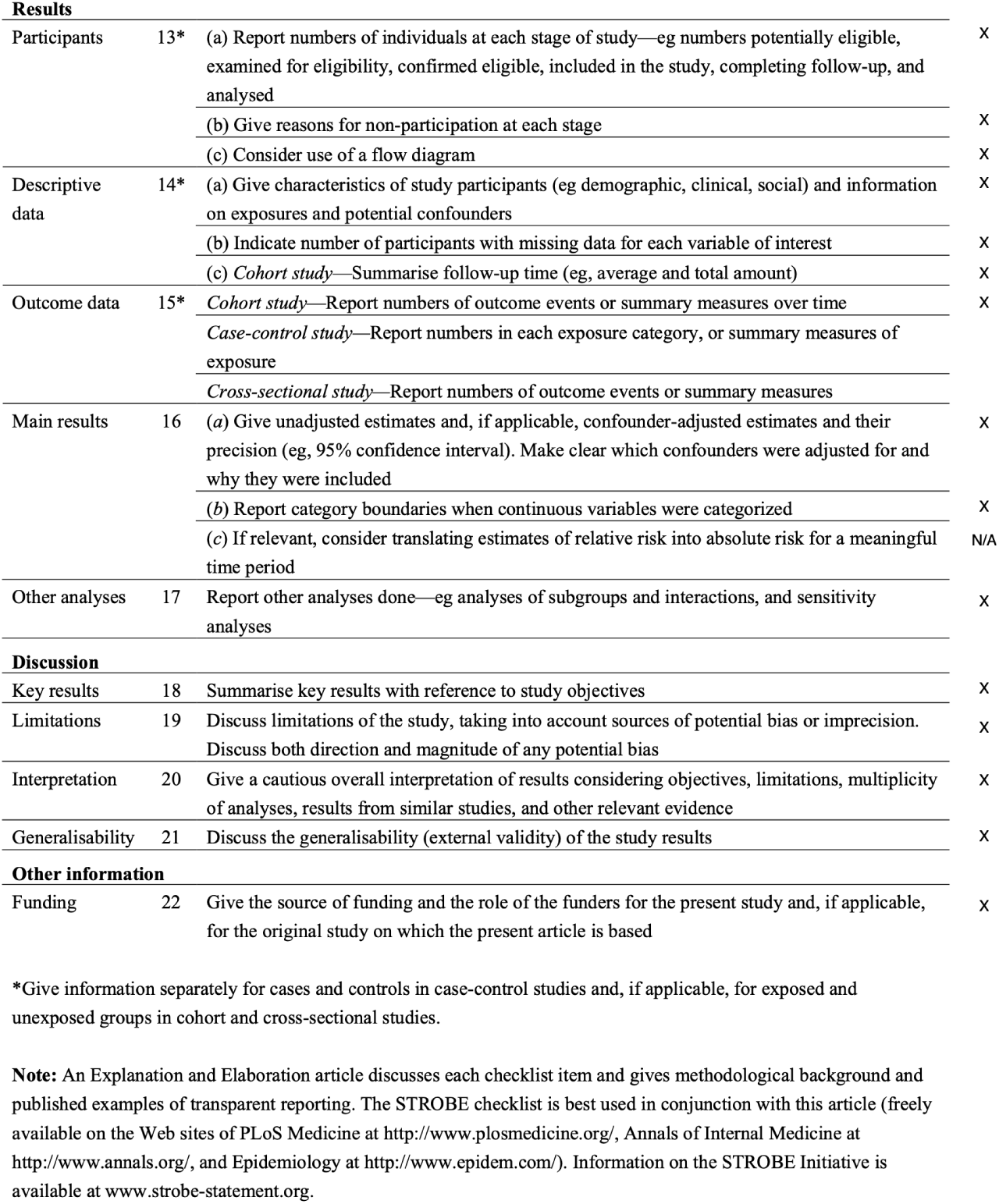

